# RFM: A Lightweight Retinal Foundation Model for Generalised Oculomics

**DOI:** 10.64898/2026.10.02.26362554

**Authors:** Jiri Fajtl, Roshan Welikala, Royce Shakespeare, Ryan Chambers, Louis Bolter, John Anderson, Paolo Remagnino, Arya Pradeep Menon, Gordon Johnson, Farzana Rahman, Abraham Olvera-Barrios, Alasdair Warwick, Catherine Egan, Adnan Tufail, Christopher G Owen, Alicja R Rudnicka, Sarah Barman, UK Biobank Eye and Vision Consortium, ARIAS Research Group

## Abstract

Foundation models for colour fundus photography (CFP) are candidate visual encoders for multimodal medical AI, but existing models are large and are assessed under fine-tuning rather than the frozen-encoder conditions such systems impose. We present RFM (Retinal Foundation Model), a 64-million-parameter Vision Transformer initialised from DINOv3-Base and domain-adapted with the LeJEPA joint-embedding predictive self-supervised objective with domain-specific pretext tasks, on 4.3 million CFPs from the North East London Diabetic Eye Screening Programme. A single linear head per task on one frozen backbone measures representation generalisability, not fine-tuning capacity. Across six external datasets spanning diabetic retinopathy (DR) grading, glaucoma detection, ten UK Biobank biomarker phenotypes and 3-year incidence of myocardial infarction, ischaemic stroke and Parkinson’s disease, RFM attains the lowest mean absolute error of any frozen model on all seven regression phenotypes and a higher area under the receiver operating characteristic curve (AUROC) than any RETFound variant on all eight matched classification benchmarks. It also attains the highest AUROC of any model evaluated on four of the five ophthalmic benchmarks, with 4.8 × fewer parameters than RETFound-MAE and RETFound-DINOv2. Applied to UK Biobank without retraining, RFM exceeds the best published retinal foundation-model AUROC for 3-year myocardial infarction and ischaemic stroke; the Parkinson’s endpoint (11 cases) is inconclusive. DINOv3-Base without retinal pre-training, read the same way, stays competitive on DR grading but falls clearly behind on glaucoma, and the feature-extraction convention alone is worth up to 0.154 AUROC. One lightweight backbone yields generalised representations serving ophthalmic grading, biomarker regression and incidence prediction.

## 1. Introduction

Colour fundus photography (CFP) provides non-invasive optical access to the retina, simultaneously imaging the retinal vasculature, neural architecture, and optic disc, structures that share embryological origins and haemodynamic coupling with the cerebral and systemic vasculature [1]. National screening programmes have accumulated large image archives well suited to self-supervised pre-training. Beyond ophthalmic disease detection, the retinal fundus also encodes information relevant to systemic physiology, including cardiovascular risk and glycaemic control [2] and neurodegenerative pathology [3].

Despite rapid growth in the number of AI models proposed for retinal imaging, translation into deployed clinical benefit remains limited [4, 5]. One barrier is size: current foundation models are computationally demanding, which constrains point-of-care and screening-scale deployment.

Deep learning analysis of retinal fundus images has matured from task-specific convolutional networks [6, 7] to foundation models pre-trained by self-supervised learning (SSL) on large unlabelled datasets [8, 9]. The prevailing paradigm, exemplified by RETFound [8], trains a large Vision Transformer (ViT) backbone on millions of unlabelled fundus images using masked image modelling, then fine-tunes the model on individual downstream tasks with labelled data and non-linear prediction heads. While effective, this paradigm carries three constraints on deployment breadth: the fine-tuned models are task-specific and cannot generalise across endpoints without retraining; existing retinal foundation models are large (307 million parameters for RETFound), increasing inference cost and memory footprint; and benchmarking has been limited to a narrow set of ophthalmic and incidence endpoints, leaving the breadth of frozen-feature generalisation poorly characterised. How pre-training data composition modulates performance has been examined directly [10]; how well frozen features carry across diverse clinical endpoints has not.

A compelling application horizon for retinal foundation mod-els is their use as visual encoders in multimodal architectures that couple CFPs with electronic health records, clinical text or genomics [11]. There the vision encoder cannot be retrained per task: one frozen backbone must serve every predictive target at once. Evaluating exclusively under that constraint tests whether the representations are linearly separable and task-transferable, an architecture-neutral measure of quality, and makes frozen-linear performance across a broad task spectrum the criterion that matters for such a model rather than its fine-tuning capacity on any one task.

We present RFM (Retinal Foundation Model), a 64-million-parameter DINOv3-Base ViT pre-trained with the LeJEPA joint-embedding predictive self-supervised objective [12] on 4.3 million CFP images from the North East London Diabetic Eye Screening Programme (DESP) [13]. We propose deliberately simple domain-specific pretext tasks that combine standard global crops with circular-area-constrained local crops designed to capture multi-scale retinal features, and a projector adaptation warmup schedule intended to keep early random gradient propagation from degrading the initialised DINOv3 representations. To our knowledge this is the first application of LeJEPA to retinal imaging; the only prior biomedical-signal application is by Panchavati et al. [14], on EEG.

We evaluate RFM under the frozen-backbone linear-head protocol on six external datasets: diabetic retinopathy (DR) grading on APTOS 2019, IDRiD and Messidor-2, glaucoma detection on GFID [15] and PAPILA [16], ten UK Biobank [17] biomarker phenotypes, and 3-year incidence of myocardial infarction (MI), ischaemic stroke and Parkinson’s disease on internal (DESP) and external (UKBB) cohorts. The un-adapted natural-image DINOv3-Base checkpoint [18], read exactly as RFM is (DINOv3b-9L), is evaluated under the identical protocol as a baseline; it remains competitive with RFM on DR grading while falling clearly behind on glaucoma, a task-dependent pattern that carries implications for retinal SSL pretext task design.

The principal contributions of this work are:

1. The first application of the LeJEPA self-supervised objective to retinal imaging, yielding a model at approximately one-fifth the parameter count of RETFound-MAE and RETFound-DINOv2 that leads every RETFound variant on the matched regression and classification benchmarks (Section 4).
2. Domain-specific LeJEPA pretext tasks for CFP incorporating circular-area-constrained local crops, a projector adaptation warmup schedule and a prediction centre averaged over all views rather than the global views alone, adopted on the basis of pilot experiments to address early-training instability.
3. The broadest frozen-linear evaluation of a retinal foundation model we are aware of, spanning six datasets, ten UKBB biomarker phenotypes, and three systemic incidence endpoints, with external cohort transfer validation.
4. A controlled analysis of the frozen-linear evaluation recipe itself, isolating the feature-extraction convention as an effect worth up to 0.154 AUROC on the UK Biobank phenotypes and large enough to reorder close comparisons on the ophthalmic benchmarks, and characterising where a natural-image backbone still competes with retinal-domain pre-training.

The remainder of this paper is organised as follows. Section 2 reviews SSL methodology and retinal foundation model evaluation. Section 3 describes the RFM architecture, pre-training procedure, datasets, and evaluation protocol. Section 4 presents quantitative results across all tasks. Section 5 analyses the findings and discusses limitations. Section 6 provides overall conclusions of the work.

## 2. Related Work

### 2.1. Foundation Models and Self-Supervised Representation Learning for Retinal Imaging

Foundation models – large networks pre-trained on broad data and adapted to many downstream tasks [19] – reached vision through the Vision Transformer [20] and self-supervised pre-training, which suits medical imaging because labels are scarce and clinically heterogeneous.

Two families dominate. Joint-embedding methods learn invariance between augmented views: SimCLR [21], MoCo [22] and BYOL [23] established that strong augmentation policies, momentum-updated encoders and student–teacher architectures each yield high-quality representations with or without explicit negative pairs; DINO [24] brought the student–teacher form to Vision Transformers, showing emergent semantic segmentation properties without pixel-level annotation, and DINOv2 [25] scaled it to a curated corpus of 142 million web images, with DINOv3 [18] extending that lineage. Masked image modelling instead reconstructs: Masked Autoencoders [26] mask 75% of patches and rebuild the raw pixels. LeJEPA [12], the objective used here, belongs to neither family: it predicts in representation space and prevents collapse through an explicit distributional regulariser rather than the engineering heuristics the joint-embedding methods rely on (Section 3.1), which matters for domain-specific pre-training where extensive hyperparameter search is computationally costly.

Medical imaging adds domain shift, annotation scarcity and structurally heterogeneous downstream tasks. Tham et al. [11] describe a globally representative medical foundation model pre-trained across multiple geographic cohorts, arguing that demographic and clinical diversity in the pre-training corpus is a prerequisite for equitable cross-population generalisation.

RETFound [8] established the reference benchmark for self-supervised retinal foundation models: a ViT-Large backbone pre-trained with a Masked Autoencoder on approximately one million unlabelled colour fundus photographs from Moorfields Eye Hospital and Kaggle EyePACS, reaching state-of-the-art performance across ophthalmic and systemic endpoints through full fine-tuning. RETFound-Green [9] then matched or exceeded it with far less data (75,000 public retinal images) and roughly 400-fold less compute, challenging the premise that data volume is the primary determinant of representation quality in domain-specific SSL. The role of pre-training data composition was examined systematically by Zhou et al. [10], who evaluated how source population characteristics, imaging hardware, and clinical case mix modulate downstream performance using two large fundus cohorts. That analysis identified a non-trivial interaction between the pre-training domain and evaluation task: clinically diverse pre-training improved generalisation to external sites, while tasks requiring fine-grained morphology were sensitive to imaging protocol consistency.

### 2.2 Retinal Foundation Models on Downstream Clinical and Biomarker Prediction Tasks

Automated fundus analysis for ophthalmic disease detection dates to Gulshan et al. [6], whose convolutional network reached specialist-level accuracy on diabetic retinopathy (DR) referral, and Ting et al. [7], who addressed DR grading, glaucomatous optic neuropathy and age-related macular degeneration together in a multiethnic Singapore cohort. More recently Hu et al. [3] reported a multi-task framework spanning ophthalmic and neurodegenerative endpoints. Frozen-backbone evaluation of retinal foundation models for DR and glaucoma detection, in which a single linear layer is trained on fixed self-supervised features, was reported by Zhou et al. [8] alongside RETFound’s fine-tuned results, where a substantial gap between the two protocols was observed, and was later broadened by the same authors into a comparison of generalist and specialist backbones under both protocols [27].

That fundus photographs also encode systemic physiology – the field termed oculomics [1] – was shown at scale by Poplin et al. [2], who predicted age, sex, smoking status, systolic blood pressure and body mass index from the retinal images of 284,335 patients with clinically informative accuracy, establishing that the retinal vasculature carries multidimensional physiological state accessible to deep learning. Qian et al. [28] extended the analysis to UK Biobank [29], asking whether such estimated risk factors are statistically equivalent to direct clinical measurements for cardiovascular risk stratification.

Predicting incident events from a single baseline photograph is a more demanding axis than cross-sectional biomarker estimation, requiring features that capture subclinical pathology antedating clinical onset: vascular for myocardial infarction and ischaemic stroke, and for Parkinson’s disease the retinal ganglion cell and inner plexiform layer changes that reflect early neurodegeneration [3]. Systematic benchmarking of retinal foundation models on incident event prediction under frozen-backbone conditions has not been reported for any of these endpoints.

Taken together these tasks form a demanding multi-task benchmark for the generality of self-supervised retinal representations. Existing evaluations address subsets of them, mostly under fine-tuning conditions [8, 9, 27]; none reports a single frozen-linear evaluation spanning disease grading, biomarker regression and incident event prediction across both internal and external cohorts, which is the gap this work addresses.

## 3. Materials and Methods

### 3.1. RFM Pre-training Architecture and Self-Supervised Objective

RFM is initialised from a DINOv3-Base Vision Transformer [18] checkpoint pre-trained on natural images, with approximately 86 million parameters, 12 transformer layers, a patch size of 16× 16 pixels, and 768-dimensional token embeddings. For all downstream feature extraction the backbone is truncated after the ninth transformer layer (Section 3.2): the final three layers are discarded together with the projection network, the two conceptually forming a single projector that is used only during self-supervised training and plays no part at inference. This leaves an encoder of nine transformer layers and 64,378,368 parameters, 64 million and 25% smaller than the full DINOv3-Base checkpoint. We measured the resulting inference saving directly: op-level FLOP counting gives a 24.8% reduction in per-image computation, and wall-clock inference time falls by 24.4% on a single NVIDIA RTX 4090 at batch size 64 and 336 × 336 input resolution. Because the discarded blocks contribute nothing to the extracted feature, this saving carries no accuracy cost (Section 3.2). During self-supervised learning, the CLS token at the output of DINOv3 is fed into a randomly initialised three-layer projection network *g* (768→ 2048 →2048→ 256) with batch normalisation and ReLU activations. We further pre-train this DINOv3 checkpoint together with the projection network on CFPs using the LeJEPA objective to obtain RFM.

The DINOv3-Base checkpoint is itself a distillation of the much larger DINOv3 ViT-7B/16 model (approximately 7 billion parameters), which was pre-trained on LVD-1689M, a curated corpus of roughly 1.7 billion natural-world images [18]. RFM therefore inherits a general-purpose visual prior from natural images and adapts it to colour fundus photography, rather than being trained on retinal images from random initialisation; the DINOv3b-9L baseline (Section 3.4) is the same checkpoint without that retinal adaptation, which makes the RFM-versus-DINOv3b-9L contrast a direct measure of what the retinal LeJEPA stage contributes.

LeJEPA [12] combines a predictive loss that encourages consistency between representations of different augmented views of the same image with Sketched Isotropic Gaussian Regularisation (SIGReg), which encourages an isotropic Gaussian embedding distribution and so prevents representation collapse. It operates in representation space, without pixel-level reconstruction [26]. Figure 1 illustrates the RFM pre-training architecture.

**Figure 1.**
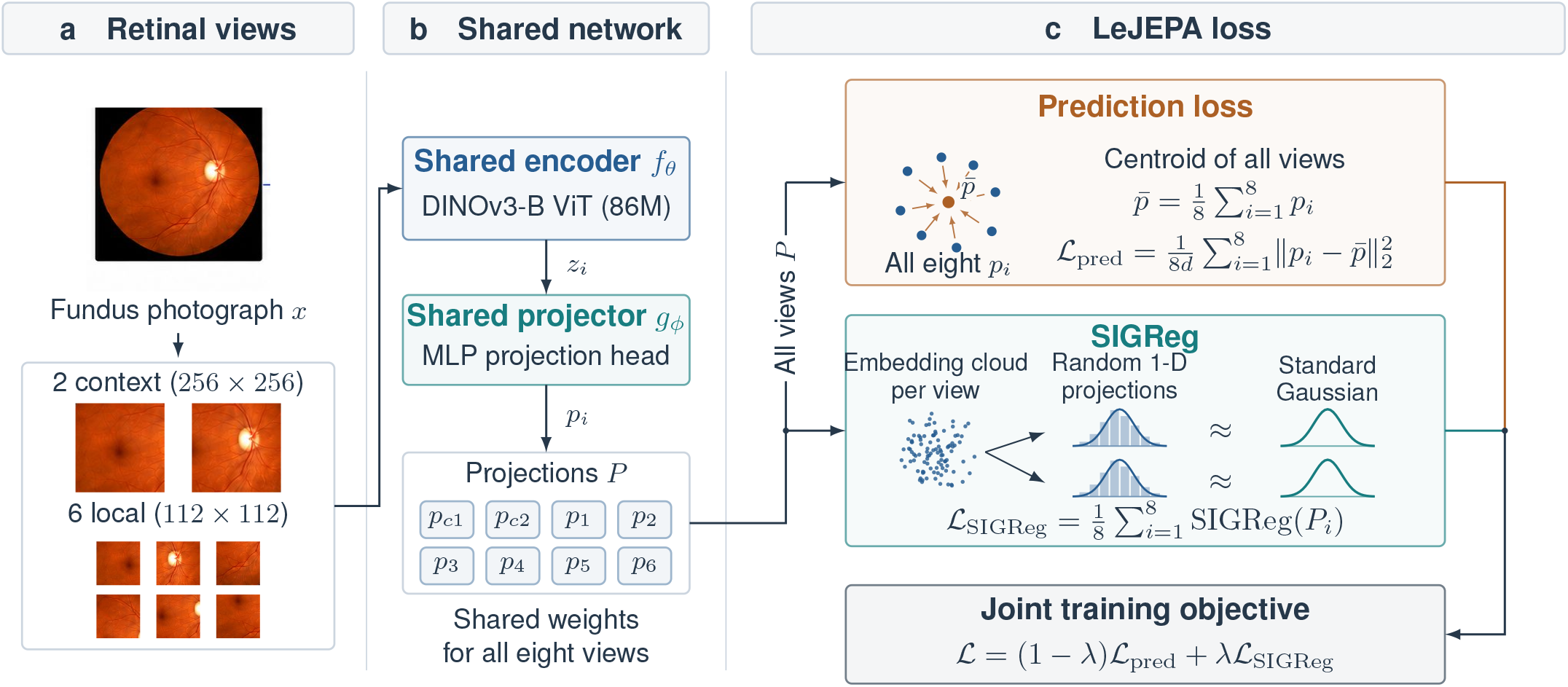
RFM LeJEPA pre-training objective. Two context views (256× 256) and six local views (112 ×112) are sampled from each retinal fundus image and encoded by the shared DINOv3-Base ViT *f*_*θ*_, and the resulting CLS embeddings *z*_*i*_ are mapped by the shared projector *g*_*ϕ*_ to the projected embeddings *p*_*i*_ on which both losses act. Every view is aligned to the view centre 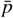, the mean of all eight projected view embeddings (Eq. (1)); a Sketched Isotropic Gaussian Regularisation term (Eq. (2)) matches random one-dimensional projections of each view’s embedding distribution across the mini-batch to a standard Gaussian, preventing representational collapse. The two losses are linearly combined into ℒ_LeJEPA_ (Eq. (3)).

Concretely, each fundus image is used to sample two context views (256 ×256 pixels) and six local views (112 ×112 pixels), for a total of *V* = 8 views per image. All eight views are passed through the shared DINOv3-Base encoder to produce CLS embeddings *z*_1_, …, *z*_*V*_ ∈ ℝ^768^, which are then mapped by the projector *g* to *p*_*i*_ = *g*(*z*_*i*_) ∈ ℝ^*d*^ with *d* = 256. Both LeJEPA losses are computed on the projected embeddings *p*_*i*_, and both are evaluated over a mini-batch of *N* images; we write *p*_*n,i*_ for view *i* of image *n* where the batch index matters. The projector is discarded after pre-training; the downstream representation is a distinct feature *h*, taken from the truncated ninth layer rather than from the layer-12 CLS embedding *z*_*i*_ used here (Section 3.2).

The view centre 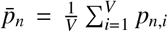 is the mean of all *V* projected view embeddings of image *n*, and every view is trained to align with it through the predictive loss

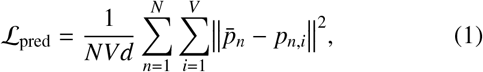

which averages the squared error over the *d* embedding dimensions as well as over views and images. Unlike LeJEPA, which centres on the global views only [12], we average all *V* views; during pre-training this was empirically more stable and gave slightly higher linear-probe accuracy on the DESP validation split, an observation we did not quantify in a controlled ablation. Because minimising ℒ_pred_ alone admits the trivial solution in which all embeddings collapse to a constant vector, LeJEPA couples it with the SIGReg term, applied to the same projected embeddings. SIGReg draws *S* random directions *a*_*s*_ ∈ ℝ^*d*^, ∥ *a*_*s*_ ∥ _2_ = 1, projects the embeddings onto each, and penalises the departure of every one-dimensional projection from a standard Gaussian using the characteristic-function (Epps–Pulley) goodness-of-fit statistic,

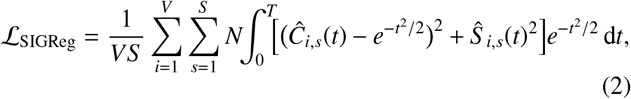

where *Ĉ*_*i,s*_ and *Ŝ*_*i,s*_ are the empirical cosine and sine transforms of the projected embeddings and *N* is the Epps–Pulley samplesize normalisation, so *λ* is defined relative to a given batch size. We compute the statistic per view slot *i*, over the *N* embeddings of that view in the mini-batch,^2^ rather than on the pooled set of *NV* embeddings, which constrains each view’s marginal distribution to isotropy individually. Balestriero and LeCun [12] prove that this term prevents representational collapse without negative memory queues, centring, sharpening, stop-gradient operations or a momentum-updated target encoder. The two terms are combined into the final training objective

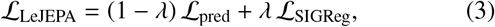

with *λ* balancing predictive alignment across views against representational isotropy.

We use *S* = 1024 sketched directions, resampled at every step, and evaluate the integral in Eq. (2) over *t* ∈ [0, 5] by trapezoidal quadrature on 17 equally spaced nodes. The balance is set to *λ* = 0.05, which weights the objective towards predictive alignment while retaining enough isotropy pressure to prevent collapse.

#### 3.1.1. Domain-specific pretext tasks

We propose pretext tasks designed for the circular geometry and pathological heterogeneity of CFP (Figure 2). Each image contributes two global crops and six local crops during each pre-training step. Global crops are drawn by random resized cropping from the full image area and resized to 256 ×256 pixels; local crops are sampled exclusively within the circular valid-pixel region and resized to 112 ×112. Both undergo flipping and rotation, global crops additionally Gaussian blurring and local crops colour jittering. Exact crop scales, rotation ranges and augmentation probabilities are listed in Supplementary Section 2 (Table S9). The restriction of local crops to the valid circular CFP area prevents sampling from the uninformative dark border regions common in fundus photography, ensuring that local augmentations consistently capture pathologically relevant retinal content. Global crops encourage learning of large-scale structural representations, such as optic disc morphology and major vessel architecture, while local crops are intended to promote feature learning at the finer spatial scale of microaneurysms, haemorrhages, and other fine-grained retinal pathology.

**Figure 2.**
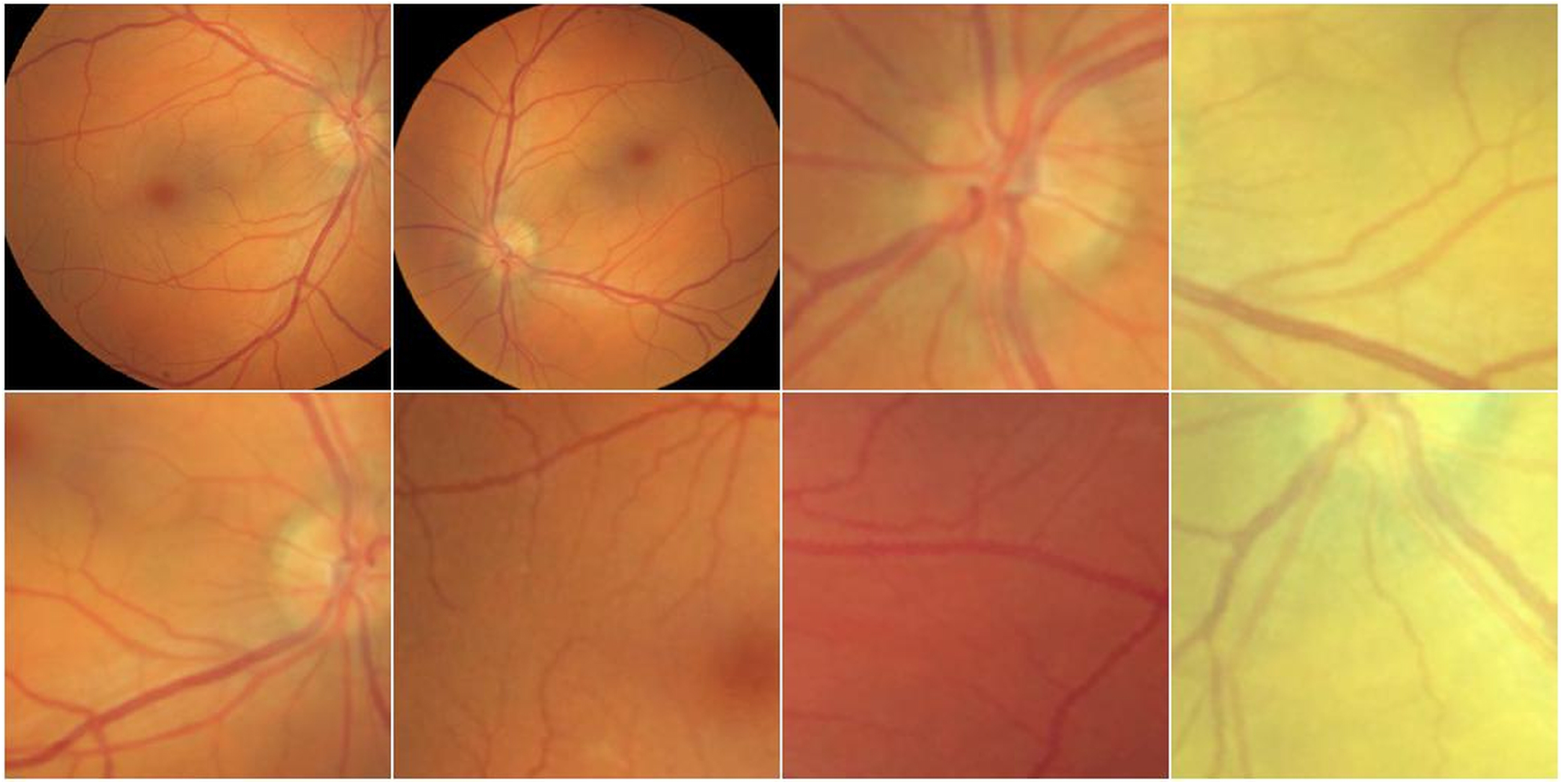
Domain-specific pretext tasks for RFM pre-training. Example shown on a public APTOS 2019 image for data-protection reasons (training used DESP). Two global crops (256 ×256, full image area) and six local crops (112 ×112, restricted to the valid circular CFP region) are generated per image. Local crops are constrained to the circular fundus area to prevent sampling uninformative border pixels.

#### 3.1.2. Training schedule and projector adaptation warmup

The model is optimised with AdamW (learning rate 10^−4^, weight decay 0.05) for 200 epochs with a batch size of 220 images per GPU on four NVIDIA A100 GPUs, giving a global batch of 880 images, with mixed-precision training and gradient-norm clipping. The learning rate schedule has three phases: a projector-only adaptation warmup over the first five epochs with the backbone frozen, a backbone warmup over the next five in which all parameters are updated, and cosine decay for the remainder. Complete hyperparameters are given in Supplementary Section 2 (Table S9).

The projector adaptation warmup was adopted on the basis of pilot experiments rather than of a controlled ablation. The motivating observation is that at the start of training the randomly initialised projector propagates high-variance gradients through the backbone, and pilot runs without this phase showed slow convergence and degraded final performance. Training the projector in isolation first, allowing it to adapt to the pre-existing DINOv3 latent space and bridge it to the LeJEPA prediction objective before backbone weights are updated, was intended to preserve the inherited representations through that early phase. We did not run the matched with-and-without comparison that would quantify its contribution (Section 5.5). Training was monitored by following the decrease and subsequent plateauing of the LeJEPA loss components.

#### 3.1.3. Pre-training dataset

RFM is pre-trained on the 4.3 million DESP images described in Section 3.3. All images are preprocessed by detecting the circular valid-pixel region, cropping to that region, and resizing to 1024 ×1024 pixels. Pixel intensities are normalised using the per-channel mean and standard deviation computed over the entire DESP dataset. No disease labels or demographic annotations are used during pre-training. The self-supervised stage draws on DESP images exclusively: neither UK Biobank nor any of the downstream evaluation cohorts (APTOS 2019, IDRiD, Messidor-2, GFID, PAPILA) enter self-supervised training, validation or model selection at any point. This concerns the self-supervised stage described here; the natural-image corpus used to pre-train the initialising DINOv3 checkpoint is a separate question, considered in Section 5.2.

### 3.2. Linear Head Training and Evaluation Protocol

All downstream evaluation uses a frozen RFM backbone with no gradient updates to backbone parameters, and the feature vec-tor for inference is taken from the output of the ninth transformer layer (Section 3.1). This configuration was chosen in preference to the full 12-layer output on the UK Biobank validation splits, as described below; the same contrast, measured end-to-end as DINOv3b-9L versus the twelve-layer DINOv3b, is reported in Section 5.2. Truncation is applied identically to RFM and to DINOv3b-9L, so the two share an inference cost as well as an architecture. The layer-9 CLS-plus-mean-patch configuration was selected once, on the UK Biobank *validation* splits (Section 3.3.2), using only the sex, age and HbA1c phenotypes; no incidence labels and no test split were used, and the configuration was subsequently frozen for all head training and evaluation; because splits are grouped at participant level (Section 3.3.2), no participant contributing to any reported evaluation was involved in it. Features are extracted by concatenating the classification token (CLS) representation (768-dimensional) with the mean-pooled patch token representations (768-dimensional) at layer 9 of DINOv3-Base, followed by layer normalisation, yielding a 768 + 768 = 1536-dimensional feature vector *h* per image. The linear head is trained on *h* for each downstream task. Figure 3 illustrates the inference pipeline.

**Figure 3.**
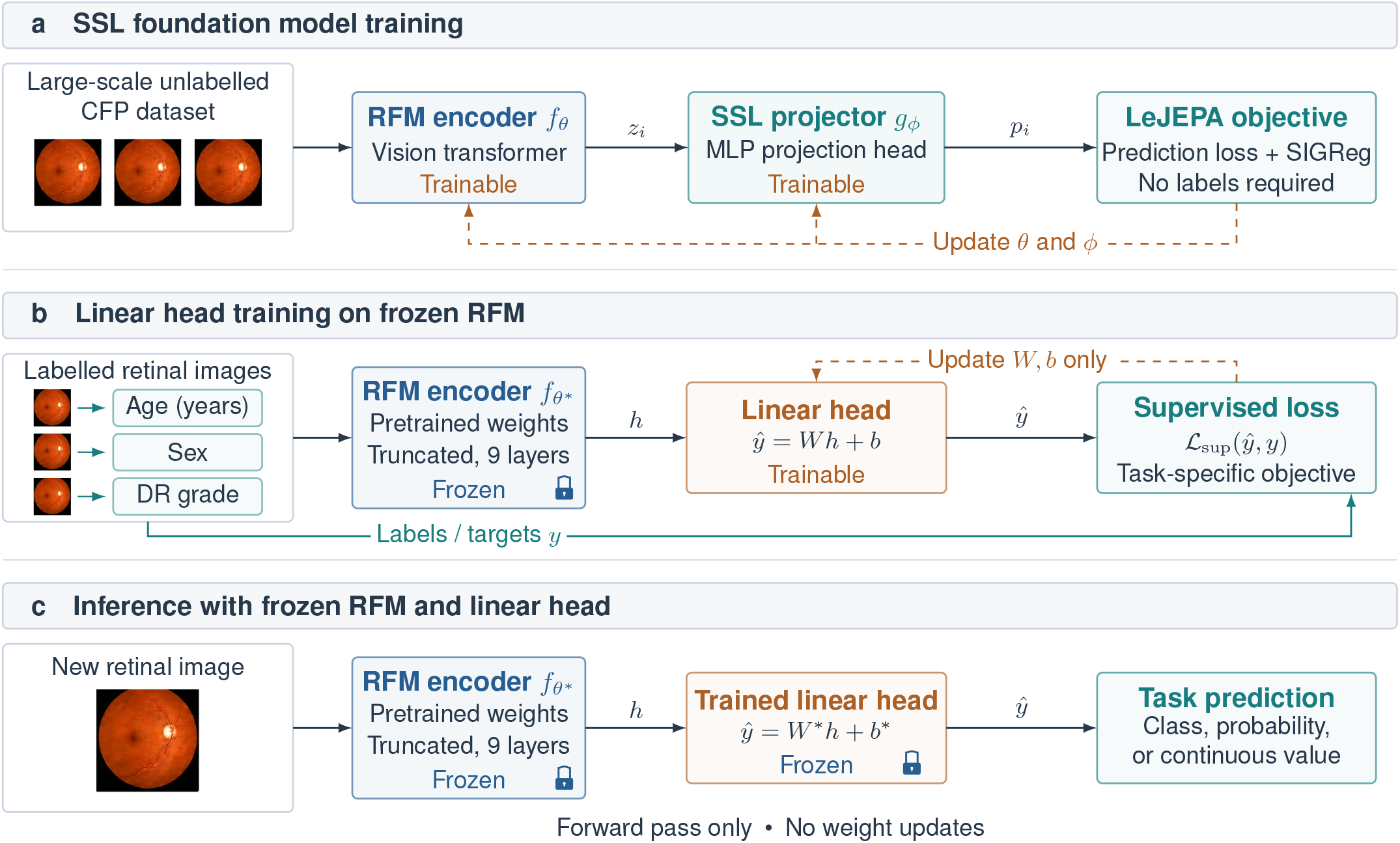
Pre-training, downstream linear head training, and inference. (a) Self-supervised pre-training: the 12-layer DINOv3-Base encoder and the projector are trained on 4.3 million unlabelled DESP CFP images under the LeJEPA objective, with no labels at any point. Layers 10–12 are treated as part of the SSL projector and discarded alongside it after pre-training, leaving a nine-layer truncated backbone *f*_*θ*_^*^ that is frozen in (b) and (c). (b) Linear head training on labelled images paired with task targets (e.g. age, sex, DR grade) and (c) inference: the RFM backbone is frozen throughout (no gradient updates) and produces the 1536-dimensional feature vector *h* from the concatenation of the classification token (CLS) and mean-pooled patch tokens at layer 9, followed by layer normalisation. A single linear head is trained per task and dataset on top of this frozen feature vector; at inference the same frozen backbone and trained linear head are applied to new images.

The linear head is a single affine layer with no non-linear activation, with output dimension one for continuous regression targets or *N* for *N*-class classification. Training-time augmentation, the evaluation transform and the optimiser settings are given in full in Supplementary Table S10; RFM heads are trained at 336 ×336, a resolution that marginally and consistently improved downstream performance over the 256 ×256 used during SSL pre-training, and the reported checkpoint is the epoch with the best validation score. Every model is frozen and probed with this same single-affine-head protocol, on identical splits and with the same augmentation pipeline. For the three RETFound variants the resolution, feature-extraction convention and input normalisation follow each backbone’s own published linear-probe recipe, so that no comparator is penalised by a convention it was not designed for; the optimiser settings also follow that recipe for RETFound-MAE and RETFound-DINOv2, while RETFound-Green’s head is fitted with the shared optimiser and schedule (Section 3.4.4). DINOv3b-9L and DINOv3b are the exception, and deliberately so: being the checkpoint from which RFM is initialised, they are given RFM’s resolution and DESP normalisation statistics rather than their own natural-image conventions, and DINOv3b-9L also RFM’s feature extraction, which is what makes the RFM-versus-DINOv3b-9L contrast single-variable (Section 3.4). Exact augmentation parameters, the shared protocol and the per-backbone settings are listed in Supplementary Section 2 (Tables S10 and S11). One independent head is trained per task and dataset. Classification heads are optimised with a cross-entropy loss (CrossEntropyLoss); regression heads are optimised with an L1 loss (L1Loss), directly matching the Mean Absolute Error (MAE) metric reported for regression tasks.

### 3.3 Datasets

#### 3.3.1. Pre-training and internal incidence dataset

The DESP dataset [13] comprises colour fundus photographs from a national diabetic eye screening programme in North East London. A detailed characterisation of the DESP cohort, including demographic composition, diabetic retinopathy grading distribution, and linked electronic health record coverage, is provided in a dedicated cohort profile [30]. Raw screening captures include non-retinal and low-quality images; these were excluded with the QUARTZ tool [31] to yield the 4.3 million retinal CFP images used for pre-training (Supplementary Section 1.2). A subset of participants with linked electronic health records is used for training and evaluating linear heads for 3-year incidence prediction of myocardial infarction, ischaemic stroke, and Parkinson’s disease, with outcomes derived from linked primary and secondary care records. For the 3-year incidence tasks, only high-quality DESP images without diabetic retinopathy or macular degeneration were used: DR/AMD-free images were identified from the DESP grading, and image quality was separately assessed using the QUARTZ tool [31].

#### 3.3.2. Data splits

All training, validation and test splits used in this work were generated by us rather than adopted from prior benchmarks, under one uniform protocol: grouping at the patient level wherever subject identifiers are published, class-stratified allocation, and label assignment verified against each dataset’s own released metadata. Patient-level grouping assigns every image of an individual to one split, so fellow eyes and repeat visits can never be divided between training and test. PAPILA and Messidor-2 publish the identifiers this requires; GFID, APTOS 2019 and IDRiD do not, so those three are partitioned at the image level and rely on the independence of images asserted by the source distributions, with the residual risk that entails (Supplementary Section 1.1). Applying one protocol throughout is what makes the comparisons here internally consistent, since differences between models cannot then be confounded by differences in how each benchmark was partitioned; because the splits necessarily differ from those used in some previously published evaluations, all comparator models were re-run on them (Section 3.4.3). Split definitions are released with the source code (Data Availability Statement).

All training, evaluation and reported counts are at the image level, and a participant may contribute several images to their own split, so counts in this paper and in the Supplementary Material are image counts unless a participant count is stated explicitly. Per-split counts and class distributions for every cohort are given in Supplementary Tables S1–S8.

#### 3.3.3. 3-year incidence cohort definition and evaluation protocol

Three-year incident outcomes were defined relative to the baseline retinal imaging date. Patients with a recorded history of the target condition at or before baseline were excluded. Positive cases were defined as patients who experienced the target event within three years after baseline, including those who subsequently died during this period. Negative cases were required to have at least three years of follow-up without the target event. Patients with shorter event-free follow-up, including those who died before three years without experiencing the target event, were excluded.

We followed the RETFound 3-year incidence evaluation protocol [8] in two further respects, so that our figures are constructed on the same basis as the published figures we compare against.

First, we constructed *balanced case–control subsets*, retaining all eligible incident events and drawing an equal number of controls at random, giving a 1:1 event-to-control ratio in every training and test set (Section 4.4.1). Second, the incidence sets were restricted to a single left-eye image per participant, taken at one visit: no participant contributed multiple images or repeated visits to these sets. Per-endpoint event and control counts for the internal DESP and external UK Biobank sets are given in Supplementary Table S3.

#### 3.3.4. UK Biobank

The UK Biobank (UKBB) [17, 29] is a large population-based prospective cohort study; its recruitment and ocular imaging protocol are summarised in Supplementary Section 1.2. Its fundus images are used here in two roles: to train and evaluate linear heads for the ten biomarker and demographic phenotypes of Section 4.1, and as an external validation cohort for the 3-year incidence heads trained on DESP, an out-of-cohort transfer assessment in which no head is retrained and no UK Biobank incidence label is used (Section 4.4.3).

In accordance with UK Biobank’s policy on reporting small numbers,^3^ confusion matrix cells derived from UK Biobank data that represent fewer than five participants are reported as “*<*5” rather than as exact counts.

#### 3.3.5. Public ophthalmic benchmarks

Five public datasets are used. DR grading: APTOS 2019 (3,662 images, India) [32], IDRiD (516 images, India) [33] and Messidor-2 (1,744 images from 874 patients, France) [34, 35, 36], all graded on the five-class International Clinical Diabetic Retinopathy scale. Glaucoma: GFID (1,544 optic-nerve-head crops, South Korea) [15] and PAPILA (488 images with paired eyes, Spain) [16], both evaluated as three-class grading tasks under their own taxonomies, no glaucoma / early / advanced for GFID and healthy / glaucoma / suspicious for PAPILA, the last a borderline group of uncertain diagnosis. Acquisition details, camera models, resolutions and licensing are given in Supplementary Section 1.2.

#### 3.3.6. MEH-MIDAS and AlzEye datasets

MEH-MIDAS [8] and AlzEye [37] are distinct Moorfields Eye Hospital-derived datasets serving different roles here: MEH-MIDAS is part of RETFound’s pre-training corpus (Section 2), while AlzEye is the cohort on which the published RETFound incidence heads were fine-tuned and internally evaluated. Neither is used for RFM at any stage.

### 3.4. Baselines, Comparators, and Evaluation Metrics

We compare RFM against five baselines evaluated under the same frozen-linear protocol, each with its own published featureextraction and linear-probe recipe (Supplementary Table S11):

- **RETFound-MAE**^4^ **and RETFound-DINOv2** [8, 27]: Both variants use a ViT-Large backbone (approximately 307M parameters, 1024-dimensional embedding) pre-trained on the curated MEH-MIDAS and Kaggle EyePACS datasets (Section 3.3.6). RETFound-MAE is pre-trained with a Masked Autoencoder, while RETFound-DINOv2 is pre-trained with a DINOv2-based objective. Both were run by us (Section 3.4.3) under their published feature conventions: the final-layer CLS token for RETFound-DINOv2 and mean-pooled final-layer patch tokens for RETFound-MAE. Only their 3-year incidence figures are taken from the literature.
- **DINOv3b-9L** [18]: The same DINOv3-Base checkpoint from which RFM is initialised, without any retinal domain adaptation. Included as a natural-image generalist baseline to isolate the contribution of the retinal LeJEPA stage. It is truncated after the ninth transformer layer and its features extracted with the same CLS-plus-mean-patch protocol used for RFM (Section 3.2), so RFM and DINOv3b-9L are identical in architecture, inference cost and feature extraction, and differ only in pre-training data and objective.
- **DINOv3b** [18]: The same checkpoint as released, all twelve transformer layers, read by the common global average pooling of the final-layer patch tokens in place of the ninth-layer CLS-plus-mean-patch features of DINOv3b-9L. DINOv3b is trained with the same hyperparameters as DINOv3b-9L (Supplementary Table S11), so the two differ only in feature extraction (Section 5.2).
- **RETFound-Green** [9]: A parameter-efficient RETFound variant based on DINOv2-S (22M parameters, 384-dimensional embedding), trained on 75,000 publicly available retinal images (AIROGS, DDR, and ODIR-2019) with 400× less compute than RETFound-MAE. It too was run by us rather than read from the published figures; one deviation from its original evaluation is noted in Section 3.4.4.

#### 3.4.1. Evaluation metrics

Regression tasks are evaluated using mean absolute error (MAE), relative MAE improvement over a mean-predictor baseline, and Spearman rank correlation *ρ*. Classification tasks are evaluated using the area under the receiver operating characteristic curve (AUROC), the area under the precision–recall curve (AUPRC), and F1 score. DR grading is five-class, glaucoma grading three-class and ethnicity four-class; for these, all three metrics are computed one-vs-rest and macro-averaged over classes, F1 over the argmax prediction. For the binary tasks (sex, smoking status and the 3-year incidence endpoints) AUROC and AUPRC are computed on the positive-class score.

F1 requires a decision threshold, which is 0.5 for sex and smoking status and, for the 3-year incidence endpoints, 0.2 (MI) and 0.3 (stroke and Parkinson’s disease). F1 for the incidence endpoints is reported only in the supplementary per-draw tables (Supplementary Tables S14–S16); because those thresholds were chosen per endpoint while the published RETFound figures use their authors’ own operating points, it is not a threshold-matched comparison and is not used for model comparison.

#### 3.4.2. Confidence intervals and the resampling unit

All metrics are reported with 95% confidence intervals obtained by bootstrap resampling with 4000 replicates. Where a dataset provides subject identifiers the resampling unit is the participant: participants are drawn with replacement and all of their images are carried together, so the interval reflects the number of independent individuals rather than the number of images. This applies to UK Biobank, PAPILA and Messidor-2, in which a participant contributes on average 1.78, 2.00 and 2.00 test images respectively. GFID, APTOS 2019 and IDRiD publish no subject identifiers (Section 3.3.2), so the image is the only available resampling unit for those three. The metrics themselves are computed per image throughout, because the eye rather than the patient is the unit of disease: fellow eyes can carry different diabetic retinopathy or glaucoma grades. For the 3-year incidence endpoints each participant contributes exactly one left-eye image from a single visit (Section 3.3.3), so the image, the eye and the participant are the same unit and no clustering correction applies.

#### 3.4.3. Provenance of comparator results

Every frozen-linear comparator value in this paper was computed by us, under the protocol of Section 3.2 and on the splits of Section 3.3.2, with a single exception. Every model was run inhouse on all five ophthalmic benchmarks and all ten UK Biobank phenotypes, so the extraction-convention ablation of Section 5.2 spans both; only the 3-year incidence endpoints lie outside it, neither DINOv3b-9L nor DINOv3b having been run on those. These comparisons are therefore matched on head architecture, optimiser family, training and evaluation data splits and augmentation pipeline; each RETFound variant additionally carries its own published input resolution, feature-extraction convention, input normalisation and linear-probe schedule (Supplementary Table S11), so that no comparator is evaluated outside the regime it was released for. DINOv3b-9L and DINOv3b instead carry RFM’s settings throughout (Section 3.2). Training and evaluation code for the diabetic retinopathy and glaucoma experiments, including how each RETFound variant was instantiated, is available in the project repository (Data Availability Statement).

The exception is the 3-year incidence endpoints, for which neither RETFound variant was re-computed: RETFound-DINOv2 figures were read from Supplementary Figure 10 of Zhou et al. [27], and RETFound-MAE figures were taken from the RET-Found publication [8], which reports both internal AlzEye and external UK Biobank results. Those rows, and only those rows,are marked with a dagger (^†^) in the results tables; every un-marked row above the rule is our own run, and the published end-to-end results below the rule (Section 3.4.5) are quoted from their publications for context. Every comparison in this paper except those rows is computed on one test set shared by all models (Section 3.4.6).

#### 3.4.4. Linear head used for RETFound-Green

The original RETFound-Green evaluation fits its linear probe with scikit-learn, standardising features on the training split and then fitting a logistic regression to convergence. We instead trained RETFound-Green’s head with the same single affine layer, AdamW optimiser and 200-epoch cosine schedule used for RFM, DINOv3b-9L and DINOv3b (Section 3.2), while keeping its own published input resolution, normalisation statistics and pooled post-norm feature. This removes the head-fitting procedure as a confound between RETFound-Green and RFM.

#### 3.4.5. Published end-to-end comparators

As a frame of reference, published results from state-of-the-art end-to-end trained models are also included in the UK Biobank biomarker and classification tables: Poplin et al. [2], Rim et al. [38], Squirrell et al. [39], White et al. [40], Zhu et al. [41], Korot et al. [42], and Hu et al. [3] (FundusNet). These methods typically use full end-to-end fine-tuning, larger or task-specific architectures and, in some cases, different cohorts, so they are included for context rather than comparison (Section 4).

#### 3.4.6. Comparability of the incidence comparisons

RETFound heads for MI, stroke, and Parkinson’s incidence are reported as trained on the AlzEye dataset, while RFM heads in this work are trained on DESP. As these datasets differ in patient population, demographics, imaging protocols, and outcome prevalence, the DESP/AlzEye tables (Section 4.4) report two measurements made on different cohorts rather than a comparison, and should be read as separate per-dataset evaluations. In the external UKBB evaluation (Section 4.4.3) both models’ heads are applied to an unseen cohort without retraining. We did not re-run RETFound on our own UK Biobank subset: the RETFound values are those reported by Zhou et al. [27] and by the RETFound publication [8], computed on their own UK Biobank subsets under their own inclusion criteria. Both follow the same balanced case–control construction (Section 3.3.3), so the metrics are constructed on a common basis, but they are computed on different participants.

## 4. Results

This section reports frozen-backbone linear-head performance across four task groups, one subsection each, with one table per phenotype or dataset: UK Biobank (UKBB) biomarkers, diabetic retinopathy (DR) grading, glaucoma detection, and 3-year incidence of myocardial infarction (MI), ischaemic stroke and Parkinson’s disease on internal (DESP for RFM, AlzEye for RETFound) and external (UKBB) cohorts. Tables report the metrics defined in Section 3.4.1, with 95% confidence intervals in parentheses where available; the 3-year incidence tables omit *F*_1_ for the reason given there. In every table, bold marks the best and underline the second-best value in each column among the frozen-linear models, and a dagger (^†^) marks a value reported by the cited publication rather than reproduced by us. Two conventions are worth stating once: the published end-to-end results below the rule in each table are shown for context and excluded from the ranking, and the internal incidence tables carry no ranking at all, their rows being measurements on different cohorts (Section 3.4.6). The Supplementary Material carries the per-split counts and class distributions (Tables S1–S8), the LDL and HDL tables (Tables S12–S13), the predicted-vs-true density plots for all seven regression biomarkers (Figures S1–S7), the precision–recall curves for the classification tasks whose main-text figures show only ROC curves and confusion matrices, and the five individual control draws behind each external incidence result (Tables S14–S16, Figures S8–S13). The external incidence evaluation is the one place where both ROC and precision–recall curves appear in the main text (Section 4.4.3). Figure 4 summarises frozen-linear performance for RFM, the best-performing RETFound variant per task, and RETFound-Green across all 15 regression, UKBB classification, DR, and glaucoma tasks reported below, before the per-phenotype tables that follow.

**Figure 4.**
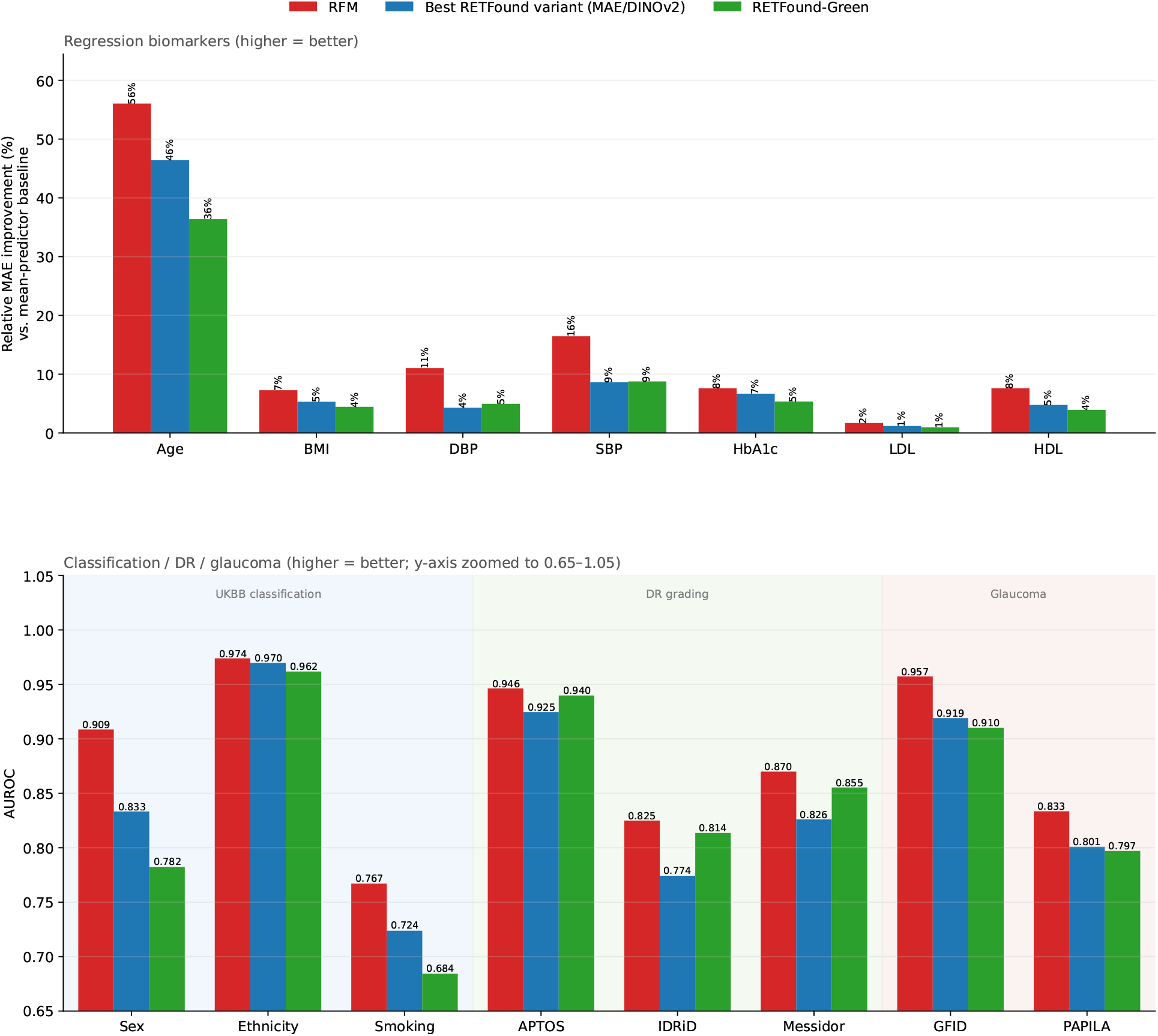
Frozen-linear performance of RFM, the best-performing RETFound variant per task (MAE or DINOv2), and RETFound-Green across all 15 regression, UK Biobank classification, diabetic retinopathy, and glaucoma tasks reported in this work. Top: relative MAE improvement over a mean-predictor baseline (regression biomarkers). Bottom: AUROC (classification/DR/glaucoma tasks; y-axis zoomed to 0.65–1.05 to make margins visible). 3-year incidence tasks are excluded (evaluated only for RFM and RETFound, on different cohorts; Section 4.4).

### 4.1. UKBB Biomarker Prediction

#### 4.1.1. Regression biomarkers

Seven continuous UKBB biomarkers are evaluated: age, BMI, diastolic blood pressure (DBP), systolic blood pressure (SBP), LDL cholesterol, HDL cholesterol, and HbA1c. Tables 1–5 report frozen-linear prediction performance for five of these (age, BMI, DBP, SBP, and HbA1c); LDL and HDL cholesterol results are reported in the Supplementary Material (Tables S12 and S13) for space, and summarised within the narrative below. Age is RFM’s strongest regression result (Table 1). RFM achieves a MAE of 3.09 years (95% CI: 3.06–3.12) and a Spearman correlation of *ρ* = 0.870 (0.867–0.873), representing a 56.0% relative MAE improvement over the mean-predictor baseline. RETFound-MAE achieves a MAE of 3.76 (*ρ* = 0.815), a gap of 0.67 years in absolute MAE (21.7% higher error). DINOv3b-9L, with no retinal pre-training, yields a MAE of 3.95 (*ρ* = 0.784), indicating that retinal-domain SSL improves age encoding. Among all published methods, the highest age prediction accuracy is reported by Hu et al. [3], whose FundusNet attains MAE = 2.55; that result uses full fine-tuning with a non-linear head, whereas RFM is the best performer under the frozen-linear constraint.

**Table 1.**
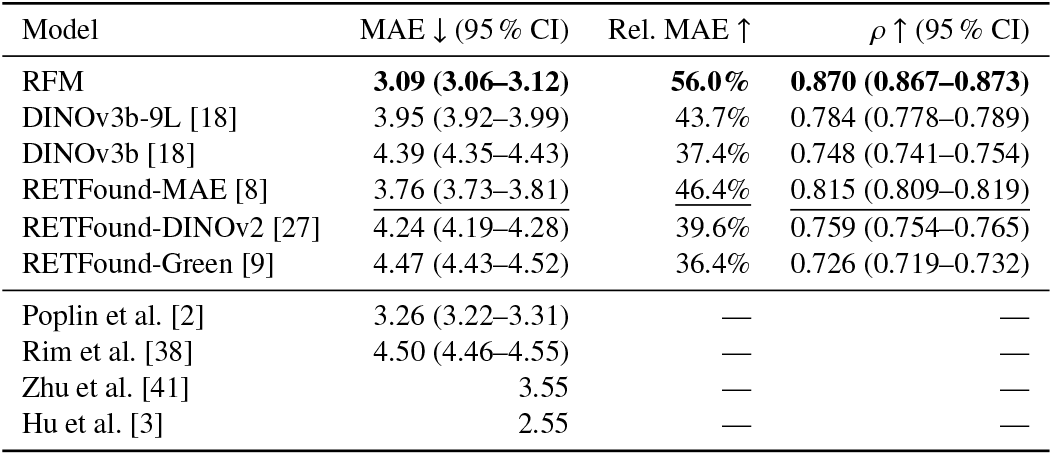
Age (years) prediction from frozen retinal embeddings on UK Biobank [17]. Bold/underline rank the frozen-linear models only; the published end-to-end results below the rule use different protocols and are shown for context, not ranked.

| Model | MAE ↓ (95 % CI) | Rel. MAE ↑ | $\rho$ ↑ (95 % CI) |
| --- | --- | --- | --- |
| RFM | **3.09 (3.06–3.12)** | **56.0%** | **0.870 (0.867–0.873)** |
| DINOV3b-9L [18] | 3.95 (3.92–3.99) | 43.7% | 0.784 (0.778–0.789) |
| DINOV3b [18] | 4.39 (4.35–4.43) | 37.4% | 0.748 (0.741–0.754) |
| RETFound-MAE [8] | 3.76 (3.73–3.81) | 46.4% | 0.815 (0.809–0.819) |
| RETFound-DINOV2 [27] | 4.24 (4.19–4.28) | 39.6% | 0.759 (0.754–0.765) |
| RETFound-Green [9] | 4.47 (4.43–4.52) | 36.4% | 0.726 (0.719–0.732) |
| Poplin et al. [2] | 3.26 (3.22–3.31) | — | — |
| Rim et al. [38] | 4.50 (4.46–4.55) | — | — |
| Zhu et al. [41] | 3.55 | — | — |
| Hu et al. [3] | 2.55 | — | — |

**Table 2.** BMI (kg/m^2^) prediction from frozen retinal embeddings on UK Biobank. Bold/underline rank the frozen-linear models only; the published end-to-end results below the rule use different protocols and are shown for context, not ranked.

| Model | MAE ↓ (95 % CI) | Rel. MAE ↑ | $\rho$ ↑ (95 % CI) |
| --- | --- | --- | --- |
| RFM | **3.32 (3.28–3.36)** | **7.3%** | **0.315 (0.304–0.328)** |
| DINOV3b-9L [18] | 3.40 (3.37–3.45) | 4.8% | 0.238 (0.227–0.250) |
| DINOV3b [18] | 3.56 (3.52–3.61) | 0.6% | 0.193 (0.182–0.205) |
| RETFound-MAE [8] | 3.39 (3.35–3.43) | 5.3% | 0.270 (0.258–0.282) |
| RETFound-DINOV2 [27] | 3.41 (3.37–3.45) | 4.8% | 0.248 (0.236–0.260) |
| RETFound-Green [9] | 3.42 (3.38–3.46) | 4.4% | 0.219 (0.207–0.232) |
| Poplin et al. [2] | 3.29 (3.24–3.34) | — | — |
| Rim et al. [38] | 3.47 (3.44–3.51) | — | — |

**Table 3.** Diastolic blood pressure (mmHg) prediction from frozen retinal embeddings on UK Biobank. Bold/underline rank the frozen-linear models only; the published end-to-end results below the rule use different protocols and are shown for context, not ranked.

| Model | MAE ↓ (95 % CI) | Rel. MAE ↑ | $\rho$ ↑ (95 % CI) |
| --- | --- | --- | --- |
| RFM | **7.06 (7.00–7.13)** | **11.0%** | **0.447 (0.436–0.457)** |
| DINOV3b-9L [18] | 7.50 (7.44–7.58) | 5.5% | 0.336 (0.324–0.347) |
| DINOV3b [18] | 8.05 (7.99–8.15) | −1.5% | 0.302 (0.290–0.314) |
| RETFound-MAE [8] | 7.59 (7.53–7.68) | 4.3% | 0.313 (0.302–0.326) |
| RETFound-DINOV2 [27] | 8.01 (7.94–8.09) | −1.0% | 0.246 (0.235–0.259) |
| RETFound-Green [9] | 7.54 (7.48–7.62) | 5.0% | 0.311 (0.300–0.322) |
| Poplin et al. [2] | 6.42 (6.33–6.52) | — | — |
| Rim et al. [38] | 7.67 (7.59–7.74) | — | — |
| White et al. [40] | 6.54 | — | — |

**Table 4.** Systolic blood pressure (mmHg) prediction from frozen retinal embeddings on UK Biobank. Bold/underline rank the frozen-linear models only; the published end-to-end results below the rule use different protocols and are shown for context, not ranked.

| Model | MAE ↓ (95 % CI) | Rel. MAE ↑ | $\rho$ ↑ (95 % CI) |
| --- | --- | --- | --- |
| RFM | **12.11 (12.00–12.26)** | **16.5%** | **0.534 (0.525–0.543)** |
| DINOV3b-9L [18] | 13.07 (12.96–13.25) | 9.8% | 0.421 (0.410–0.431) |
| DINOV3b [18] | 13.83 (13.72–14.03) | 4.6% | 0.395 (0.384–0.406) |
| RETFound-MAE [8] | 13.24 (13.13–13.41) | 8.6% | 0.407 (0.396–0.418) |
| RETFound-DINOV2 [27] | 14.62 (14.49–14.78) | −0.9% | 0.279 (0.267–0.293) |
| RETFound-Green [9] | 13.22 (13.11–13.39) | 8.8% | 0.396 (0.384–0.407) |
| Poplin et al. [2] | 11.35 (11.18–11.51) | — | — |
| Rim et al. [38] | 13.57 (13.44–13.70) | — | — |
| Squirrell et al. [39] | 12.35 | — | — |
| White et al. [40] | 11.41 | — | — |

**Table 5.** HbA1c (mmol/mol) prediction from frozen retinal embeddings on UK Biobank. Bold/underline rank the frozen-linear models only; the published end-to-end results below the rule use different protocols and are shown for context, not ranked.

| Model | MAE ↓ (95 % CI) | Rel. MAE ↑ | $\rho$ ↑ (95 % CI) |
| --- | --- | --- | --- |
| RFM | **3.45 (3.39–3.52)** | **7.6%** | **0.347 (0.335–0.359)** |
| DINOV3b-9L [18] | 3.50 (3.45–3.58) | 6.0% | 0.302 (0.290–0.314) |
| DINOV3b [18] | 3.73 (3.67–3.81) | 0.0% | 0.272 (0.259–0.284) |
| RETFound-MAE [8] | 3.48 (3.42–3.56) | 6.7% | 0.324 (0.312–0.336) |
| RETFound-DINOV2 [27] | 3.56 (3.50–3.64) | 4.5% | 0.252 (0.239–0.265) |
| RETFound-Green [9] | 3.53 (3.47–3.61) | 5.4% | 0.270 (0.258–0.283) |
| White et al. [40] | 3.72 | — | — |

BMI prediction (Table 2) shows a smaller advantage for RFM than age: RFM achieves a MAE of 3.32 kg/m^2^ (*ρ* = 0.315), a 7.3% relative improvement over the mean-predictor baseline, ahead of RETFound-MAE (MAE 3.39, *ρ* = 0.270) and DINOv3b-9L (MAE 3.40, *ρ* = 0.238); Poplin et al. [2] report a marginally lower absolute MAE (3.29) under full fine-tuning.

For blood pressure prediction, RFM leads all foundation models on both DBP (MAE 7.06, *ρ* = 0.447) and SBP (MAE 12.11,*ρ* = 0.534); DINOv3b-9L is the second-best model on both endpoints (DBP *ρ* = 0.336; SBP *ρ* = 0.421), ahead of all three RETFound variants despite no retinal-domain pre-training. For HDL (Supplementary Table S13), RFM achieves *ρ* = 0.373 compared with *ρ* = 0.311 for RETFound-MAE, *ρ* = 0.294 for RETFound-DINOv2, and *ρ* = 0.290 for DINOv3b-9L. LDL prediction (Supplementary Table S12) yields uniformly weak Spearman correlations, RFM highest at *ρ* = 0.130 and every other model at *ρ* ≤0.100, suggesting that LDL is the least linearly accessible of the evaluated biomarkers from frozen retinal features.

HbA1c prediction shows a small but consistent advantage for RFM (MAE 3.45, *ρ* = 0.347) over RETFound-MAE (MAE 3.48, *ρ* = 0.324), DINOv3b-9L (MAE 3.50, *ρ* = 0.302), and RETFound-DINOv2 (MAE 3.56, *ρ* = 0.252). RFM’s advantage over RETFound-MAE is small and their 95% confidence intervals for *ρ* are closely overlapping; the Spearman correlations across all models (*ρ* ≈ 0.25–0.35) indicate that glycaemic state is partially, but not yet clinically actionably, encoded in frozen retinal features from a single fundus image.

The predicted-vs-true density plots of RFM and the three RET-Found variants (Supplementary Figures S1–S7) mirror these results. For age, RFM’s predictions follow the identity line across the age range, whereas the RETFound variants’ predictions are more dispersed and flatter at the extremes, most visibly for RETFound-Green. For every other biomarker all four models compress their predictions towards the cohort mean, underestimating high values and overestimating low ones; RFM’s prediction distribution is the most inclined towards the identity line for blood pressure and HDL, consistent with its higher *ρ*, while for LDL all four are close to flat.

#### 4.1.2. Classification phenotypes

Tables 6–8 report frozen-linear classification performance for sex, ethnicity, and smoking status. Per-split image counts and category distributions for all three phenotypes are given in Supplementary Table S1.

**Table 6.** Sex classification from frozen retinal embeddings on UK Biobank. AUROC, AUPRC, and F1 with 95 % CI. Bold/underline rank the frozen-linear models only; the published end-to-end results below the rule use different protocols and are shown for context, not ranked.

| Model | AUROC ↑ (95 % CI) | AUPRC ↑ (95 % CI) | F1 ↑ (95 % CI) |
| --- | --- | --- | --- |
| RFM | **0.909 (0.905–0.912)** | **0.922 (0.918–0.926)** | **0.851 (0.846–0.856)** |
| DINOV3b-9L [18] | 0.808 (0.802–0.813) | 0.832 (0.825–0.839) | 0.764 (0.758–0.769) |
| DINOV3b [18] | 0.654 (0.648–0.659) | 0.660 (0.653–0.668) | 0.735 (0.730–0.740) |
| RETFound-MAE [8] | 0.833 (0.828–0.838) | 0.854 (0.848–0.859) | 0.796 (0.791–0.801) |
| RETFound-DINOV2 [27] | 0.820 (0.814–0.825) | 0.844 (0.837–0.850) | 0.783 (0.778–0.788) |
| RETFound-Green [9] | 0.782 (0.776–0.788) | 0.808 (0.800–0.815) | 0.747 (0.742–0.753) |
| Poplin et al. [2] | 0.97 (0.966–0.971) | — | — |
| Rim et al. [38] | 0.80 (0.79–0.80) | — | — |
| Korot et al. [42] | 0.93 | — | — |
| Hu et al. [3] | 0.98 | — | — |

The smoking task is defined as never-smoker versus currentsmoker. The remaining responses (“occasionally”, “previous”, and “prefer not to say”/missing) are excluded because they correspond to neither state: “occasionally” does not specify a frequency and “previous” does not specify how long ago smoking ceased. Together these excluded categories account for 37% of the cohort (Supplementary Table S1), so the smoking results should be read as discriminating established current smoking from lifelong abstention, not as a general smoking-status classifier. For the same reason we excluded the “Unknown/didn’t answer” category from the ethnicity task.

RFM achieves AUROC of 0.909 for sex, 0.974 for ethnicity, and 0.767 for smoking status, leading all compared foundation models across all three phenotypes. For sex classification, DINOv3b-9L (AUROC 0.808) trails RETFound-MAE (0.833) and RETFound-DINOv2 (0.820) but is ahead of RETFound-Green (0.782); methods using full end-to-end fine-tuning report higher AUROC still, e.g. 0.97 for Poplin et al. [2] and 0.98 for Hu et al. [3], again confirming the gap between frozen-linear and fine-tuning evaluation conditions. To our knowledge, no prior foundation model AUROC for ethnicity has been reported in the existing literature. Consequently, RFM’s result (0.974; Figure 5) serves as the new reference point for this task, leading RETFound-MAE (0.970), DINOv3b-9L (0.966), RETFound-DINOv2 (0.965), and RETFound-Green (0.962). For smoking, RFM (AUROC 0.767) surpasses RETFound-MAE (0.724), DINOv3b-9L (0.707), RETFound-DINOv2 (0.699), RETFound-Green (0.684), and prior published methods, 0.71 for Poplin et al. [2] and 0.64 for Korot et al. [42].

**Figure 5.**
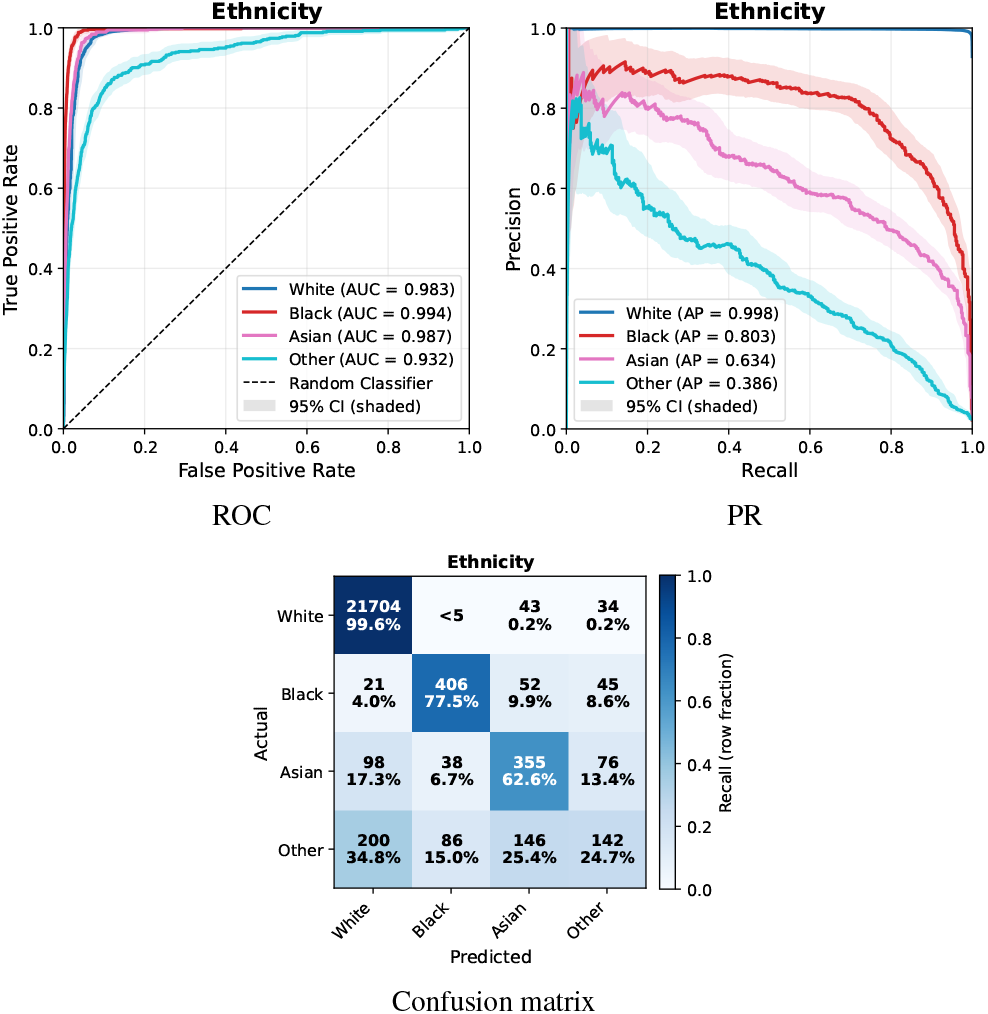
Classification diagnostics for ethnicity classification on UK Biobank (RFM only). Top row: ROC and precision–recall curves, with shaded 95% confidence intervals where available. Bottom: confusion matrix of the argmax prediction (rows: true class; columns: predicted class).

We note that the F1 column of Table 8 is low for every model, at most 0.112 (RFM). F1 is computed at the 0.5 threshold, and current smokers make up 9.3% of the never-versus-current test cohort (Supplementary Table S1), so at that threshold all models predict the negative class almost everywhere. These values therefore establish low sensitivity at a single arbitrary operating point; they do not on their own separate how much of that is attributable to the threshold and how much to the representations. F1 at a fixed threshold is not a meaningful basis for comparing models on this task; AUROC and AUPRC, which are computed over the full range of thresholds, are the informative measures here, and a deployment would select its operating point on a validation set rather than use 0.5.

**Table 7.** Ethnicity classification from frozen retinal embeddings on UK Biobank. No published end-to-end results for this phenotype. AUROC, AUPRC, and F1 with 95 % CI.

| Model | AUROC ↑ (95 % CI) | AUPRC ↑ (95 % CI) | F1 ↑ (95 % CI) |
| --- | --- | --- | --- |
| RFM | **0.974 (0.970–0.977)** | **0.705 (0.687–0.725)** | **0.674 (0.656–0.691)** |
| DINOv3b-9L [18] | 0.966 (0.961–0.970) | 0.635 (0.620–0.652) | 0.585 (0.569–0.601) |
| DINOv3b [18] | 0.957 (0.952–0.962) | 0.585 (0.569–0.602) | 0.520 (0.505–0.534) |
| RETFound-MAE [8] | 0.970 (0.965–0.974) | 0.674 (0.659–0.691) | 0.620 (0.604–0.637) |
| RETFound-DINOv2 [27] | 0.965 (0.961–0.970) | 0.637 (0.620–0.654) | 0.598 (0.579–0.614) |
| RETFound-Green [9] | 0.962 (0.957–0.966) | 0.616 (0.600–0.632) | 0.571 (0.553–0.589) |

**Table 8.** Smoking-status classification from frozen retinal embeddings on UK Biobank. AUROC, AUPRC, and F1 with 95 % CI. Bold/underline rank the frozen-linear models only; the published end-to-end results below the rule use different protocols and are shown for context, not ranked.

| Model | AUROC ↑ (95 % CI) | AUPRC ↑ (95 % CI) | F1 ↑ (95 % CI) |
| --- | --- | --- | --- |
| RFM | **0.767 (0.753–0.780)** | **0.323 (0.297–0.350)** | **0.112 (0.089–0.137)** |
| DINOv3b-9L [18] | 0.707 (0.693–0.720) | 0.224 (0.205–0.245) | 0.001 (0.000–0.006) |
| DINOv3b [18] | 0.579 (0.566–0.592) | 0.112 (0.106–0.120) | 0.098 (0.080–0.118) |
| RETFound-MAE [8] | <u>0.724 (0.709–0.738)</u> | <u>0.248 (0.227–0.272)</u> | 0.014 (0.006–0.023) |
| RETFound-DINOv2 [27] | 0.699 (0.685–0.715) | 0.211 (0.192–0.231) | 0.023 (0.013–0.034) |
| RETFound-Green [9] | 0.684 (0.671–0.698) | 0.193 (0.176–0.212) | 0.000 (0.000–0.000) |
| Poplin et al. [2] | 0.71 (0.70–0.73) | — | — |
| Korot et al. [42] | 0.64 | — | — |

### 4.2. Diabetic Retinopathy Classification

APTOS 2019 [32], IDRiD [33] and Messidor-2 [34, 35, 36] all use the same five-class international clinical DR severity scale; Table 9 gives the grade numbering used in the confusion matrices and per-class curves throughout this paper and the Supplementary Material, and per-split class counts for each dataset are given in Supplementary Tables S4–S6. Tables 10–12 report DR grading performance on the three datasets.

**Table 9.** Diabetic retinopathy grade numbering used in all confusion matrices and per-class ROC/PR curves in this paper and the Supplementary Material. NPDR: non-proliferative diabetic retinopathy; PDR: proliferative diabetic retinopathy.

| Plot label | DR severity class |
| --- | --- |
| Grade 0 | No DR |
| Grade 1 | Mild NPDR |
| Grade 2 | Moderate NPDR |
| Grade 3 | Severe NPDR |
| Grade 4 | PDR (proliferative DR) |

**Table 10.** Diabetic retinopathy grading on APTOS 2019 [32]. Frozen backbone + linear head. AUROC, AUPRC, and F1 with 95 % CI.

| Model | AUROC ↑ (95 % CI) | AUPRC ↑ (95 % CI) | F1 ↑ (95 % CI) |
| --- | --- | --- | --- |
| RFM | **0.946 (0.935–0.957)** | **0.706 (0.669–0.756)** | **0.667 (0.625–0.706)** |
| DINOv3b-9L [18] | 0.942 (0.931–0.952) | 0.672 (0.639–0.714) | 0.614 (0.573–0.654) |
| DINOv3b [18] | 0.933 (0.923–0.944) | 0.637 (0.606–0.679) | 0.536 (0.495–0.578) |
| RETFound-MAE [8] | 0.925 (0.913–0.937) | 0.629 (0.598–0.674) | 0.553 (0.517–0.590) |
| RETFound-DINOv2 [27] | 0.922 (0.910–0.934) | 0.620 (0.586–0.665) | 0.627 (0.588–0.665) |
| RETFound-Green [9] | 0.940 (0.929–0.950) | 0.651 (0.618–0.698) | 0.563 (0.518–0.603) |

**Table 11.** Diabetic retinopathy grading on IDRiD [33]. Frozen backbone + linear head. AUROC, AUPRC, and F1 with 95 % CI.

| Model | AUROC ↑ (95 % CI) | AUPRC ↑ (95 % CI) | F1 ↑ (95 % CI) |
| --- | --- | --- | --- |
| RFM | 0.825 (0.771–0.880) | **0.551 (0.484–0.658)** | **0.464 (0.359–0.567)** |
| DINOv3b-9L [18] | **0.842 (0.794–0.887)** | 0.534 (0.468–0.640) | 0.419 (0.336–0.502) |
| DINOv3b [18] | 0.796 (0.742–0.850) | 0.497 (0.434–0.593) | 0.410 (0.310–0.496) |
| RETFound-MAE [8] | 0.754 (0.690–0.812) | 0.455 (0.396–0.551) | 0.346 (0.255–0.433) |
| RETFound-DINOv2 [27] | 0.774 (0.719–0.838) | 0.472 (0.405–0.581) | 0.398 (0.301–0.489) |
| RETFound-Green [9] | 0.814 (0.771–0.861) | 0.500 (0.440–0.591) | 0.405 (0.316–0.493) |

**Table 12.**
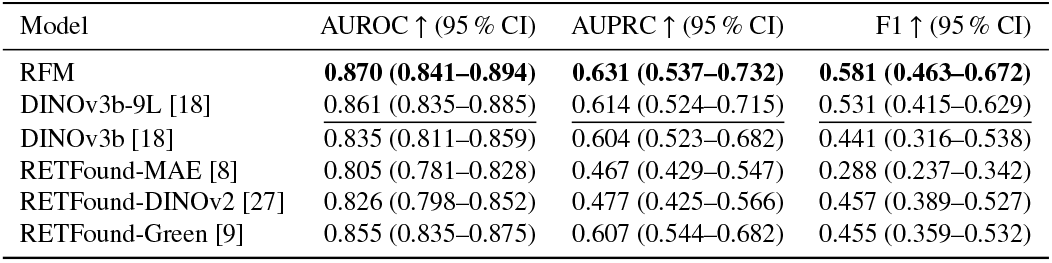
Diabetic retinopathy grading on Messidor-2 [34, 35, 36]. Frozen backbone + linear head. AUROC, AUPRC, and F1 with 95 % CI.

| Model | AUROC ↑ (95 % CI) | AUPRC ↑ (95 % CI) | F1 ↑ (95 % CI) |
| --- | --- | --- | --- |
| RFM | **0.870 (0.841–0.894)** | **0.631 (0.537–0.732)** | **0.581 (0.463–0.672)** |
| DINOv3b-9L [18] | 0.861 (0.835–0.885) | 0.614 (0.524–0.715) | 0.531 (0.415–0.629) |
| DINOv3b [18] | 0.835 (0.811–0.859) | 0.604 (0.523–0.682) | 0.441 (0.316–0.538) |
| RETFound-MAE [8] | 0.805 (0.781–0.828) | 0.467 (0.429–0.547) | 0.288 (0.237–0.342) |
| RETFound-DINOv2 [27] | 0.826 (0.798–0.852) | 0.477 (0.425–0.566) | 0.457 (0.389–0.527) |
| RETFound-Green [9] | 0.855 (0.835–0.875) | 0.607 (0.544–0.682) | 0.455 (0.359–0.532) |

RFM achieves the highest AUROC on two of the three DR benchmarks, APTOS (0.946) and Messidor-2 (0.870), and the highest AUPRC and F1 on all three. On both of those datasets DINOv3b-9L is second on AUROC (0.942 and 0.861), ahead of every RETFound variant. The ordering below RFM is metric-dependent: on APTOS F1 the runner-up is RETFound-DINOv2 (0.627), ahead of DINOv3b-9L (0.614). IDRiD is the exception (Figure 6): DINOv3b-9L leads on AUROC (0.842) with RFM at 0.825, although RFM retains the best AUPRC (0.551 vs. 0.534) and F1 (0.464 vs. 0.419) on that dataset. IDRiD is also by some margin the smallest DR benchmark, with 103 test images (Supplementary Table S5), and its confidence intervals are correspondingly the widest of the three.

**Figure 6.**
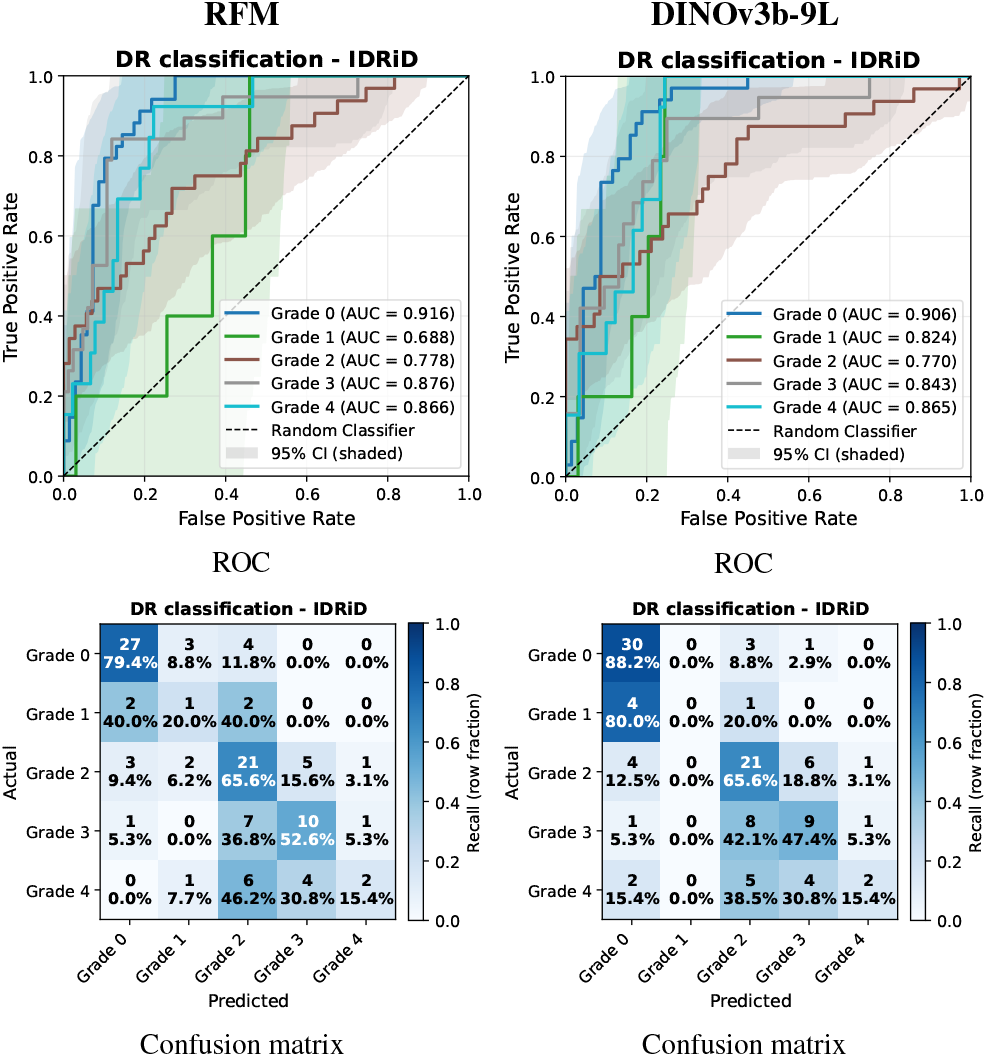
Per-class ROC curves (top row) and confusion matrices of the argmax prediction (bottom row; rows: true class, columns: predicted class) for diabetic retinopathy grading on IDRiD.

Two observations hold across all three datasets. First, RFM outperforms every RETFound variant on AUROC, though by small margins: the margin over the strongest RETFound variant ranges from 0.006 on APTOS (0.946 versus 0.940) to 0.015 on Messidor-2 (0.870 versus 0.855). Second, RETFound-Green is the strongest of the RETFound family on all three DR benchmarks despite being the smallest model evaluated, and is close enough to DINOv3b-9L (0.940 versus 0.942 on APTOS, 0.855 versus 0.861 on Messidor-2) that the two cannot be separated on these data. The residual gap between RFM and DINOv3b-9L on DR, and its interaction with feature extraction, is examined in Section 5.2.

### 4.2. Glaucoma Detection

Tables 13 and 14 report glaucoma detection performance on GFID and PAPILA.

**Table 13.** Glaucoma detection on GFID [15]. Frozen backbone + linear head. AUROC, AUPRC, and F1 with 95 % CI.

| Model | AUROC ↑ (95 % CI) | AUPRC ↑ (95 % CI) | F1 ↑ (95 % CI) |
| --- | --- | --- | --- |
| RFM | **0.957 (0.944–0.969)** | **0.888 (0.851–0.921)** | **0.814 (0.775–0.851)** |
| DINOv3b-9L [18] | 0.921 (0.900–0.939) | 0.821 (0.779–0.862) | 0.745 (0.698–0.785) |
| DINOv3b [18] | 0.893 (0.870–0.915) | 0.768 (0.726–0.813) | 0.714 (0.669–0.757) |
| RETFound-MAE [8] | 0.919 (0.900–0.937) | 0.802 (0.761–0.846) | 0.719 (0.667–0.764) |
| RETFound-DINOv2 [27] | 0.917 (0.897–0.937) | 0.802 (0.763–0.845) | 0.720 (0.676–0.764) |
| RETFound-Green [9] | 0.910 (0.888–0.933) | 0.790 (0.749–0.838) | 0.685 (0.636–0.734) |

**Table 14.**
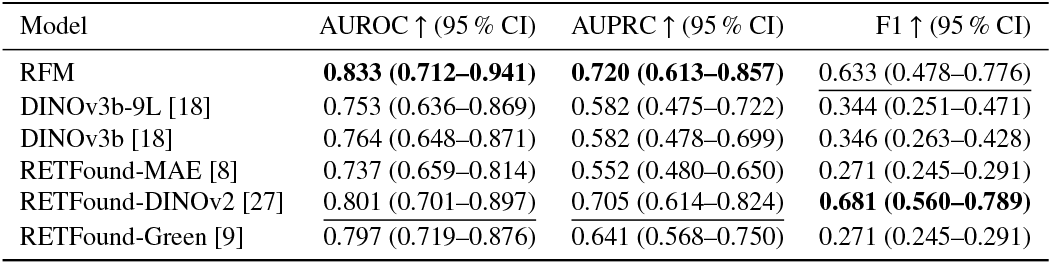
Glaucoma detection on PAPILA [16]. Frozen backbone + linear head. AUROC, AUPRC, and F1 with 95 % CI.

| Model | AUROC ↑ (95 % CI) | AUPRC ↑ (95 % CI) | F1 ↑ (95 % CI) |
| --- | --- | --- | --- |
| RFM | **0.833 (0.712–0.941)** | **0.720 (0.613–0.857)** | 0.633 (0.478–0.776) |
| DINOv3b-9L [18] | 0.753 (0.636–0.869) | 0.582 (0.475–0.722) | 0.344 (0.251–0.471) |
| DINOv3b [18] | 0.764 (0.648–0.871) | 0.582 (0.478–0.699) | 0.346 (0.263–0.428) |
| RETFound-MAE [8] | 0.737 (0.659–0.814) | 0.552 (0.480–0.650) | 0.271 (0.245–0.291) |
| RETFound-DINOv2 [27] | 0.801 (0.701–0.897) | 0.705 (0.614–0.824) | **0.681 (0.560–0.789)** |
| RETFound-Green [9] | 0.797 (0.719–0.876) | 0.641 (0.568–0.750) | 0.271 (0.245–0.291) |

RFM achieves the strongest performance on both glaucoma datasets, and by a wider margin than on DR. On GFID (Figure 7), RFM reaches AUROC 0.957 (95% CI: 0.944–0.969) and AUPRC 0.888 (0.851–0.921), ahead of DINOv3b-9L (0.921), RETFound-MAE (0.919) and RETFound-DINOv2 (0.917), which are closely grouped. On PAPILA, RFM leads with AUROC 0.833 (0.712–0.941) and AUPRC 0.720 (0.613–0.857), ahead of RETFound-DINOv2 (0.801, 0.701–0.897); PAPILA is a substantially smaller dataset than GFID (98 test images, Supplementary Table S8), and the overlapping confidence intervals indicate that this ranking should be treated with caution. On the argmax prediction RETFound-DINOv2 attains the higher F1 on this dataset (0.681 versus 0.633), so the ordering on PAPILA is metric-dependent.

**Figure 7.**
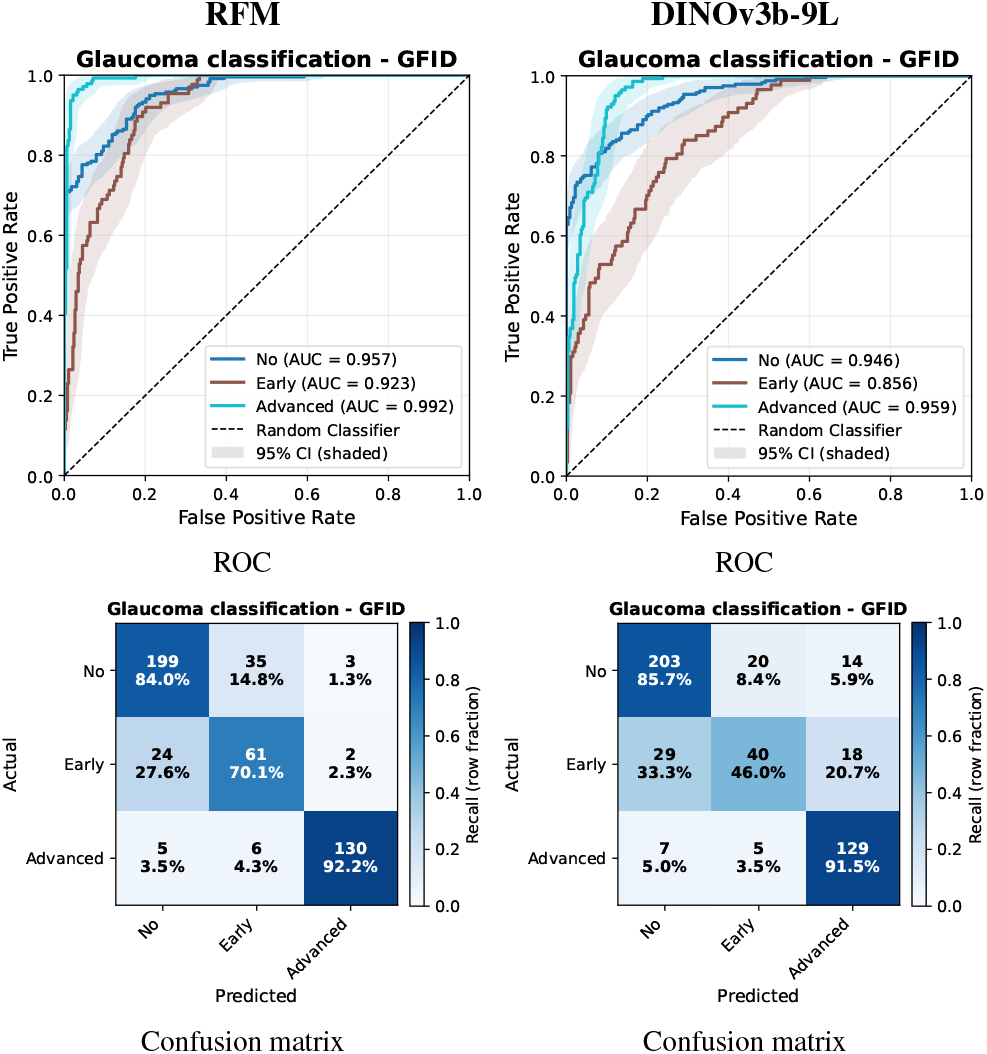
Per-class ROC curves (top row) and confusion matrices of the argmax prediction (bottom row; rows: true class, columns: predicted class) for glaucoma detection on GFID.

Retinal domain-specific SSL provides an advantage for glaucoma detection that is robust on GFID and directionally consistent on the smaller PAPILA cohort, where the margin is larger but the confidence intervals are much wider, in contrast with the DR findings above. The natural-image DINOv3b-9L baseline, which led or was second on every DR benchmark, drops to 0.753 on PAPILA, behind two of the three RETFound variants.

### 4.4. 3-Year Systemic Disease Incidence Prediction

Myocardial infarction and ischaemic stroke were selected as incidence endpoints given the established link between retinal microvascular features and systemic cardiovascular risk [2]. Parkinson’s disease is a distinct, neurodegenerative rather than vascular, disease process; it is included here because retinal imaging has separately been proposed as a candidate screening modality for neurodegenerative disease [3], and its inclusion tests whether frozen RFM features generalise to incidence prediction beyond the cardiovascular domain.

#### 4.4.1. Balanced evaluation design

The 1:1 construction of Section 3.3.3 leaves AUROC unchanged, AUROC being invariant to class prevalence, and fixes the AUPRC no-skill baseline at 0.5 so that AUPRC is comparable across models rather than being driven by how rare the endpoint happens to be in each cohort.

For the external UK Biobank evaluation a single trained head was scored against five random control draws, which together scored 524 (MI), 598 (stroke) and 55 (Parkinson’s disease) distinct control participants. Rather than treat those draws as five separate experiments to be averaged, we pool the participants they scored and estimate one quantity: discrimination in that scored population at a 50 % prevalence, with a percentile bootstrap over 4000 replicates for the interval and class weighting rather than subsampling to hold the prevalence at 50 %. Supplementary Section 4 gives the construction in full. Because the incident cases are an order of magnitude fewer than the controls, the resulting variability is dominated by the case count.

Figures 8–10 are drawn from the same replicates. The band they show is a pointwise interval on the curve, not the confidence interval quoted for AUROC or AUPRC in the table.

**Figure 8.**
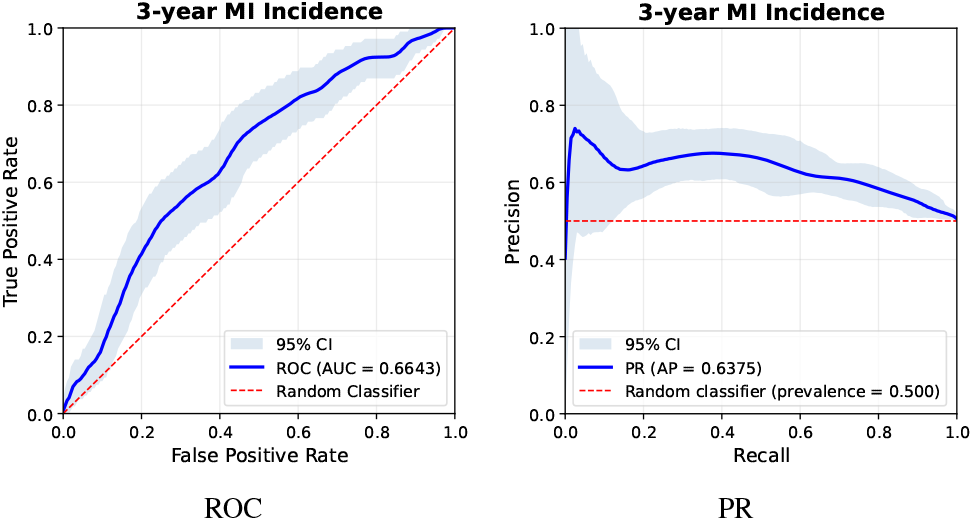
3-year myocardial infarction (MI) incidence, external transfer to UK Biobank. Mean over 4000 bootstrap replicates with the 2.5–97.5 percentile band (Section 4.4.1). Each replicate resamples the 107 cases and the 524 controls scored for this endpoint at their observed sizes; controls are weighted to a 50% prevalence, so the no-skill precision is 0.5.

The internal DESP incidence figures (Section 4.4.2) are not computed this way: their intervals come from the per-image bootstrap used for every other table in this paper, so the internal and external incidence intervals are not constructed identically and should not be compared as if they were.

#### 4.4.2. Internal evaluation

Tables 15, 16, and 17 report internal cohort results. As noted in Section 3.4, RFM heads are trained and evaluated on DESP and RETFound heads on AlzEye; the RFM and RETFound rows are therefore measurements on different cohorts, and the absolute AUROC values are not comparable between them (Section 3.4.6).

**Table 15.** Internal evaluation of 3-year MI incidence. RFM trained/evaluated on DESP; RETFound baselines on AlzEye. AUROC and AUPRC with 95 % CI. ^†^Value reported by the cited publication, not reproduced by us.

| Model | Dataset | AUROC ↑ (95 % CI) | AUPRC ↑ (95 % CI) |
| --- | --- | --- | --- |
| RFM | DESP | 0.665 (0.652–0.676) | 0.631 (0.615–0.648) |
| RETFound-MAE [8]† | AlzEye | 0.737 (0.731–0.743) | 0.726 (0.720–0.733) |
| RETFound-DINOv2 [27]† | AlzEye | 0.708 (0.677–0.741) | 0.693 (0.658–0.730) |

**Table 16.**
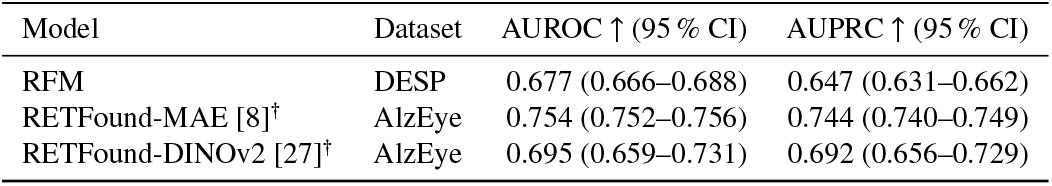
Internal evaluation of 3-year ischaemic stroke incidence. RFM trained/evaluated on DESP; RETFound baselines on AlzEye. AUROC and AUPRC with 95 % CI. ^†^Value reported by the cited publication, not reproduced by us.

**Table 17.** Internal evaluation of 3-year Parkinson’s disease incidence. RFM trained/evaluated on DESP; RETFound baselines on AlzEye. AUROC and AUPRC with 95 % CI. ^†^Value reported by the cited publication, not reproduced by us.

| Model | Dataset | AUROC ↑ (95 % CI) | AUPRC ↑ (95 % CI) |
| --- | --- | --- | --- |
| RFM | DESP | 0.705 (0.651–0.758) | 0.684 (0.609–0.753) |
| RETFound-MAE [8]† | AlzEye | 0.669 (0.650–0.688) | 0.662 (0.641–0.683) |
| RETFound-DINOv2 [27]† | AlzEye | 0.633 (0.525–0.734) | 0.637 (0.533–0.731) |

On the DESP dataset, RFM achieves AUROC of 0.665 for MI incidence, 0.677 for stroke, and 0.705 for Parkinson’s disease. RETFound variants on AlzEye report higher AUROC for MI and stroke (up to 0.737 and 0.754 respectively for RETFound-MAE), but this reflects dataset differences rather than a controlled model comparison. On the Parkinson’s disease endpoint the ordering is reversed: RFM on DESP attains 0.705 against 0.669 for RETFound-MAE on AlzEye [8]. As with MI and stroke, the two figures come from different cohorts and the difference is not attributable to the models.

#### 4.4.3. External transfer to UK Biobank without retraining

Tables 18, 19, and 20 report external UKBB results. In this evaluation, heads trained on DESP (for RFM) or AlzEye (for RETFound) are applied without retraining to the UKBB cohort, and no UK Biobank incidence label was used to fit them (Section 3.4.6).

**Table 18.** External evaluation of 3-year MI incidence on UK Biobank. RFM head trained on DESP; applied without retraining. AUROC and AUPRC with 95 % CI (Section 4.4.1). ^†^Value reported by the cited publication, not reproduced by us.

| Model | AUROC ↑ (95 % CI) | AUPRC ↑ (95 % CI) |
| --- | --- | --- |
| RFM | **0.664 (0.610–0.718)** | **0.638 (0.584–0.696)** |
| RETFound-MAE [8]<sup>†</sup> | 0.579 (0.566–0.593) | 0.563 (0.551–0.575) |
| RETFound-DINOv2 [27]<sup>†</sup> | <u>0.620</u> | <u>0.605</u> |

**Table 19.** External evaluation of 3-year ischaemic stroke incidence on UK Biobank. RFM head trained on DESP; applied without retraining. AUROC and AUPRC with 95 % CI (Section 4.4.1). ^†^Value reported by the cited publication, not reproduced by us.

| Model | AUROC ↑ (95 % CI) | AUPRC ↑ (95 % CI) |
| --- | --- | --- |
| RFM | **0.685 (0.635–0.736)** | **0.671 (0.622–0.727)** |
| RETFound-MAE [8]<sup>†</sup> | 0.594 (0.580–0.608) | 0.587 (0.575–0.601) |
| RETFound-DINOv2 [27]<sup>†</sup> | 0.545 | 0.535 |

**Table 20.** External evaluation of 3-year Parkinson’s disease incidence on UK Biobank. RFM head trained on DESP; applied without retraining. AUROC and AUPRC with 95 % CI (Section 4.4.1). ^†^Value reported by the cited publication,not reproduced by us.

| Model | AUROC ↑ (95 % CI) | AUPRC ↑ (95 % CI) |
| --- | --- | --- |
| RFM | **0.643 (0.491–0.793)** | 0.591 (0.517–0.749) |
| RETFound-MAE [8]<sup>†</sup> | <u>0.570 (0.554–0.586)</u> | **0.598 (0.561–0.637)** |
| RETFound-DINOv2 [27]<sup>†</sup> | 0.560 | 0.585 |

RFM’s AUROC exceeds that of every RETFound variant on all three external UKBB endpoints, though the evidence differs sharply in strength between them. For MI, RFM achieves AUROC 0.664 (0.610–0.718) versus RETFound-DINOv2 at 0.620 and RETFound-MAE at 0.579. For stroke, RFM achieves AUROC 0.685 (0.635–0.736) against a maximum RETFound AUROC of 0.594 (RETFound-MAE). For Parkinson’s disease, RFM achieves AUROC 0.643 (0.491–0.793), above RETFoundMAE (0.570) and RETFound-DINOv2 (0.560), but this endpoint rests on only 11 incident cases (Supplementary Table S3); the confidence interval spans 0.5, the individual control draws range from 0.529 to 0.769 AUROC (Supplementary Table S16), and on AUPRC RFM (0.591) falls below RETFound-MAE (0.598). The result should be regarded as inconclusive rather than as evidence of an advantage. The MI and stroke endpoints, with 107 and 120 incident cases, therefore carry the substantive evidence that RFM features transfer to an unseen external cohort (Section 5.4). Figures 8–10 show the corresponding ROC and precision–recall curves.

**Figure 9.**
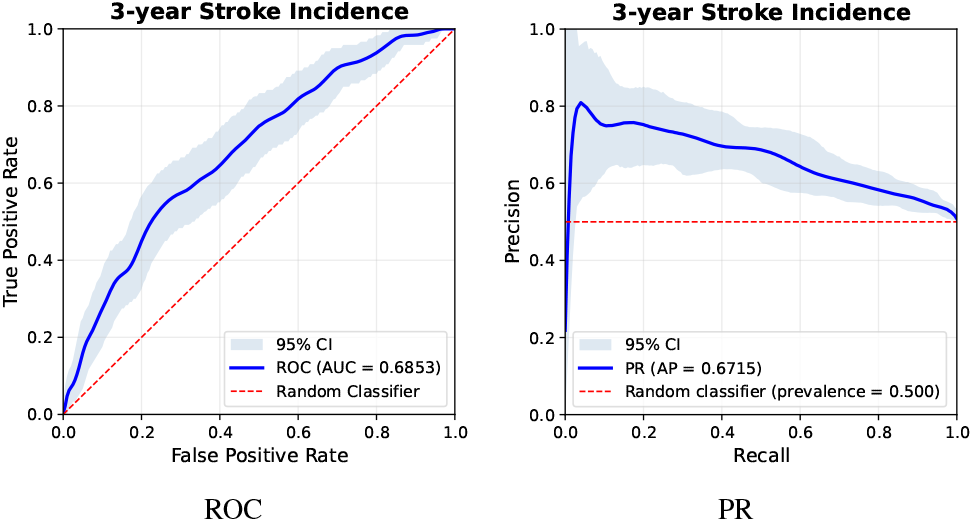
3-year ischaemic stroke incidence, external transfer to UK Biobank. Mean over 4000 bootstrap replicates with the 2.5–97.5 percentile band (Section 4.4.1). Each replicate resamples the 120 cases and the 598 controls scored for this endpoint at their observed sizes; controls are weighted to a 50% prevalence, so the no-skill precision is 0.5.

**Figure 10.**
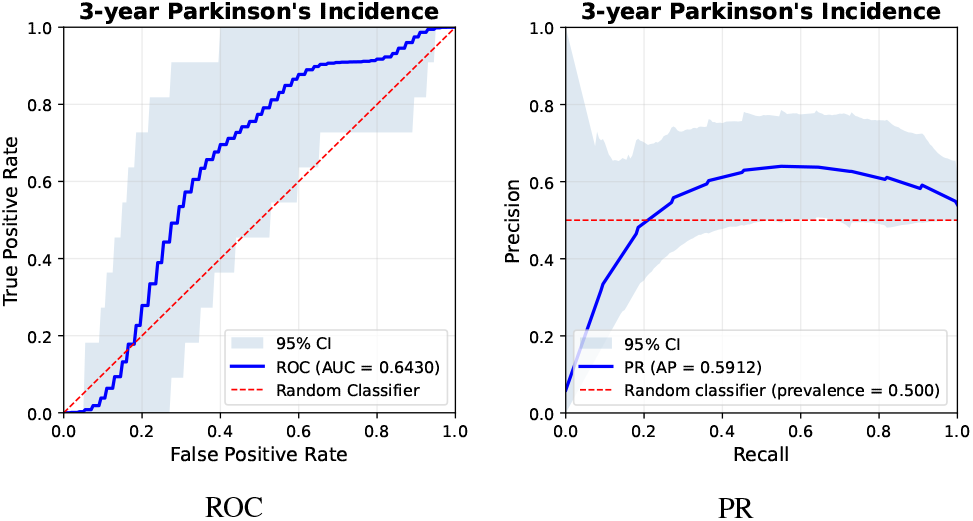
3-year Parkinson’s disease incidence, external transfer to UK Biobank. Mean over 4000 bootstrap replicates with the 2.5–97.5 percentile band (Section 4.4.1). Each replicate resamples the 11 cases and the 55 controls scored for this endpoint at their observed sizes; controls are weighted to a 50% prevalence, so the no-skill precision is 0.5.

## 5. Discussion

### 5.1. Parameter Efficiency and Comparison with Existing Models

RFM achieves strong frozen-linear performance with 64 million parameters – 4.8× fewer than the 307M-parameter RETFound [8]. While ultra-compact architectures such as RETFound-Green operate at an even smaller scale (22M) [9], RFM delivers a substantial reduction in model capacity relative to standard foundation models without sacrificing representation quality. Across all evaluated tasks, RFM performs competitively with, or superior to, RETFound-MAE under the frozen-linear protocol. This indicates that the parameter efficiency of ViT-Base architectures is well-preserved when paired with an effective SSL objective and pre-training corpus. From a deployment perspective, smaller backbones reduce inference latency, memory consumption, and integration overhead, especially in multi-modal pipelines where the vision encoder must process inputs alongside other system components. The nine-layer truncation alone accounts for a measured 24.4% reduction in inference time relative to the full DINOv3-Base encoder (Section 3.1).

Figure 11 plots mean AUROC against backbone parameter count across the subset of classification, diabetic retinopathy, and glaucoma tasks on which every model was evaluated, avoiding biases from differing evaluation coverage. It plots two families: the published retinal foundation models, and the two readings of the un-adapted natural-image DINOv3-Base checkpoint from which RFM is trained. RFM attains the highest mean AU-ROC among the compared models (0.885) while operating with markedly fewer parameters than the per-task best RETFound variant (0.846 at 307M), and it also exceeds the smallest retinal model, RETFound-Green (0.843 at 22M).

**Figure 11.**
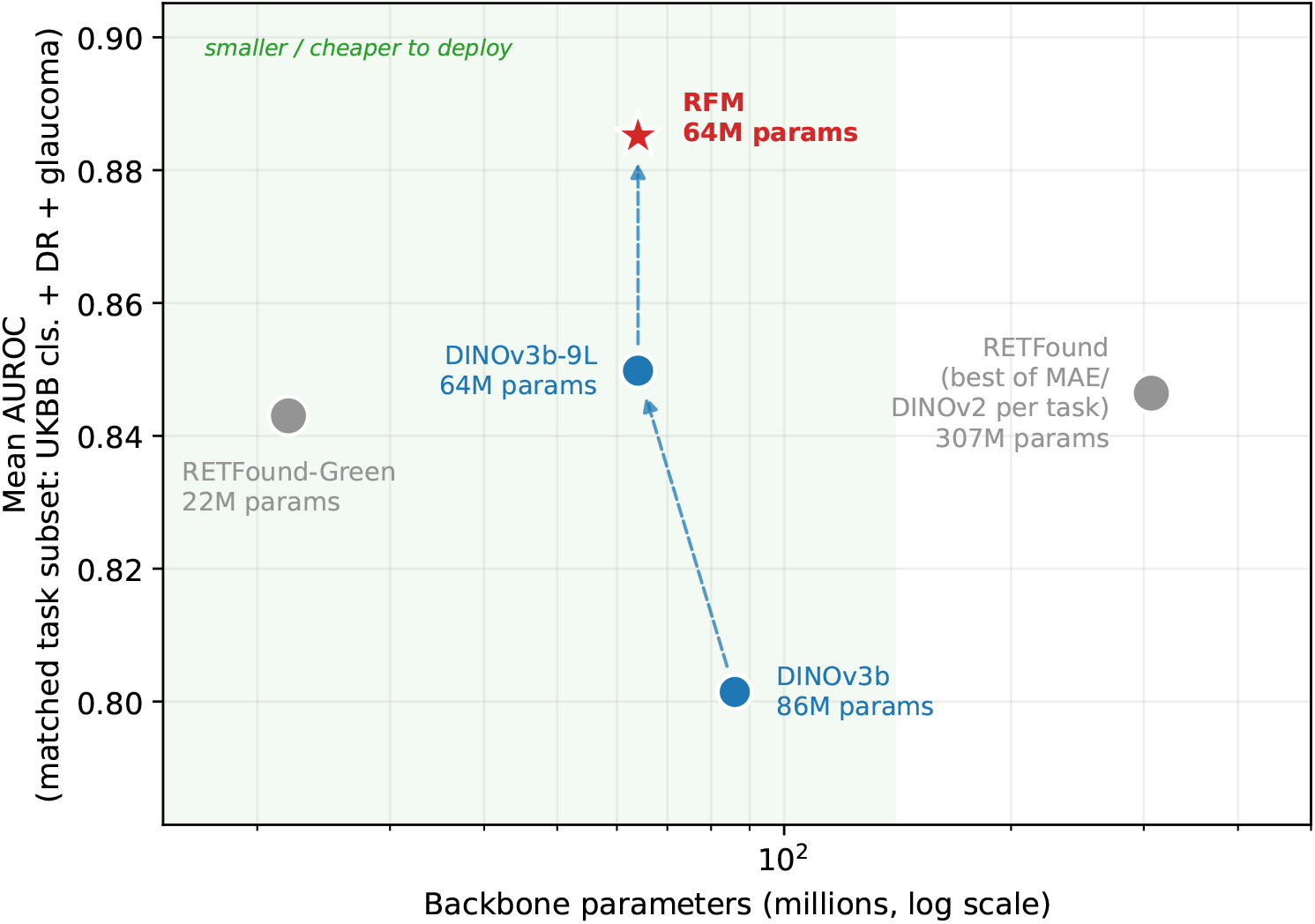
Frozen-linear mean AUROC vs. backbone parameter count, on the 8 classification/DR/glaucoma tasks common to RFM and the best-performing RETFound variant per task. Each point is a mean over all 8 tasks. Every model is evaluated on the same task subset and under the same head-training protocol, so no model is rewarded for narrower evaluation coverage. Grey points are the published retinal foundation models. The RETFound point takes, on each task, the higher AUROC of RETFound-MAE and RETFound-DINOv2, so it is a per-task envelope rather than a single model, and favours RETFound. The two blue points are the natural-image DINOv3-Base checkpoint before any retinal adaptation, read two ways: DINOv3b, the unmodified 86M-parameter twelve-layer encoder as released, read by global average pooling of the final-layer patch tokens; and DINOv3b-9L, truncated after the ninth layer and read with the CLS-plus-mean-patch features used for RFM (64M parameters). The dashed arrows trace the two ablation steps that separate the released checkpoint from RFM – first the readout, then retinal LeJEPA pre-training – each of which raises mean AUROC, the first while also removing a quarter of the inference cost. 3-year incidence tasks are excluded throughout (only evaluated for RFM and RETFound, whose heads were trained on different cohorts – DESP vs. AlzEye – so not a controlled comparison).

The dashed arrows in Figure 11 trace the two steps that separate the released checkpoint from RFM, each isolating one decision. Taken as released (DINOv3b: twelve layers, global average pooling of the final layer), DINOv3-Base reaches a mean AUROC of 0.801 on the matched subset, below every retinal foundation model evaluated here. Reading the same weights at the truncated ninth layer with CLS-plus-mean-patch pooling (DINOv3b-9L) raises that to 0.850 (0.049) while discarding a quarter of the network, so the gain is obtained at lower inference cost rather than in exchange for it; at that point the un-adapted natural-image backbone already matches the per-task best RETFound variant (0.846) with a 4.8 ×smaller backbone. Retinal LeJEPA pre-training then adds 0.035 to reach RFM’s 0.885, at identical architecture and inference cost, since RFM and DINOv3b-9L execute the same nine transformer layers under the same extraction. The two steps together are worth 0.084 mean AUROC. That the first step is comparable in size to the second is the substantive point: how a frozen backbone is read is not a presentational detail, and a natural-image ViT-Base already carries much of what these tasks require, with domain-specific pre-training refining those features towards clinically relevant retinal structure rather than supplying them from nothing.

### 5.2. Feature Extraction and the Natural-Image Backbone

Two findings deserve separate treatment: the contribution of the feature-extraction convention, which we can now measure directly on every benchmark, and the residual standing of the natural-image backbone relative to retinal-domain pre-training on DR.

#### 5.2.1. The extraction convention is a first-order effect

Frozen-linear evaluations differ in which transformer layer they read and how they pool it, and this choice is rarely reported as a design variable. Because we evaluated the same DINOv3-Base checkpoint twice, once truncated after the ninth layer with CLS-plus-mean-patch pooling and once with the conventional global pooling over the final layer, on the same splits and under the same head-training protocol, the difference between those two rows isolates the extraction recipe.

How much it is worth depends strongly on the task. Across the five ophthalmic benchmarks it reaches 0.046 AUROC, favouring the truncated CLS-plus-mean-patch features on four of the five and the conventional global pooling on PAPILA (Tables 10–14); the signed mean over the five is 0.020 AUROC in favour of truncation. On the UK Biobank phenotypes it is several times larger and uniform in direction: the truncated readout leads on all three classification phenotypes (Tables 6–8), by 0.154 AUROC for sex (0.654 to 0.808) and 0.128 for smoking status (0.579 to 0.707), with a mean of 0.097, and it attains the lower MAE on all seven regression biomarkers (Tables 1–5 and Supplementary Tables S12 and S13). As released, DINOv3b is close to chance on smoking status (0.579) and weak on sex (0.654); the same weights read at the ninth layer (DINOv3b-9L) reach 0.707 and 0.808. The systemic signal is therefore present in the mid-network representation and largely lost when the final layer is pooled, an effect the disease-grading benchmarks alone would not have revealed. The two rows differ in layer depth, pooling operator and feature dimensionality at once, so what is measured is the recipe as a whole; attributing the effect to any one of those three components would require a factorial ablation we did not run.

On the ophthalmic benchmarks the effect is small in absolute AUROC, but it is not too small to change comparative conclusions. It moves where the checkpoint falls among the comparators on four of the five datasets: on APTOS (0.933 to 0.942) and Messidor-2 (0.835 to 0.861) it passes RETFound-Green (0.940 and 0.855); on GFID (0.893 to 0.921) it moves from last to second, ahead of all three RETFound variants; and on IDRiD (0.796 to 0.842), where the effect is largest (0.046), it moves from third to first, passing both RETFound-Green (0.814) and RFM (0.825). On those same four benchmarks the extraction effect is larger than the margin separating the checkpoint from the nearest RETFound variant. PAPILA is the exception on both counts: it is the one benchmark where global pooling is ahead (0.764 versus 0.753), by a margin that leaves the ordering unchanged. IDRiD is accordingly the one benchmark on which the convention decides which model leads: RFM holds the highest AUROC on all five ophthalmic benchmarks against DINOv3b, and on four of the five against DINOv3b-9L.

The practical implication is that extraction convention is a real and reportable degree of freedom in frozen-linear benchmarking, and one worth holding fixed wherever the comparison permits. We hold it fixed between RFM and DINOv3b-9L, which differ only in pre-training; the RETFound rows are evaluated with each model’s own published convention, as stated in Section 5.5. A comparison spanning models with different conventions therefore carries an extraction-related offset of unmeasured size, which is why we report the convention for every row explicitly. The magnitude measured here is specific to this DINOv3-Base checkpoint: it gives no estimate of what RETFound would gain from an equivalent search over its own extraction recipe, so the RETFound figures here should be read as the performance of their published convention rather than as a ceiling for that model family.

#### 5.2.2. Natural-image pre-training remains competitive on DR

On DR grading RFM leads the RETFound variants only narrowly, and DINOv3b-9L, without any retinal adaptation, is close behind on APTOS and Messidor-2 and ahead of it on IDRiD (Section 4.2) – a markedly different picture from the glaucoma benchmarks, where retinal-domain pre-training gives RFM a clear advantage. Hou et al. [43] independently report the same generalist advantage on DR using DINOv2.

A plausible account is that the two task families reward different representations. DR grading turns on haemorrhages, microa-neurysms and exudates: lesions characterised by local texture at the scale of individual patches rather than by global structural organisation. DINO-family objectives, which do not reconstruct pixel intensities, are known to develop strong local feature detectors through cross-view prediction, whereas RFM’s pretext tasks, local crops included, present the retina at reduced resolution: each local crop spans 128–512 px of the 1024-px preprocessed image and is resized to 112× 112, so small lesions are seen downsampled by up to about 4.6×. Glaucoma assessment turns instead on optic disc and rim morphology, a global property, and that is where RFM’s advantage is largest.

A complementary explanation is training data contamination: APTOS 2019, IDRiD and Messidor-2 are public datasets whose images may have been included in the web-crawled corpus used for DINOv3 pre-training. Because RFM is initialised from that same checkpoint it would inherit any such exposure, so contamination cannot explain the difference *between* RFM and DINOv3b-9L; it would, however, inflate both relative to the RETFound variants, which were pre-trained on curated retinal corpora. We could neither confirm nor exclude this.

Either way the implication is the same: on texture-heavy pathology RFM’s pretext-task composition may not develop local detectors as well as it develops structural ones, so the retinal LeJEPA stage yields less over its natural-image initialization there than it does on glaucoma. The directions that follow are set out in Section 6.

### 5.3. LeJEPA as a Practical SSL Objective for Retinal Imaging

The LeJEPA objective offers practical advantages over competing SSL families for retinal foundation model training. Against DINO-family methods [24, 25] it dispenses with the stabilisation machinery listed in Section 3.1. Compared with Masked Autoencoders [26], LeJEPA operates entirely in latent representation space and avoids pixel-level reconstruction targets, so it is not required to model high-frequency image compression artefacts or clinical acquisition variability in its training signal. We did not measure sensitivity to either factor. This distinction is supported by direct evidence in the retinal domain: using identical compute budgets, Zhou et al. [10] trained matched retinal foundation models with a Masked Autoencoder and with DINOv2 (a self-distillation, non-reconstructive objective related in spirit to LeJEPA) and found the DINOv2-based model consistently outperformed the Masked Autoencoder variant across both pre-training cohorts and downstream tasks. The theoretical grounding of LeJEPA [12] provides guarantees against representational collapse without heuristic stabilisation, reducing the hyperparameter sensitivity that complicates large-scale medical image SSL where long pre-training runs are not easily repeatable.

Whether the projector adaptation warmup (Section 3.1.2) contributes to RFM’s age and biomarker performance relative to RETFound is a hypothesis we did not test: no matched run without the warmup was trained, so its effect on downstream representation quality remains unquantified.

The retinal stage is what separates RFM from the DINOv3-Base checkpoint it is initialised from (DINOv3b-9L), by 0.035 mean AUROC on the matched subset (Section 5.1); the incidence endpoints are outside this comparison, DINOv3b-9L not having been evaluated on them.

### 5.4. External Generalisation to UK Biobank

The UKBB results provide this study’s evidence on cross-cohort transfer, and two comparisons should be distinguished.

The first is internal to RFM and is directly controlled: the same frozen backbone, the same linear-head protocol and the same metric, applied to a cohort on which no head was trained. Moving from the internal DESP evaluation to the external UKBB evaluation, RFM’s AUROC point estimate is similar for MI (0.665 to 0.664) and marginally higher for ischaemic stroke (0.677 to 0.685), with overlapping confidence intervals, while for Parkinson’s disease it falls (0.705 to 0.643) on an endpoint with only 11 incident cases and a confidence interval spanning 0.5. For the two endpoints with usable event counts, frozen RFM features therefore show comparable observed discrimination in a London diabetic screening population and a UK-wide volunteer cohort recorded with different equipment and a different case mix. We did not test for equivalence, and intervals of this width would not exclude a modest loss. This claim rests on the study’s own design and does not depend on comparison with another model.

The second comparison is with RETFound. RFM’s AUROC exceeds the published RETFound figures on all three endpoints, though the Parkinson’s result is inconclusive (Section 4.4.3). What is compared is each model’s reported performance on the same cohort resource rather than on one shared test set (Section 3.4.6). The comparison is between the models as released and adapted, each with its own pre-training corpus, imaging domain and adaptation regime; it does not identify which of those ingredients is responsible, and nothing here shows that a particular pre-training objective or corpus yields more transferable representations in general.

These observations are consistent with a pattern reported across the RETFound line of work: cross-cohort transfer gaps concentrate in oculomics and systemic-incidence tasks rather than in ocular disease detection. Zhou et al. [10] isolate pretraining corpus and cohort distribution shift and find the subtler oculomics tasks most affected; the same group report a marked fall in systemic-disease AUROC when models adapted on AlzEye are evaluated on UK Biobank, alongside more stable ocular disease detection [27]; and the RETFound paper attributes its own external UK Biobank drop to demographic and imaging-device differences between MEH-AlzEye and UK Biobank [8]. The endpoints examined here fall in that more transfer-fragile class, so the absence of a measurable drop between DESP and UKBB for MI and ischaemic stroke is a useful observation; it is, however, a result for two endpoints in one external cohort and should not be read as establishing transfer behaviour in general.

Importantly, the same frozen RFM backbone, without any task-specific adaptation, supports linear heads for all ten UKBB biomarker phenotypes and three incidence tasks simultaneously. This property is a prerequisite for the intended use case of RFM as a vision encoder in multimodal architectures, where a single model must serve diverse downstream tasks without per-task backbone updates.

### 5.5. Limitations

Several limitations should be noted. First, this study evaluates only the frozen-linear protocol; fine-tuning with non-linear heads is left to future work. Second, the internal incidence figures are measurements on two different cohorts and are not a model comparison (Section 3.4.6). Third, the Parkinson’s disease endpoint rests on 11 incident events, and its interval spans 0.5. Fourth, RFM is not evaluated on retinal image segmentation tasks. Fifth, the prediction of sensitive demographic attributes (sex, ethnicity, smoking status) from fundus images raises privacy and fairness considerations [44]; these should be carefully considered in any deployment context. Sixth, the potential data contamination of DR benchmark datasets in DINOv3 training data could not be confirmed or excluded, limiting the interpretation of the DINOv3b-9L and DINOv3b DR results. Seventh, the internal DESP incidence heads are trained and evaluated on a subset of the same DESP corpus used for RFM’s self-supervised pre-training. Pre-training used no labels, but the images are not disjoint from the internal incidence evaluation set, so the internal DESP incidence figures should be read as in-domain performance, and the UK Biobank evaluation (Section 4.4.3) is the out-of-domain test. Eighth, the feature-extraction configuration was selected on UK Biobank validation-split data (Section 3.2). No participant in any reported evaluation contributed to that selection, but the configuration is nonetheless not blind to the UK Biobank resource itself, having been chosen on images from that cohort’s imaging protocol and population, and a configuration chosen on DESP alone might transfer differently. Ninth, unlike RETFound we did not evaluate 3-year heart failure incidence, because the corresponding UK Biobank phenotype was not available to us; our incidence comparison therefore covers three of the four endpoints reported by RETFound. Tenth, each backbone is evaluated with its own published feature-extraction convention (Section 3.4, Supplementary Table S11), and we measure what that is worth (Section 5.2); comparisons across models that do not share a convention therefore carry an offset of that order. Eleventh, we report bootstrap confidence intervals but no paired statistical test between models; where confidence intervals overlap, the ranking should be treated as indicative rather than established. Twelfth, all public-benchmark results here are computed on splits we generated (Section 3.3.2) and are therefore not directly comparable with previously published figures on these datasets, which is why every comparator was re-run rather than cited. Finally, this work does not include a systematic ablation of RFM’s self-supervised pre-training design choices (e.g. pretext-task crop composition, masking/crop ratio, projector-adaptation warmup schedule); we isolate the feature-extraction convention (Section 5.2) but leave a controlled ablation of pre-training design to future work.

## 6. Conclusion

We have presented RFM, a lightweight retinal foundation model trained with the LeJEPA joint-embedding predictive SSL objective on 4.3 million colour fundus photographs from a large NHS diabetic eye screening programme, using domain-specific pretext tasks and a projector adaptation warmup schedule to train a ViT-Base backbone stably at that scale. Under a strict frozen-backbone linear-head protocol it leads the retinal foundation models compared here on the regression and matched classification benchmarks (Section 4) at 4.8×fewer parameters than RETFound-MAE and RETFound-DINOv2, though more than RETFound-Green. These are observed differences: several are small relative to their confidence intervals, no superiority or equivalence test was performed, and on the smallest cohorts the ordering can depend on the metric, RETFound-DINOv2 attaining the higher F1 on PAPILA, computed on the argmax prediction. Applied without retraining to UK Biobank, RFM’s frozen features exceed the published RETFound AUROC for 3-year myocardial infarction and ischaemic stroke incidence; the Parkinson’s endpoint is inconclusive.

That the natural-image-pretrained DINOv3b-9L remains competitive with retinal-domain pre-training on DR grading, while falling clearly behind on glaucoma, is consistent with an interplay between pre-training domain and the spatial scale of the discriminative features, a plausible account that we did not test directly but one that opens directions for future pretext task design targeting texture-sensitive endpoints. LeJEPA is demonstrated to be a practical and theoretically grounded SSL objective for retinal foundation model training, avoiding the heuristic stabilisation requirements of competing families while producing representations with broad task transferability.

Beyond the fine-tuning and ablation studies noted in Section 5.5, future work will target three directions: pretext tasks at finer spatial scales, to improve RFM on texture-heavy pathology; a short further SSL phase at higher resolution (512 × 512 or larger), to exploit the detail modern fundus cameras capture; and integration of the frozen backbone into multimodal architectures coupling retinal images with clinical records and genomic data, the motivating application for a generalised, parameter-efficient foundation model.

## Data Availability

All data produced in the present work are contained in the manuscript and the code is available at https://github.com/ok1zjf/RFM

## Ethics Statement

The DESP dataset comprises clinical images collected as part of routine NHS diabetic eye screening. All data were managed according to UK NHS information governance requirements, in accordance with the principles of the Data Protection Act 2018. Methodologies adhere to the Homerton Healthcare NHS Foundation Trust Security Policy and Information Governance Policy, and General Data Protection Regulation principles relating to the processing of personal data. The study has been approved by the Health Research Authority (IRAS project ID 265637). The NHS Health Research Authority toolkit identified that research ethics approval was not required for this project, as all data are pseudonymised and presented in aggregate form. A Data Protection Impact Assessment was submitted to the Homerton Healthcare NHS Foundation Trust Information Governance Lead and approved by the information governance team in December 2022.

The UK Biobank study was carried out following the principles of the Declaration of Helsinki and the Research Governance Framework for Health and Social Care. The UK Biobank study was approved by the North West Multi-Centre Research Ethics Committee (11/NW/0382). All participants gave written, informed consent.

### Collaborators

UK Biobank Eye and Vision Consortium and The Artificial Intelligence Automated Retinal Image Analysis Systems (ARIAS) Research Group: John Anderson (Diabetes and Endocrinology, Homerton Healthcare NHS Foundation Trust, London, UK), Sarah Barman (School of Computer Science and Mathematics, Kingston University, London, UK), Louis Bolter (Diabetes and Endocrinology, Homerton Healthcare NHS Foundation Trust, London, UK), Ryan Chambers (Diabetes and Endocrinology, Homerton Healthcare NHS Foundation Trust, London, UK), Lakshmi Chandrasekaran (St George’s School of Health and Medical Sciences, City St George’s, University of London, London, UK), Umar Chaudhry (St George’s School of Health and Medical Sciences, City St George’s, University of London, London, UK), Catherine Egan (NIHR Biomedical Research Centre, Moorfields Eye Hospital NHS Foundation Trust and UCL Institute of Ophthalmology, London, UK), Jiri Fajtl (School of Computer Science and Mathematics, Kingston University, London, UK), Aaron Lee (Department of Ophthalmology, University of Washington, Seattle, WA, USA), Samantha Mann (South East London Diabetic Eye Screening Programme, Guy’s and St Thomas’ NHS Foundation Trust, London, UK), Abraham Olvera-Barrios (NIHR Biomedical Research Centre, Moorfields Eye Hospital NHS Foundation Trust and UCL Institute of Ophthalmology, London, UK), Christopher G Owen (St George’s School of Health and Medical Sciences, City St George’s, University of London, London, UK), Paolo Remagnino (Department of Computer Science, Durham University, Durham, UK), Alicja R Rudnicka (St George’s School of Health and Medical Sciences, City St George’s, University of London, London, UK), Adnan Tufail (NIHR Biomedical Research Centre, Moorfields Eye Hospital NHS Foundation Trust and UCL Institute of Ophthalmology, London, UK), Charlotte Wahlich (St George’s School of Health and Medical Sciences, City St George’s, University of London, London, UK), Laura Webster (South East London Diabetic Eye Screening Programme, Guy’s and St Thomas’ NHS Foundation Trust, London, UK), Roshan Welikala (School of Computer Science and Mathematics, Kingston University, London, UK), Kathryn Willis (St George’s School of Health and

Medical Sciences, City St George’s, University of London, London, UK).

## Data Availability Statement

The North East London DESP dataset is not publicly available due to patient privacy constraints and NHS data governance requirements. UK Biobank data are available to approved researchers via the UK Biobank Access Management System (https://www.ukbiobank.ac.uk/enable-your-research/apply-for-access). All public benchmark datasets (APTOS 2019, IDRiD, Messidor-2, GFID, PA-PILA) are available at their respective repositories cited in the text. RFM source code, pretrained model weights, and the train/validation/test splits used for all public benchmark datasets (APTOS 2019, IDRiD, Messidor-2, GFID, PAPILA) are available at https://github.com/ok1zjf/RFM.

## Acknowledgements

This research has been conducted using the UK Biobank Resource under Application Number 522. This work is funded by The Wellcome Collaborative Award (2022, 224390/Z/21/Z). We wish to thank Niall Canavan, Sean Devine and Asif Mirza from the Homerton Healthcare NHS Foundation Trust IT department for their continued support throughout this project.

## Footnotes

2 Embeddings are gathered across all GPUs before the statistic is computed, so *N* is the global mini-batch size rather than the per-device batch.

3 https://community.ukbiobank.ac.uk/hc/en-gb/articles/24842092764061-Reporting-small-numbers-in-results-in-research-outputs-using-UK-Biobank-data

4 The variant is named for its Masked Autoencoder objective; the abbreviation is unrelated to the mean absolute error metric used for regression tasks here

