## Supplementary Material for "RFM: A Lightweight Retinal Foundation Model for Generalised Oculomics"

**Abstract**

This document reports supplementary material referenced from the main paper: descriptions of the DESP, UK Biobank and five public benchmark datasets and the composition of every cohort and split; the complete configurations for self-supervised pre-training and for linear-head training, shared and per backbone; LDL and HDL cholesterol regression on UK Biobank (full results tables); per-draw AUROC/AUPRC/F1 tables and per-draw ROC/PR curve plots for the three external UK Biobank 3-year incidence models (RFM, five balanced control draws); predicted-vs-true density plots for all seven UK Biobank regression biomarkers; and precision–recall (PR) curves for the classification tasks whose main-paper figures show only the complementary receiver operating characteristic (ROC) view. All results use the frozen-backbone linear-head protocol described in the main paper; table and figure captions repeat the essential context so this document can be read independently.

* Corresponding author
[1] Members are listed in the Collaborators section of the main paper.

### 1. Public Benchmark Datasets and Data Splits

This section describes each dataset used in the main paper and reports the composition of every cohort and how each was divided into training, validation and test sets.

#### *1.1. Split construction*

All splits were generated in-house rather than adopted from existing partitions, so that one uniform data-preparation protocol applies across every benchmark (Section 3.3.2 of the main paper). That protocol is: patient-level grouping wherever subject identifiers are published, class-stratified allocation across training, validation and test, and label assignment verified against each dataset's own released metadata. Split definitions are released with the source code.

Three of the five public benchmarks do not release subject identifiers or laterality tags. GFID is distributed as a cross-sectional collection of optic-disc crops, APTOS 2019 anonymises images behind randomised 12-character hashes, and IDRiD provides only per-image grades. For these we partition at the image level and rely on the independence of images asserted by the source distribution; if any of them does contain multiple images per patient, a residual risk of within-patient correlation across splits remains. PAPILA and Messidor-2 both publish subject identifiers and laterality, and are split strictly at the patient level.

#### *1.2. Dataset descriptions*

*DESP.* The North East London Diabetic Eye Screening Programme dataset [1] supplies the 4.3 million images used for self-supervised pre-training and the internal 3-year incidence cohort. Raw screening captures occasionally include non-retinal images (e.g. of the crystalline lens, eyelids, or hands), taken to document anterior-segment pathology or to confirm camera function when a retinal image could not be obtained [2]. Image quality was assessed with the QUARTZ tool [3] and images below a minimum quality threshold were excluded, which removes the non-retinal captures alongside blurred or otherwise unusable retinal ones. A detailed characterisation of the cohort is given in a dedicated cohort profile [4].

*UK Biobank.* UK Biobank (UKBB) [5, 6, 7] is a population-based prospective cohort study comprising 502,682 adults aged 40–69 years recruited from 22 UK centres, with baseline biomedical and physical assessments conducted between 2006 and 2013. Retinal imaging was performed in a subset of 85,746 participants using a Topcon 3D-OCT 1000 Mark 2 fundus camera, yielding 175,856 non-mydriatic, macula-centred images from both eyes, with a 45° field of view (FOV) and a resolution of $2048 \times 1536$ pixels. The UKBB study was approved by the North West Multi-Centre Research Ethics Committee (11/NW/0382). The QUARTZ tool [3] was used to exclude low-quality images. Following image quality filtering and the exclusion of participants who had withdrawn consent as of July 2025, the final dataset comprised 117,902 images from 66,299 unique participants.

*PAPILA.* The PAPILA (Patient-specific Analysis of Retinal Fundus Images and Clinical Data for Glaucoma Assessment) dataset [8] was collected by the Department of Ophthalmology at the Hospital General Universitario Reina Sofía, Spain, between 2018 and 2020. It comprises 488 images at $2576 \times 1934$ pixels, captured with a Topcon TRC-NW400 non-mydriatic retinal camera. Clinical metadata and systematic file naming explicitly link left and right eyes to individual subject identifiers, allowing strict patient-level splitting. Images are classified into three diagnostic categories: healthy; glaucoma (confirmed chronic simple glaucoma); and suspicious/uncertain, an intermediate borderline group. Distributed via Figshare under CC BY 4.0.

*GFID.* The Glaucoma Fundus Image Dataset [9] comprises 1,544 fundus photographs cropped to a $240 \times 240$ pixel region around the optic nerve head, collected at Kim's Eye Hospital, South Korea, using a Nidek AFC-330 non-mydriatic camera. The collection is cross-sectional and releases neither patient identifiers nor laterality tags, so each image is treated as an independent sample. Images are labelled across three severity categories: normal/control (no glaucomatous structural damage); early glaucoma (initial structural defects or early nerve thinning); and advanced glaucoma (severe glaucomatous damage with profound optic disc excavation). Distributed via Harvard Dataverse under CC0.

*APTOS 2019.* The APTOS 2019 Blindness Detection dataset [10] was collected by Aravind Eye Hospital across clinics in rural India to support automated diabetic retinopathy screening. It consists of 3,662 labelled training fundus images captured across multiple camera models, giving heterogeneous resolutions (approximately $474 \times 358$ to $3216 \times 2136$ pixels). Images are anonymised using randomised 12-character hexadecimal hashes with no patient identifiers or laterality metadata, requiring image-level partitioning. Ground truth is graded on the five-point International Clinical Diabetic Retinopathy (ICDR) scale.

*Messidor-2.* Messidor-2 [11, 12, 13] extends the original Messidor repository and was collected across three ophthalmology departments in France (Brest, Paris and Saint-Étienne). The release used here contains 1,744 macula-centred colour fundus images from 874 patients, acquired with a Topcon TRC-NW6 non-mydriatic camera (45° field of view) at $1440 \times 960$, $2240 \times 1488$ and $2304 \times 1536$ pixels. The accompanying reference metadata pairs both eyes for each patient via a patient identifier and laterality field, enabling strict patient-level splits that keep both eyes of an individual within one split. Images are graded on the five-point ICDR scale.

*IDRiD.* The Indian Diabetic Retinopathy Image Dataset [14] was acquired at Shri Guruoji Netralaya eye clinic in Nanded, Maharashtra, India. The disease-grading subset contains 516 macula- and optic-disc-centred fundus photographs captured with a Kowa VX-10$\alpha$ camera at a 50° field of view and $4288 \times 2848$ resolution. Images are sequentially indexed per split and the public metadata provides only image-level DR grades, without subject identifiers or laterality tags, precluding patient-level grouping. Each image is graded by retinal specialists on the five-point ICDR scale. Distributed via IEEE DataPort under CC BY 4.0.

#### *1.3. Cohort composition and per-split counts*

Tables S1–S8 report the composition of every cohort used in this work.

Two conventions apply throughout and are repeated in the individual captions so each table can be read on its own. First, the 3-year incidence sets (Table S3) follow the RETFound evaluation protocol and are balanced 1:1 between incident events and randomly sampled controls, which fixes the no-skill AUPRC baseline at 0.5 and makes the AUPRC values in the main paper's incidence tables comparable across models; only left-eye images are used for these tasks. Second, categories marked * in Table S1 exist in the raw UK Biobank data but were excluded from the corresponding experiment, for the reasons given in Section 4.1.2 of the main paper.

The event counts in Table S3 are also the direct explanation for the precision of the incidence results: the UK Biobank external test sets contain 107 myocardial infarction, 120 ischaemic stroke and only 11 Parkinson's disease events, and the confidence intervals reported in the main paper widen accordingly.

Table S1: UK Biobank categorical phenotype counts by train/validation/test split (sex, ethnicity, smoking status). All counts in this table are **images**, not participants. Splits are 70/10/20 and are drawn at the **participant** level, so no participant contributes images to more than one split, but a participant may contribute several images to their own split through both eyes and repeat visits. Percentages in the phenotype rows are within-split class shares; percentages in the **Total** rows are the share of the whole cohort held by that split. Categories marked * were present in the raw data but were excluded from the corresponding experiment: for smoking we retained only the unambiguous never-smoker and current-smoker groups (Section 4.1.2 of the main paper), and for ethnicity we excluded the "Unknown/didn't answer" group. The 117,902 images come from 66,299 unique participants.

| Phenotype | Category | Train | Val | Test | Total |
|---|---|---|---|---|---|
| Sex | Male | 36805 (45%) | 5261 (45%) | 10439 (44%) | 52505 (45%) |
| | Female | 45726 (55%) | 6529 (55%) | 13142 (56%) | 65397 (55%) |
| | **Total** | 82531 (70%) | 11790 (10%) | 23581 (20%) | 117902 (100%) |
| Ethnicity | White | 76434 (93%) | 10857 (92%) | 21783 (92%) | 109074 (93%) |
| | Black | 1857 (2%) | 276 (2%) | 524 (2%) | 2657 (2%) |
| | Asian | 1874 (2%) | 270 (2%) | 567 (2%) | 2711 (2%) |
| | Other | 1903 (2%) | 313 (3%) | 574 (2%) | 2790 (2%) |
| | Unknown/didn't answer* | 463 (1%) | 74 (1%) | 133 (1%) | 670 (1%) |
| | **Total** | 82531 (70%) | 11790 (10%) | 23581 (20%) | 117902 (100%) |
| Smoking | Never | 46910 (57%) | 6731 (57%) | 13559 (57%) | 67200 (57%) |
| | Current | 5007 (6%) | 677 (6%) | 1390 (6%) | 7074 (6%) |
| | Occasionally* | 2124 (3%) | 322 (3%) | 635 (3%) | 3081 (3%) |
| | Prefer not to say/ missing* | 414 (1%) | 75 (1%) | 127 (1%) | 616 (1%) |
| | Previous* | 28076 (34%) | 3985 (34%) | 7870 (33%) | 39931 (34%) |
| | **Total** | 82531 (70%) | 11790 (10%) | 23581 (20%) | 117902 (100%) |

Table S2: UK Biobank continuous biomarker statistics by train/validation/test split (age, systolic and diastolic blood pressure, HbA1c, LDL, HDL, BMI). Splits are 70/10/20 at the **participant** level, so no participant appears in more than one split, but each may contribute several images. *Samples* counts **images** with a non-missing value for that biomarker, which is why the totals differ between biomarkers and from Table S1; percentages are the share of that biomarker's cohort held by each split. *MAE* is the mean absolute error of a mean-predictor baseline within the split, i.e. the reference against which the relative MAE improvement in the main paper's regression tables is computed. The close agreement of mean, standard deviation and baseline MAE across splits indicates the participant-level split did not introduce distributional shift. The images derive from the same 66,299 unique participants as Table S1.

| Phenotype | Statistic | Train | Val | Test | Total |
|---|---|---|---|---|---|
| Age (year) | Samples | 82396 (70%) | 11771 (10%) | 23535 (20%) | 117702 (100%) |
| | Mean | 56.6 | 56.6 | 56.6 | 56.6 |
| | Std dev | 8.2 | 8.2 | 8.2 | 8.2 |
| | MAE | 7.0 | 7.1 | 7.0 | 7.0 |
| SBP (mmHg) | Samples | 82246 (70%) | 11760 (10%) | 23489 (20%) | 117495 (100%) |
| | Mean | 136.6 | 136.5 | 136.7 | 136.6 |
| | Std dev | 18.3 | 18.1 | 18.3 | 18.3 |
| | MAE | 14.5 | 14.4 | 14.6 | 14.5 |
| DBP (mmHg) | Samples | 82247 (70%) | 11760 (10%) | 23490 (20%) | 117497 (100%) |
| | Mean | 81.4 | 81.4 | 81.4 | 81.4 |
| | Std dev | 10.0 | 10.0 | 10.0 | 10.0 |
| | MAE | 7.9 | 8.0 | 8.0 | 7.9 |
| HbA1c (mmol/mol) | Samples | 71703 (70%) | 10187 (10%) | 20468 (20%) | 102358 (100%) |
| | Mean | 35.9 | 35.8 | 35.8 | 35.9 |
| | Std dev | 6.3 | 6.0 | 6.2 | 6.2 |
| | MAE | 3.7 | 3.7 | 3.7 | 3.7 |
| LDL (mmol/L) | Samples | 75140 (70%) | 10664 (10%) | 21496 (20%) | 107300 (100%) |
| | Mean | 3.55 | 3.56 | 3.55 | 3.55 |
| | Std dev | 0.86 | 0.87 | 0.86 | 0.86 |
| | MAE | 0.69 | 0.69 | 0.68 | 0.69 |
| HDL (mmol/L) | Samples | 70763 (70%) | 10067 (10%) | 20307 (20%) | 101137 (100%) |
| | Mean | 1.49 | 1.49 | 1.50 | 1.49 |
| | Std dev | 0.39 | 0.39 | 0.40 | 0.39 |
| | MAE | 0.31 | 0.31 | 0.31 | 0.31 |
| BMI (kg/m$^2$) | Samples | 81987 (70%) | 11705 (10%) | 23427 (20%) | 117119 (100%) |
| | Mean | 27.2 | 27.2 | 27.1 | 27.2 |
| | Std dev | 4.7 | 4.7 | 4.7 | 4.7 |
| | MAE | 3.6 | 3.6 | 3.6 | 3.6 |

Table S3: Event and control counts for the 3-year incidence tasks, on the internal DESP cohort (train, validation and test) and the external UK Biobank test cohort. The DESP validation split is listed because head checkpoints are selected on validation performance. The DESP validation and test sets are equal-sized halves of the held-out participants. Following the RETFound 3-year incidence evaluation protocol, every set is constructed 1:1: all eligible incident events are retained and an equal number of controls is drawn at random, so Total $N = 2 \times$ Events. For the external UK Biobank cohort this table therefore reports two quantities. *Per control draw* gives the counts of one such balanced draw, as used for Tables S14–S16. *Pooled* gives the distinct participants scored across the five draws, which is the sample the headline external estimates in the main paper are computed on; there the 50 % prevalence is imposed by weighting the controls rather than by matching their number to the events, so the no-skill AUPRC baseline is 0.5 in both cases (Section 4.4.1 of the main paper). Splits are at the **participant** level and only left-eye images are used. Outcomes are defined relative to the baseline imaging date; participants with the target condition recorded at or before baseline are excluded, events are those occurring within three years of baseline, and controls require at least three event-free years of follow-up (Section 3.3.3 of the main paper). Each entry is a single, unique left-eye image from one participant at one visit: no participant contributed multiple images or repeated visits to these sets, so the image, the eye and the participant are the same unit throughout. The small number of incident Parkinson's disease cases in UK Biobank ($n = 11$) is the reason for the wide confidence intervals on that endpoint.

| | DESP Train | | | DESP Val | | | DESP Test | | | UKBB External Test (per control draw) | | | UKBB External Test (pooled) | | |
|---|---|---|---|---|---|---|---|---|---|---|---|---|---|---|---|
| Task | Events | Controls | Total N | Events | Controls | Total N | Events | Controls | Total N | Events | Controls | Total N | Events | Controls | Total N |
| MI | 12932 | 12932 | 25864 | 4101 | 4101 | 8202 | 4101 | 4101 | 8202 | 107 | 107 | 214 | 107 | 524 | 631 |
| Stroke | 12349 | 12349 | 24698 | 4280 | 4280 | 8560 | 4280 | 4280 | 8560 | 120 | 120 | 240 | 120 | 598 | 718 |
| Parkinson's | 474 | 474 | 948 | 176 | 176 | 352 | 176 | 176 | 352 | 11 | 11 | 22 | 11 | 55 | 66 |

Table S4: APTOS 2019 diabetic retinopathy grade distribution by train/validation/test split, using the five-class international clinical DR severity scale (Table 9 of the main paper). Percentages in the class rows are within-split class shares; percentages in the **Total** row are the share of the dataset held by each split. Counts are images. This dataset releases no subject identifiers, so splitting is necessarily at the **image** level (Section 3.3.2 of the main paper). APTOS is the largest of the three DR benchmarks (3,662 images), with 49% of images graded no DR.

| Phenotype | Category | Train | Val | Test | Total |
|---|---|---|---|---|---|
| Grade | No DR | 1009 (49%) | 254 (49%) | 542 (49%) | 1805 (49%) |
| | Mild NPDR | 207 (10%) | 52 (10%) | 111 (10%) | 370 (10%) |
| | Moderate NPDR | 559 (27%) | 140 (27%) | 300 (27%) | 999 (27%) |
| | Severe NPDR | 108 (5%) | 27 (5%) | 58 (5%) | 193 (5%) |
| | PDR (Proliferative DR) | 165 (8%) | 41 (8%) | 89 (8%) | 295 (8%) |
| | **Total** | 2048 (56%) | 514 (14%) | 1100 (30%) | 3662 (100%) |

Table S5: IDRiD diabetic retinopathy grade distribution by train/validation/test split, using the five-class international clinical DR severity scale (Table 9 of the main paper). Percentages in the class rows are within-split class shares; percentages in the **Total** row are the share of the dataset held by each split. Counts are images. This dataset releases no subject identifiers, so splitting is necessarily at the **image** level (Section 3.3.2 of the main paper). IDRiD is the smallest DR benchmark (516 images, 103 in test) and has the most even grade distribution but the fewest images per grade, which accounts for the wide confidence intervals on the IDRiD results in the main paper.

| Phenotype | Category | Train | Val | Test | Total |
|---|---|---|---|---|---|
| Grade | No DR | 107 (33%) | 27 (32%) | 34 (33%) | 168 (33%) |
| | Mild NPDR | 16 (5%) | 4 (5%) | 5 (5%) | 25 (5%) |
| | Moderate NPDR | 108 (33%) | 28 (33%) | 32 (31%) | 168 (33%) |
| | Severe NPDR | 59 (18%) | 15 (18%) | 19 (18%) | 93 (18%) |
| | PDR (Proliferative DR) | 39 (12%) | 10 (12%) | 13 (13%) | 62 (12%) |
| | **Total** | 329 (64%) | 84 (16%) | 103 (20%) | 516 (100%) |

Table S6: Messidor-2 diabetic retinopathy grade distribution by train/validation/test split, using the five-class international clinical DR severity scale (Table 9 of the main paper). Percentages in the class rows are within-split class shares; percentages in the **Total** row are the share of the dataset held by each split. Splits are at the **patient** level. Note the strong class imbalance: proliferative DR accounts for 2% of images, which is why macro-averaged F1, computed on the argmax prediction and weighting each grade equally, is a noisier summary for this task than AUROC.

| Phenotype | Category | Train | Val | Test | Total |
|---|---|---|---|---|---|
| Grade | No DR | 570 (58%) | 143 (58%) | 304 (58%) | 1017 (58%) |
| | Mild NPDR | 151 (15%) | 38 (15%) | 81 (16%) | 270 (15%) |
| | Moderate NPDR | 194 (20%) | 49 (20%) | 104 (20%) | 347 (20%) |
| | Severe NPDR | 42 (4%) | 11 (4%) | 22 (4%) | 75 (4%) |
| | PDR (Proliferative DR) | 20 (2%) | 5 (2%) | 10 (2%) | 35 (2%) |
| | **Total** | 977 (56%) | 246 (14%) | 521 (30%) | 1744 (100%) |

Table S7: Glaucoma Fundus Image Dataset (GFID) class distribution by train/validation/test split. Percentages in the class rows are within-split class shares; percentages in the **Total** row are the share of the dataset held by each split. Counts are images. This dataset releases no subject identifiers, so splitting is necessarily at the **image** level (Section 3.3.2 of the main paper). GFID is evaluated as a three-class grading task; the AUROC, AUPRC and F1 reported in the main paper (Section 4.3) are macro-averaged over these three classes, and the confusion matrix in Figure 7 is correspondingly $3 \times 3$.

| Phenotype | Category | Train | Val | Test | Total |
|---|---|---|---|---|---|
| Grade | No glaucoma | 440 (51%) | 111 (51%) | 237 (51%) | 788 (51%) |
| | Early glaucoma | 161 (19%) | 41 (19%) | 87 (19%) | 289 (19%) |
| | Advanced glaucoma | 260 (30%) | 66 (30%) | 141 (30%) | 467 (30%) |
| | **Total** | 861 (56%) | 218 (14%) | 465 (30%) | 1544 (100%) |

Table S8: PAPILA class distribution by train/validation/test split. Percentages in the class rows are within-split class shares; percentages in the **Total** row are the share of the dataset held by each split. PAPILA is evaluated as a three-class grading task, as for GFID. Splits are at the **patient** level, which matters for PAPILA in particular because it provides both eyes of each patient: a naive image-level split would place fellow eyes of the same patient in both training and test. PAPILA is the smallest ophthalmic benchmark used in this work (98 test images), which is why its confidence intervals are the widest of the ophthalmic benchmarks.

| Phenotype | Category | Train | Val | Test | Total |
|---|---|---|---|---|---|
| Grade | Healthy | 212 (68%) | 54 (69%) | 67 (68%) | 333 (68%) |
| | Glaucoma | 56 (18%) | 14 (18%) | 17 (17%) | 87 (18%) |
| | Suspicious | 44 (14%) | 10 (13%) | 14 (14%) | 68 (14%) |
| | **Total** | 312 (64%) | 78 (16%) | 98 (20%) | 488 (100%) |

### 2. Training Configurations

This section lists the complete configurations for the two training regimes described in the main paper: self-supervised pre-training of the RFM backbone (Section 3.1) and supervised training of the linear prediction heads (Section 3.2). Every backbone is frozen and probed with a single affine head under the shared protocol of Table S10, on identical splits; the settings that follow each backbone's own published feature-extraction and linear-probe recipe are listed per model in Table S11.

Table S9: Self-supervised pre-training configuration. The augmentation rows are applied in the order listed. Global crops are taken from the full image area; local crops are constrained to the circular valid-pixel region of the fundus (Section 3.1.1 of the main paper). "$[a, b]$" denotes a range sampled uniformly.

| Group | Parameter | Value |
|---|---|---|
| Backbone | architecture | DINOv3-Base ViT, patch 16 |
| | initialisation | LVD-1689M pre-trained checkpoint |
| | SSL input resolution | 256 × 256 |
| | mixed precision | enabled |
| LeJEPA objective | projector $g$ | 768 → 2048 → 2048 → 256 |
| | projector norm / act. | BatchNorm1d / ReLU |
| | sketched directions $S$ | 1024 |
| | integration domain | $t \in [0, 5]$, 17 quadrature nodes |
| | balance $\lambda$ | 0.05 |
| Optimisation | optimiser | AdamW |
| | base learning rate | $1 \times 10^{-4}$ |
| | weight decay | 0.05 |
| | batch size | 220 per GPU (880 over 4 GPUs) |
| | epochs | 200 |
| | gradient-norm clipping | 1.0 |
| LR schedule | projector-only warmup | epochs 0–5 (backbone frozen) |
| | backbone warmup | epochs 5–10 (linear) |
| | decay | cosine, to end of training |
| Global crops (2 per image) | random resized crop | scale $[0.4, 1.0]$ |
| | horizontal flip | $p = 0.5$ |
| | vertical flip | $p = 0.5$ |
| | rotation | $[-30°, 30°]$ |
| | Gaussian blur | $p = 0.5$ |
| | output size | 256 × 256 |
| Local crops (6 per image) | circular crop | radius fraction 0.99 |
| | crop side | $[128, 512]$ px |
| | horizontal flip | $p = 0.5$ |
| | vertical flip | $p = 0.5$ |
| | rotation | $[-30°, 30°]$ |
| | colour jitter | $p = 0.8$; brightness, contrast, saturation 0.2; hue 0.1 |
| | output size | 112 × 112 |

Table S10: Linear-head training and evaluation protocol shared by every backbone on all benchmarks. The backbone is frozen throughout; only the affine head is trained. Augmentation rows are applied in the order listed and are used at training time only. Settings that differ between backbones are given per model in Table S11.

| Group | Parameter | Value |
|---|---|---|
| Features | backbone | frozen, no gradient updates |
| | head | single affine layer |
| | head input | pooled backbone feature (per-model, Table S11) |
| Optimisation | optimiser | AdamW |
| | mixed precision | bfloat16 autocast |
| | checkpoint selection | best validation AUROC (classification), MAE (regression) |
| Objective | loss | cross-entropy (classification), L1 (regression) |
| | class weighting | none (all classes weighted equally) |
| Train-time augmentation | pre-resize (GFID only) | shorter edge to 512 px |
| | random resized crop | scale $[0.9, 1.0]$, aspect ratio 1:1 |
| | horizontal flip | $p = 0.5$ |
| | vertical flip | $p = 0.5$ |
| | Gaussian blur | $p = 1.0$, $\sigma \in [0.3, 0.9]$, $k = 7$ |
| | rotation | $[-45°, 45°]$ |
| Validation and test | transform | resize + centre crop, no augmentation (crop margin per model) |
| | normalisation | per-channel, each backbone's own pre-training statistics |

Table S11: Backbone-specific linear-head settings. Each RETFound model keeps its own published input resolution, normalisation and feature extraction; DINOv3b-9L and DINOv3b carry RFM's input and head settings. RETFound-MAE and RETFound-DINOv2 also follow RETFound's official linear-probe schedule.[†] RETFound-Green's reference implementation[‡] instead fits a scikit-learn logistic regression on standardised features; we replace it with the shared AdamW-trained affine head used for RFM, so that the head-fitting procedure is not a confound (Section 3.4.4). DESP statistics are the per-channel mean and standard deviation of the DESP pre-training corpus, $(0.427, 0.247, 0.137)/(0.261, 0.162, 0.093)$; ImageNet statistics are the standard $(0.485, 0.456, 0.406)/(0.229, 0.224, 0.225)$. RFM and DINOv3b-9L share a column: they are the same truncated backbone read with the same extraction convention and trained with the same head settings, and differ only in the pre-trained weights. All rows share the protocol of Table S10.

| | RFM / DINOv3b-9L | DINOv3b | RETFound-MAE | RETFound-DINOv2 | RETFound-Green |
|---|---|---|---|---|---|
| Backbone | ViT-B/16 (9 blocks) | ViT-B/16 (12 blocks) | ViT-L/16 | ViT-L/14 | ViT-S/14 (reg4) |
| Input resolution | 336 | 336 | 224 | 224 | 392 |
| Feature layer | 9th block | final | final | final | final |
| Pooling | CLS $\oplus$ mean patch | mean patch | mean patch | CLS | mean patch (post-norm) |
| Feature dim. | 1536 | 768 | 1024 | 1024 | 384 |
| Normalisation | DESP | DESP | ImageNet | ImageNet | 0.5/0.5 |
| Learning rate | $1 \times 10^{-3}$ | $1 \times 10^{-3}$ | $4.7 \times 10^{-4}$[a] | $4.7 \times 10^{-4}$[a] | $1 \times 10^{-3}$ |
| LR schedule | cosine | cosine | warmup + cosine[b] | warmup + cosine[b] | cosine |
| Weight decay | $5 \times 10^{-5}$ | $5 \times 10^{-5}$ | 0.05 | 0.05 | $5 \times 10^{-6}$[c] |
| Batch size | 64 | 64 | 24 | 24 | 64 |
| Epochs | 200 | 200 | 50 | 50 | 200 |
| Eval crop margin | none | none | 0.875 | 0.875 | none |

[†] RETFound linear-probe reference implementation: `https://github.com/rmaphoh/RETFound/tree/main`. [‡] RETFound-Green reference implementation: `https://github.com/justinengelmann/RETFound_Green/`. [a] RETFound scales its reference learning rate by batch size: $\text{lr} = 5 \times 10^{-3} \times 24/256$. [b] 10 epochs of linear warmup followed by half-cycle cosine decay to $1 \times 10^{-6}$, as in RETFound's own linear-probe schedule; the other backbones use flat-start cosine annealing over all 200 epochs. [c] $5 \times 10^{-5}$ on IDRiD and PAPILA.

### 3. Supplementary Tables: LDL and HDL Cholesterol

Tables S12 and S13 report frozen-linear prediction of LDL and HDL cholesterol on UK Biobank; these two biomarkers are omitted from the main paper's regression tables (Section 4.1.1) for space and are reported here in full.

Table S12: LDL cholesterol (mmol/L) prediction from frozen retinal embeddings on UK Biobank. No published end-to-end results for this phenotype.

| Model | MAE ↓ (95 % CI) | Rel. MAE ↑ | $\rho$ ↑ (95 % CI) |
|---|---:|---:|---:|
| RFM | **0.675 (0.668–0.682)** | **1.7%** | **0.130 (0.117–0.144)** |
| DINOv3b-9L [15] | 0.678 (0.672–0.686) | 1.1% | 0.088 (0.075–0.101) |
| DINOv3b [15] | 0.687 (0.681–0.695) | −0.1% | 0.075 (0.061–0.087) |
| RETFound-MAE [16] | 0.678 (0.671–0.685) | 1.2% | 0.100 (0.087–0.113) |
| RETFound-DINOv2 [17] | 0.679 (0.672–0.687) | 1.0% | 0.093 (0.081–0.107) |
| RETFound-Green [18] | 0.680 (0.673–0.687) | 0.9% | 0.072 (0.059–0.085) |

Table S13: HDL cholesterol (mmol/L) prediction from frozen retinal embeddings on UK Biobank. No published end-to-end results for this phenotype.

| Model | MAE ↓ (95 % CI) | Rel. MAE ↑ | $\rho$ ↑ (95 % CI) |
|---|---:|---:|---:|
| RFM | **0.285 (0.282–0.289)** | **7.6%** | **0.373 (0.360–0.385)** |
| DINOv3b-9L [15] | 0.295 (0.292–0.299) | 4.3% | 0.290 (0.277–0.301) |
| DINOv3b [15] | 0.303 (0.301–0.308) | 1.6% | 0.252 (0.238–0.264) |
| RETFound-MAE [16] | 0.293 (0.291–0.297) | 4.8% | 0.311 (0.299–0.324) |
| RETFound-DINOv2 [17] | 0.294 (0.291–0.298) | 4.7% | 0.294 (0.281–0.306) |
| RETFound-Green [18] | 0.296 (0.294–0.300) | 3.9% | 0.272 (0.260–0.285) |

### 4. Supplementary Tables: Per-Draw 3-Year Incidence Results

Tables S14–S16 report AUROC, AUPRC, and F1 for each of the five balanced test-control draws made for the RFM external UK Biobank 3-year incidence evaluation. RETFound baselines are evaluated as a single fine-tuned model and their published figures carry no draw-level breakdown (Section 3.4.3 of the main paper), so they are not included here. $F_1$ is shown here for completeness but is not reported in the main paper, because it depends on a per-endpoint decision threshold that the published RETFound figures do not share.

These five draws are *not* cross-validation folds and should not be read as such. All incident (positive) cases are present in every draw; only the controls are redrawn, by random sampling from the non-event pool at a 1:1 ratio (Table S3). The draws therefore differ solely in control composition and are not statistically independent of one another, so the spread across them measures sensitivity to the choice of controls alone and not the sampling variability of an independent test set.

The main paper therefore reports a single estimate over the pooled participants rather than the average of these five draws (Section 4.4.1 of the main paper). Its construction is as follows. The point estimate is the metric computed on all distinct scored participants. The interval is a percentile bootstrap in which the incident cases and the controls are each resampled with replacement at their own observed sizes, which is the standard bootstrap of the observed sample; so constructed, it reflects the sampling variability of the scored participants in both classes. The 50 % prevalence is imposed by class weighting rather than by subsampling: each control carries weight $n_{\text{cases}}/n_{\text{controls}}$, so the two weighted class totals are equal in every replicate and the AUPRC no-skill baseline is 0.5 throughout, matching the balanced protocol under which the published RETFound figures were computed. AUROC needs no weighting, being invariant to prevalence. Balance therefore takes two forms: an individual draw has equal participant counts, the pooled estimate equal weighted class totals.

The main paper's incidence figures are drawn from the same replicate distribution, with the precision–recall curves weighted to the same 50 % prevalence as the tabulated AUPRC. Each panel plots the pointwise mean curve over replicates together with the 2.5–97.5 percentile band; the mean ROC curve closely matches the tabulated AUROC when integrated, and the mean precision–recall curve is a pointwise average over a common recall grid, which is why it is less smooth at low recall where few replicates contribute.

The tables and curves in this section are retained as a description of that control pool and of how much a single control draw can move the result.

Table S14: Per-draw AUROC, AUPRC, and F1 for 3-year myocardial infarction (MI) incidence, external transfer to UK Biobank (RFM, five balanced test-control draws). All incident events appear in every draw; the draws differ only in which controls were randomly sampled, so they are repeated draws from the control pool rather than independent cross-validation folds. These rows describe the variability of a single finite balanced evaluation; the main paper targets a different quantity, discrimination in the scored population at a 50% prevalence, and neither averages these values nor derives its interval from their spread (main paper Section 4.4.1).

| Draw | AUROC ↑ (95 % CI) | AUPRC ↑ (95 % CI) | F1 ↑ (95 % CI) |
|---|---|---|---|
| Draw 1 | 0.642 (0.569–0.716) | 0.632 (0.563–0.711) | 0.671 (0.609–0.728) |
| Draw 2 | 0.639 (0.561–0.711) | 0.606 (0.539–0.697) | 0.681 (0.613–0.739) |
| Draw 3 | 0.737 (0.668–0.806) | 0.714 (0.639–0.799) | 0.708 (0.646–0.760) |
| Draw 4 | 0.661 (0.589–0.732) | 0.636 (0.566–0.721) | 0.695 (0.630–0.755) |
| Draw 5 | 0.647 (0.570–0.720) | 0.627 (0.556–0.707) | 0.683 (0.615–0.737) |

Table S15: Per-draw AUROC, AUPRC, and F1 for 3-year ischaemic stroke incidence, external transfer to UK Biobank (RFM, five balanced test-control draws). All incident events appear in every draw; the draws differ only in which controls were randomly sampled, so they are repeated draws from the control pool rather than independent cross-validation folds. These rows describe the variability of a single finite balanced evaluation; the main paper targets a different quantity, discrimination in the scored population at a 50% prevalence, and neither averages these values nor derives its interval from their spread (main paper Section 4.4.1).

| Draw | AUROC ↑ (95 % CI) | AUPRC ↑ (95 % CI) | F1 ↑ (95 % CI) |
|---|---|---|---|
| Draw 1 | 0.690 (0.623–0.754) | 0.683 (0.618–0.756) | 0.694 (0.639–0.746) |
| Draw 2 | 0.684 (0.613–0.749) | 0.668 (0.599–0.744) | 0.692 (0.638–0.746) |
| Draw 3 | 0.645 (0.571–0.713) | 0.621 (0.555–0.704) | 0.684 (0.624–0.736) |
| Draw 4 | 0.716 (0.648–0.780) | 0.714 (0.641–0.793) | 0.694 (0.639–0.747) |
| Draw 5 | 0.692 (0.626–0.757) | 0.690 (0.621–0.765) | 0.684 (0.627–0.739) |

Table S16: Per-draw AUROC, AUPRC, and F1 for 3-year Parkinson's disease incidence, external transfer to UK Biobank (RFM, five balanced test-control draws). All incident events appear in every draw; the draws differ only in which controls were randomly sampled, so they are repeated draws from the control pool rather than independent cross-validation folds. These rows describe the variability of a single finite balanced evaluation; the main paper targets a different quantity, discrimination in the scored population at a 50% prevalence, and neither averages these values nor derives its interval from their spread (main paper Section 4.4.1).

| Draw | AUROC ↑ (95 % CI) | AUPRC ↑ (95 % CI) | F1 ↑ (95 % CI) |
|---|---|---|---|
| Draw 1 | 0.545 (0.289–0.793) | 0.536 (0.429–0.796) | 0.500 (0.211–0.750) |
| Draw 2 | 0.769 (0.537–0.942) | 0.786 (0.606–0.949) | 0.556 (0.235–0.818) |
| Draw 3 | 0.529 (0.264–0.785) | 0.503 (0.414–0.753) | 0.500 (0.200–0.737) |
| Draw 4 | 0.636 (0.364–0.876) | 0.566 (0.451–0.827) | 0.526 (0.181–0.769) |
| Draw 5 | 0.736 (0.504–0.934) | 0.770 (0.591–0.955) | 0.588 (0.222–0.824) |

### 5. Supplementary Figures: Predicted-vs-True Density Plots

Figures S1–S7 show predicted-vs-true density plots for all seven UK Biobank regression biomarkers reported in the main paper and Section 3 above. Each panel is a 2D kernel density estimate of one model's predicted value (vertical axis) against the ground-truth value (horizontal axis); the red dashed line marks perfect prediction ($y = x$), and the per-model MAE and Spearman $\rho$ are annotated in the panel title. Each figure shows RFM and the three RETFound variants, the published retinal foundation models it is compared against. The two readings of the DINOv3-Base checkpoint from which RFM is initialised (DINOv3b-9L and DINOv3b) are ablations of RFM itself, analysed in Sections 5.1–5.2 of the main paper, and are not shown here. MAE and $\rho$ for every model are given in Tables 1–5 of the main paper and Tables S12–S13 above.

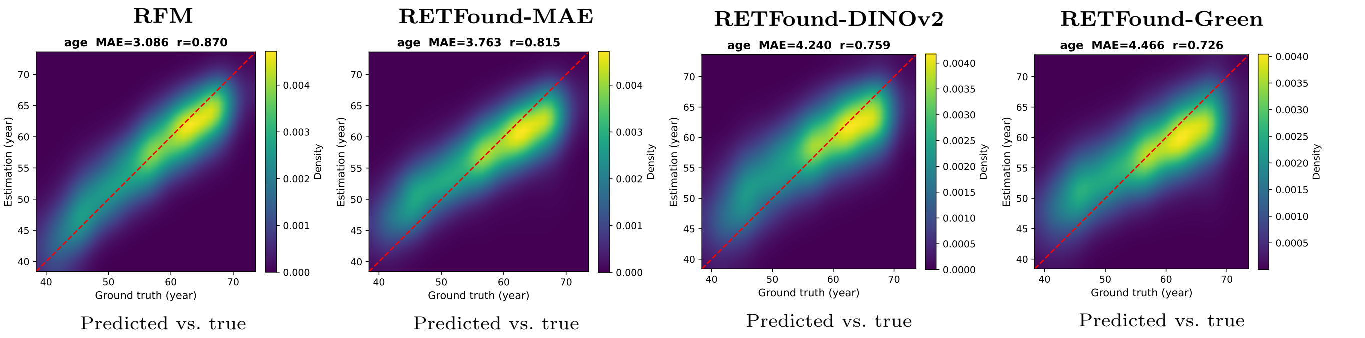


Figure S1: Predicted-vs-true density plots for age (years) regression on UK Biobank. Each panel is a 2D kernel density estimate of one model's predictions (vertical axis) against ground truth (horizontal axis); the red dashed line marks $y = x$. Panel titles give MAE and Spearman $\rho$. Shown for RFM and the three RETFound variants (Section 5).

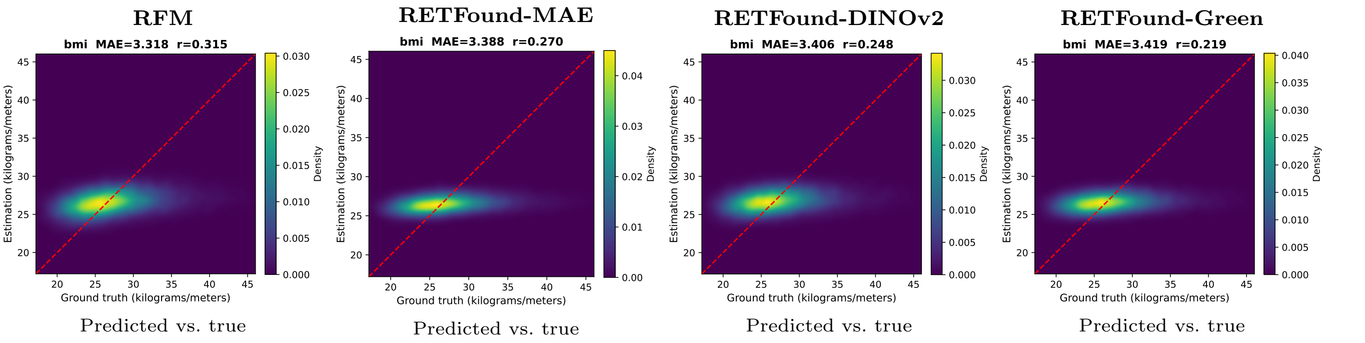


Figure S2: Predicted-vs-true density plots for BMI regression on UK Biobank. Each panel is a 2D kernel density estimate of one model's predictions (vertical axis) against ground truth (horizontal axis); the red dashed line marks $y = x$. Panel titles give MAE and Spearman $\rho$. Shown for RFM and the three RETFound variants (Section 5).

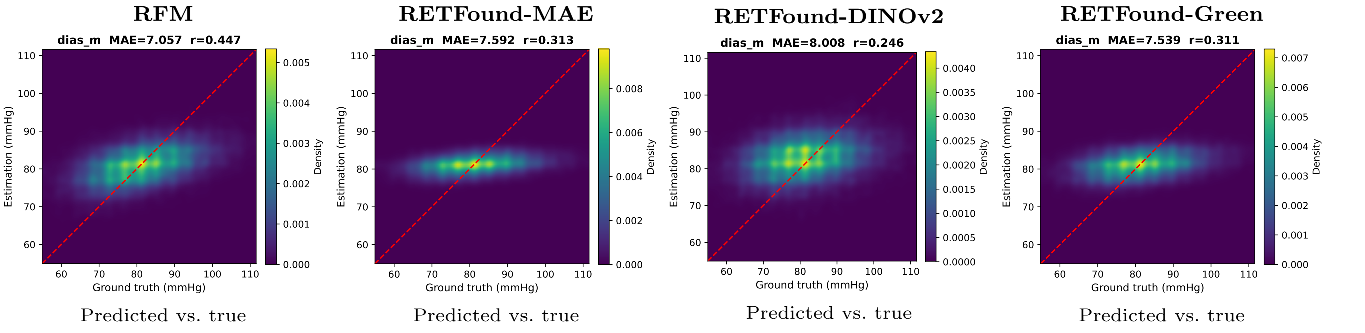


Figure S3: Predicted-vs-true density plots for diastolic blood pressure (DBP) regression on UK Biobank. Each panel is a 2D kernel density estimate of one model's predictions (vertical axis) against ground truth (horizontal axis); the red dashed line marks $y = x$. Panel titles give MAE and Spearman $\rho$. Shown for RFM and the three RETFound variants (Section 5).

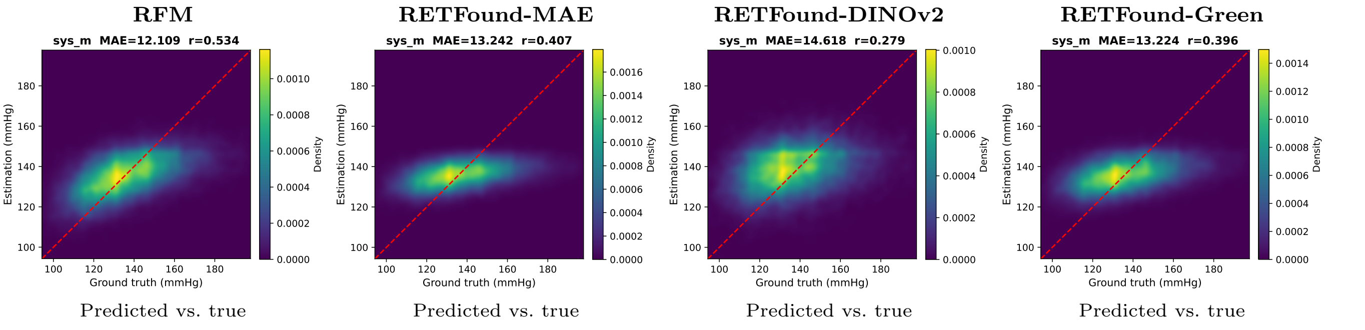


Figure S4: Predicted-vs-true density plots for systolic blood pressure (SBP) regression on UK Biobank. Each panel is a 2D kernel density estimate of one model's predictions (vertical axis) against ground truth (horizontal axis); the red dashed line marks $y = x$. Panel titles give MAE and Spearman $\rho$. Shown for RFM and the three RETFound variants (Section 5).

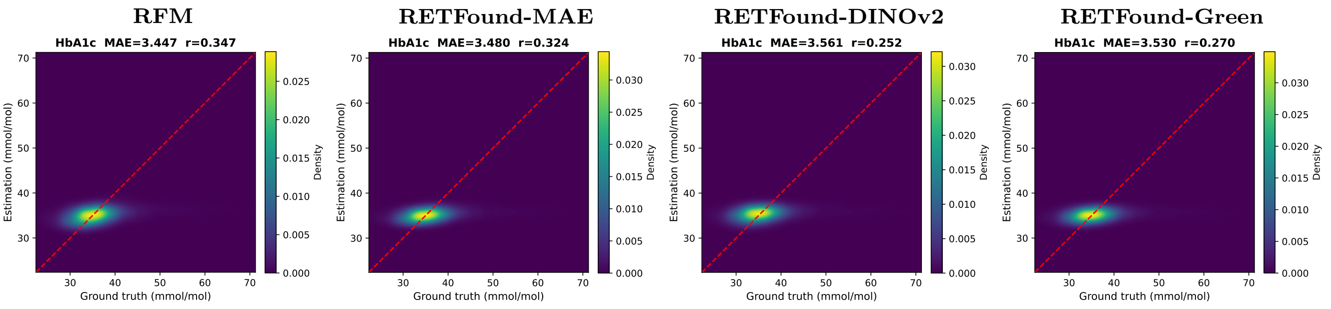


Predicted vs. true Predicted vs. true Predicted vs. true Predicted vs. true

Figure S5: Predicted-vs-true density plots for HbA1c regression on UK Biobank. Each panel is a 2D kernel density estimate of one model's predictions (vertical axis) against ground truth (horizontal axis); the red dashed line marks $y = x$. Panel titles give MAE and Spearman $\rho$. Shown for RFM and the three RETFound variants (Section 5).

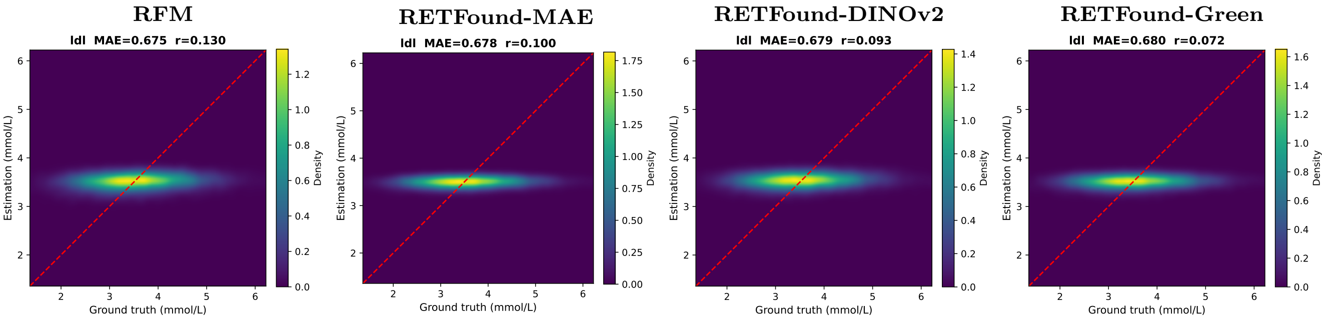


Predicted vs. true Predicted vs. true Predicted vs. true Predicted vs. true

Figure S6: Predicted-vs-true density plots for LDL cholesterol regression on UK Biobank. Each panel is a 2D kernel density estimate of one model's predictions (vertical axis) against ground truth (horizontal axis); the red dashed line marks $y = x$. Panel titles give MAE and Spearman $\rho$. Shown for RFM and the three RETFound variants (Section 5).

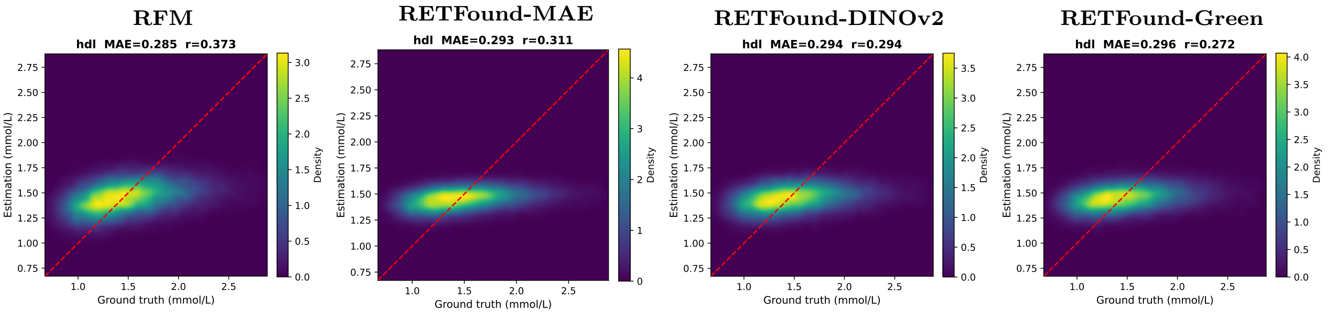


Predicted vs. true Predicted vs. true Predicted vs. true Predicted vs. true

Figure S7: Predicted-vs-true density plots for HDL cholesterol regression on UK Biobank. Each panel is a 2D kernel density estimate of one model's predictions (vertical axis) against ground truth (horizontal axis); the red dashed line marks $y = x$. Panel titles give MAE and Spearman $\rho$. Shown for RFM and the three RETFound variants (Section 5).

### 6. Supplementary Figures: Per-Draw ROC/PR Curves for 3-Year Incidence

Figures S8–S13 show the individual ROC and precision–recall (PR) curves for each of the five balanced test-control draws made for the RFM external 3-year incidence evaluation, corresponding row-for-row to Tables S14–S16 above. Each panel is labelled with its draw number.

These are not the curves reported in the main paper. The main paper's Figures 8–10 instead show, for each endpoint, the mean ROC and PR curve over bootstrap replicates of the whole evaluation together with the 2.5–97.5 percentile band from which the tabulated confidence intervals are taken (main paper Section 4.4.1). The curves below are shown because they make the practical consequence visible: the five draws differ only in which controls were sampled, yet the resulting curves are visibly different, which is why a single control draw is not a sound basis for a reported result and why the main paper bootstraps over the pooled control set instead.

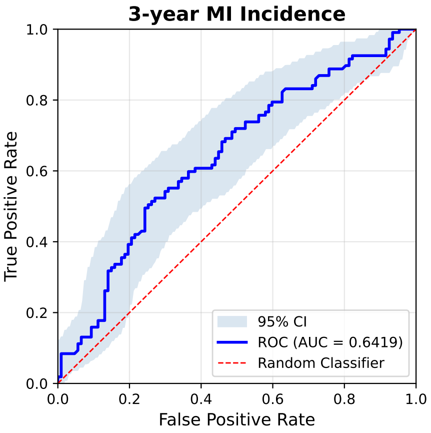


Draw 1

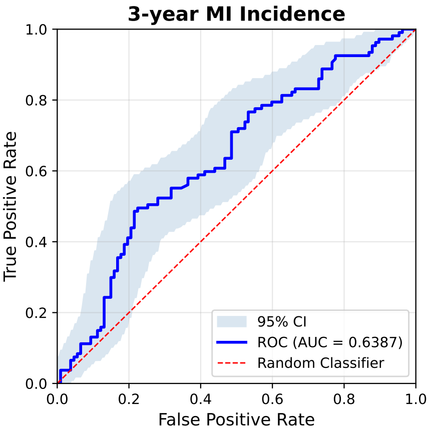


Draw 2

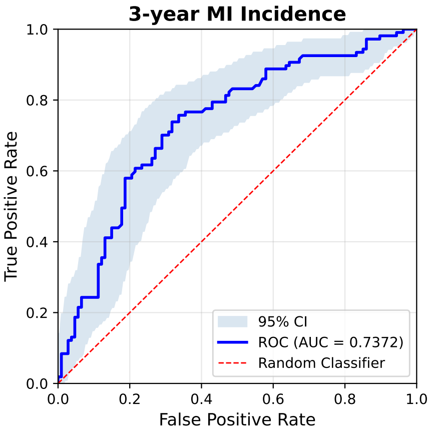


Draw 3

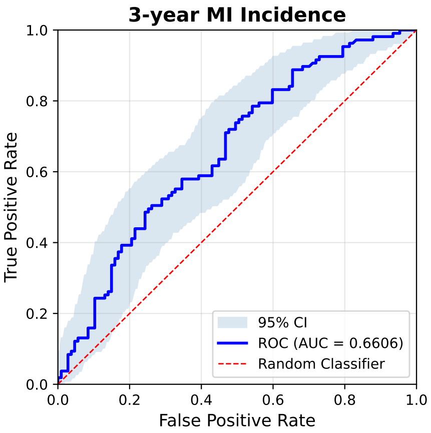


Draw 4

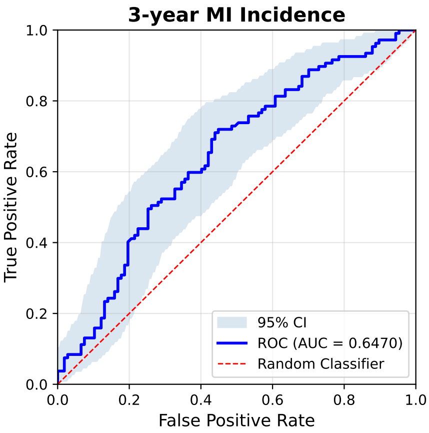


Draw 5

Figure S8: Per-draw ROC curves for 3-year myocardial infarction (MI) incidence, external transfer to UK Biobank (RFM, all five balanced test-control draws; shaded bands show bootstrap 95% confidence intervals where available). All incident events are present in every draw; the draws differ only in which controls were sampled. The corresponding main-paper figure instead shows the mean curve over bootstrap replicates of the whole evaluation, with its percentile band (main paper Section 4.4.1).

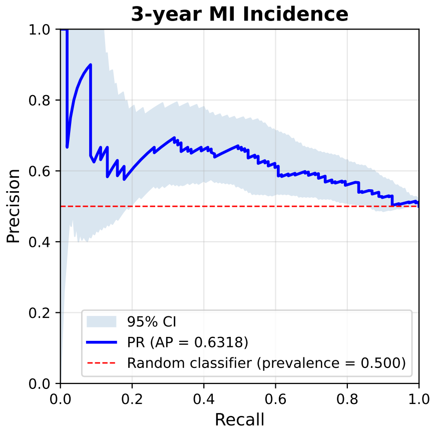


Draw 1

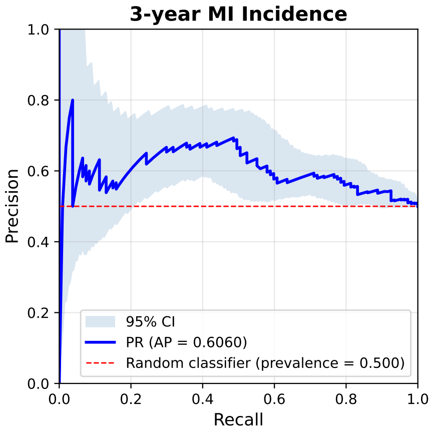


Draw 2

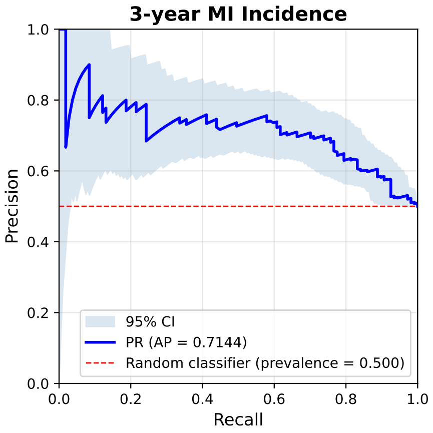


Draw 3

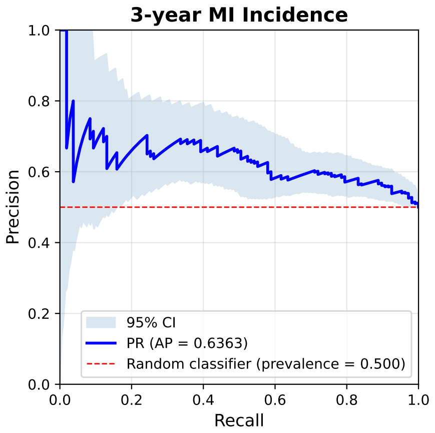


Draw 4

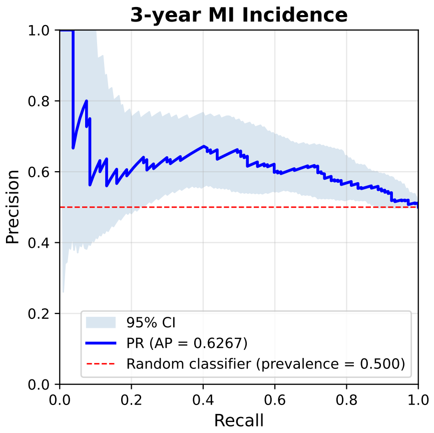


Draw 5

Figure S9: Per-draw PR curves for 3-year myocardial infarction (MI) incidence, external transfer to UK Biobank (RFM, all five balanced test-control draws; shaded bands show bootstrap 95% confidence intervals where available). All incident events are present in every draw; the draws differ only in which controls were sampled. The corresponding main-paper figure instead shows the mean curve over bootstrap replicates of the whole evaluation, with its percentile band (main paper Section 4.4.1).

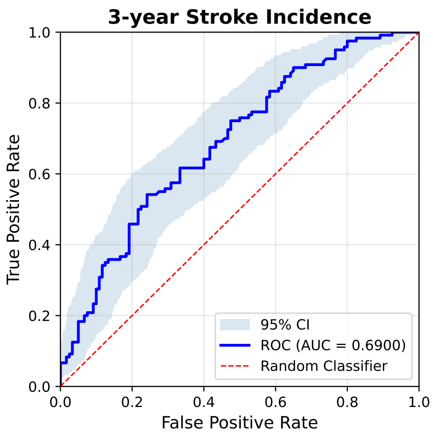


Draw 1

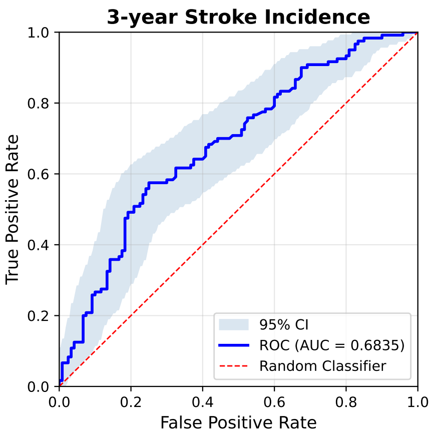


Draw 2

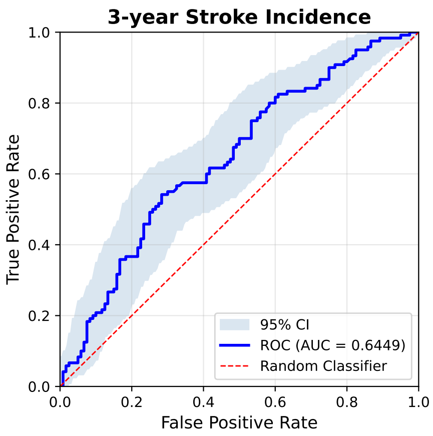


Draw 3

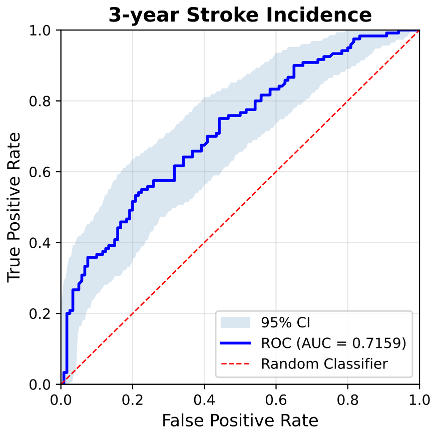


Draw 4

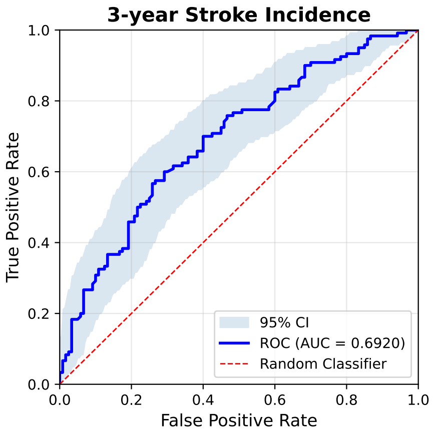


Draw 5

Figure S10: Per-draw ROC curves for 3-year ischaemic stroke incidence, external transfer to UK Biobank (RFM, all five balanced test-control draws; shaded bands show bootstrap 95% confidence intervals where available). All incident events are present in every draw; the draws differ only in which controls were sampled. The corresponding main-paper figure instead shows the mean curve over bootstrap replicates of the whole evaluation, with its percentile band (main paper Section 4.4.1).

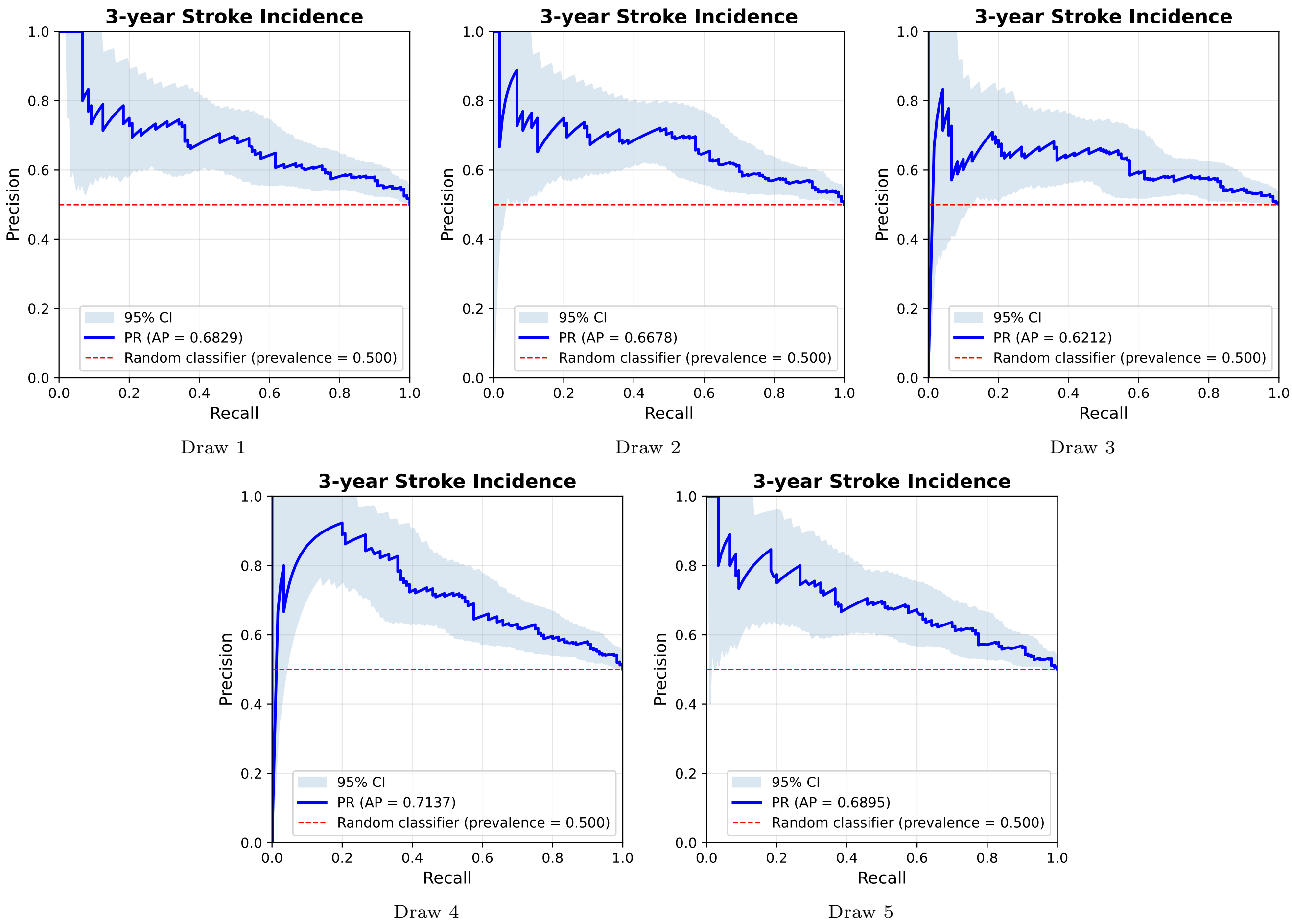


Figure S11: Per-draw PR curves for 3-year ischaemic stroke incidence, external transfer to UK Biobank (RFM, all five balanced test-control draws; shaded bands show bootstrap 95% confidence intervals where available). All incident events are present in every draw; the draws differ only in which controls were sampled. The corresponding main-paper figure instead shows the mean curve over bootstrap replicates of the whole evaluation, with its percentile band (main paper Section 4.4.1).

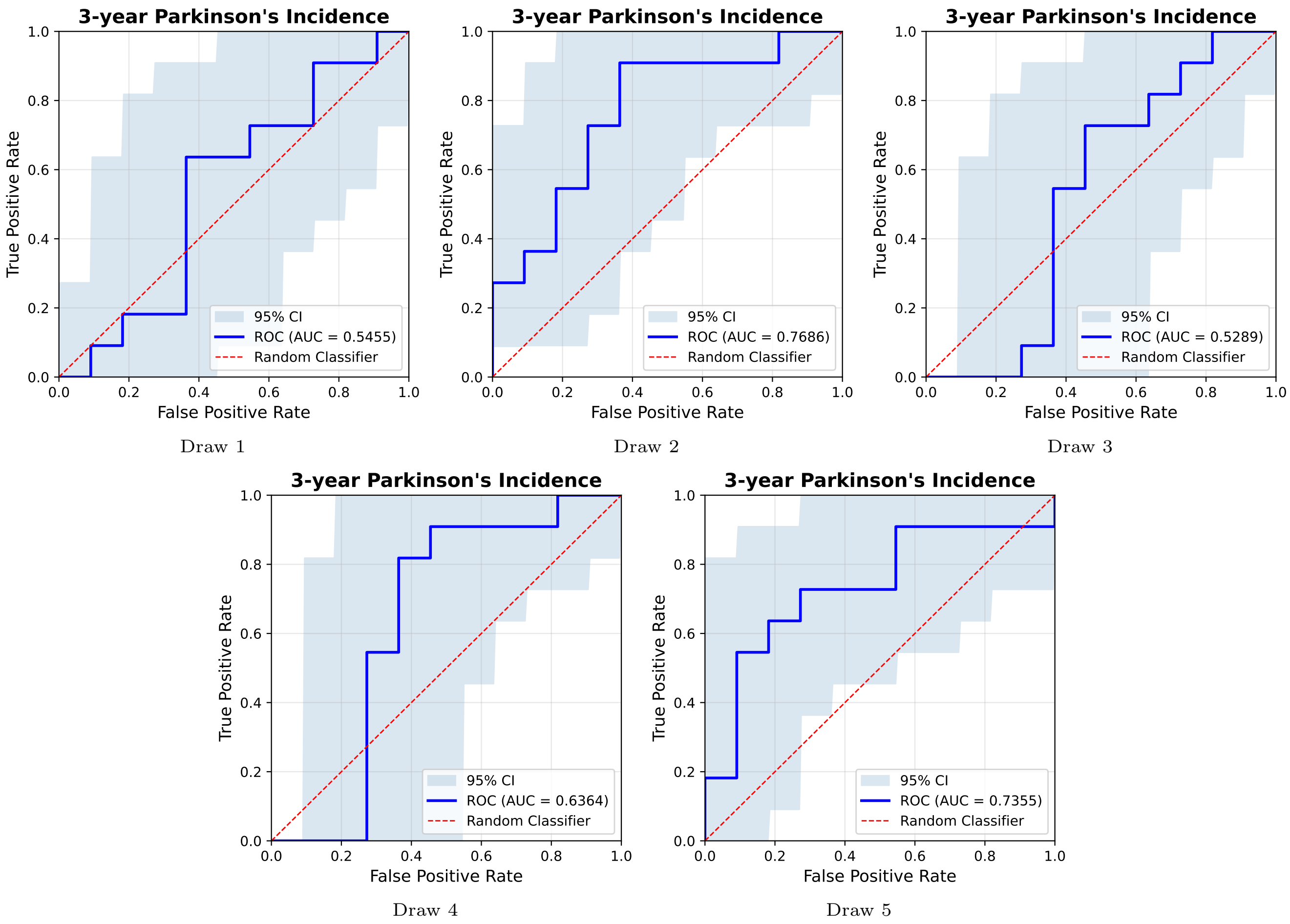


Figure S12: Per-draw ROC curves for 3-year Parkinson's disease incidence, external transfer to UK Biobank (RFM, all five balanced test-control draws; shaded bands show bootstrap 95% confidence intervals where available). All incident events are present in every draw; the draws differ only in which controls were sampled. The corresponding main-paper figure instead shows the mean curve over bootstrap replicates of the whole evaluation, with its percentile band (main paper Section 4.4.1).

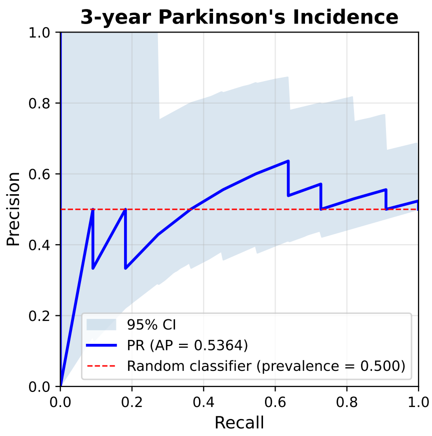


Draw 1

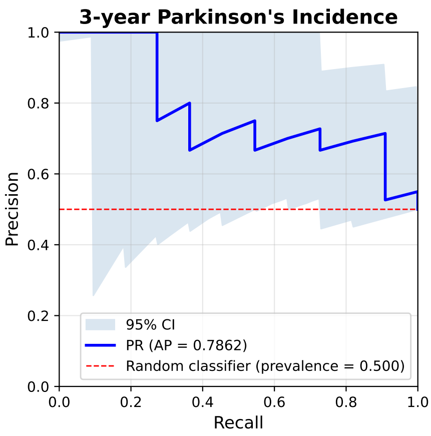


Draw 2

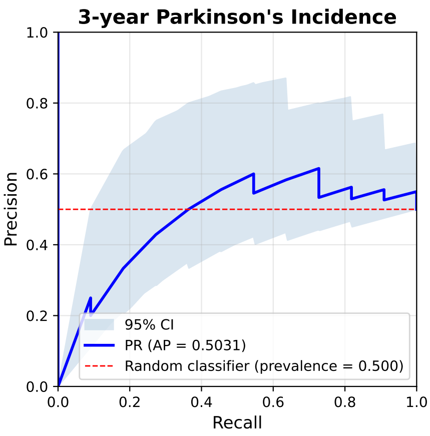


Draw 3

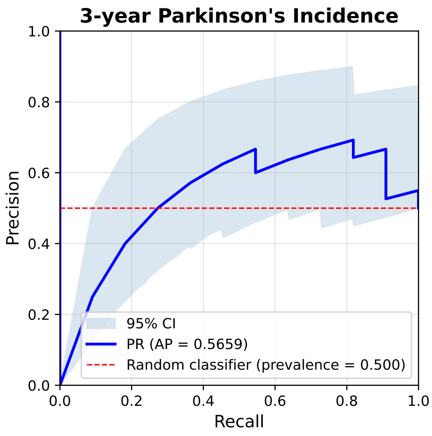


Draw 4

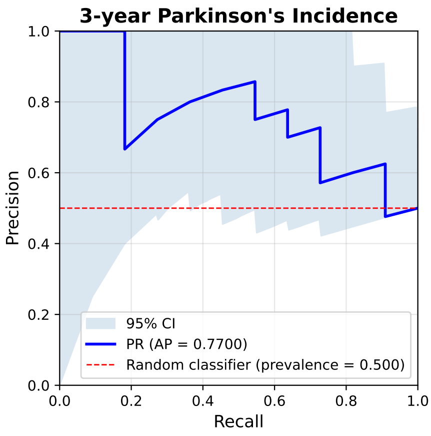


Draw 5

Figure S13: Per-draw PR curves for 3-year Parkinson's disease incidence, external transfer to UK Biobank (RFM, all five balanced test-control draws; shaded bands show bootstrap 95% confidence intervals where available). All incident events are present in every draw; the draws differ only in which controls were sampled. The corresponding main-paper figure instead shows the mean curve over bootstrap replicates of the whole evaluation, with its percentile band (main paper Section 4.4.1).

### 7. Supplementary Figures: Sex and Smoking-Status Classification

Figures S14–S15 show ROC and precision–recall (PR) curves and confusion matrices for sex and smoking-status classification on UK Biobank (RFM only; Tables 6 and 8). These two phenotypes are omitted from the main paper's figures for space; the corresponding figure for ethnicity classification, the phenotype for which RFM establishes a new reference point in the absence of prior published results (Section 4.1.2), is retained in the main paper.

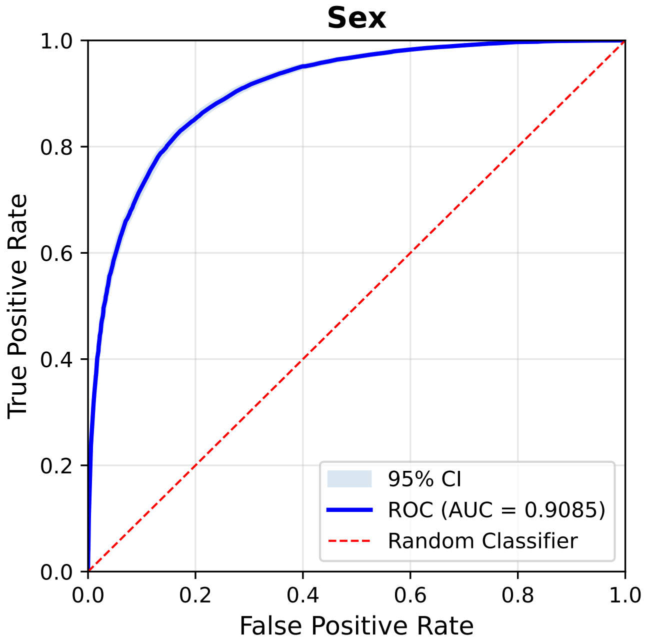


ROC

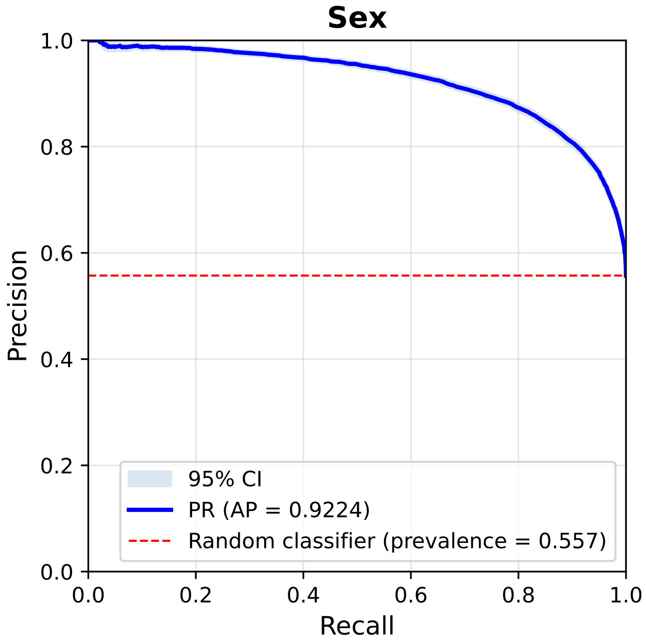


PR

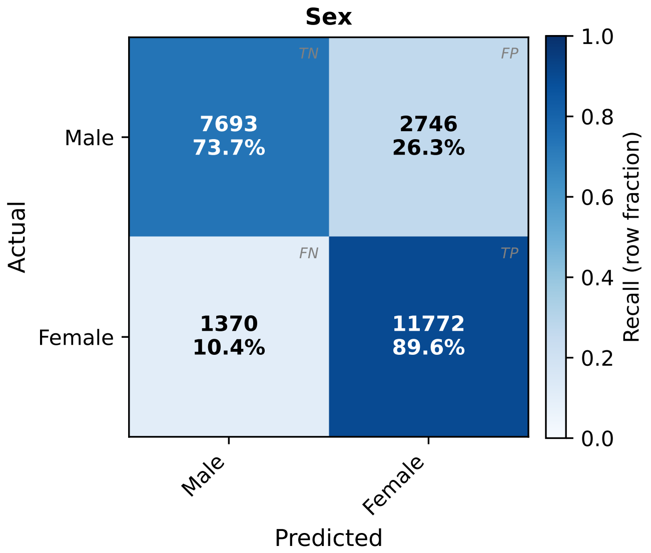


Confusion matrix

Figure S14: Classification diagnostics for sex classification on UK Biobank (RFM only). Top row: ROC and precision–recall curves, with shaded 95% confidence intervals where available. Bottom: confusion matrix at the 0.5 decision threshold on the positive-class probability (rows: true class; columns: predicted class).

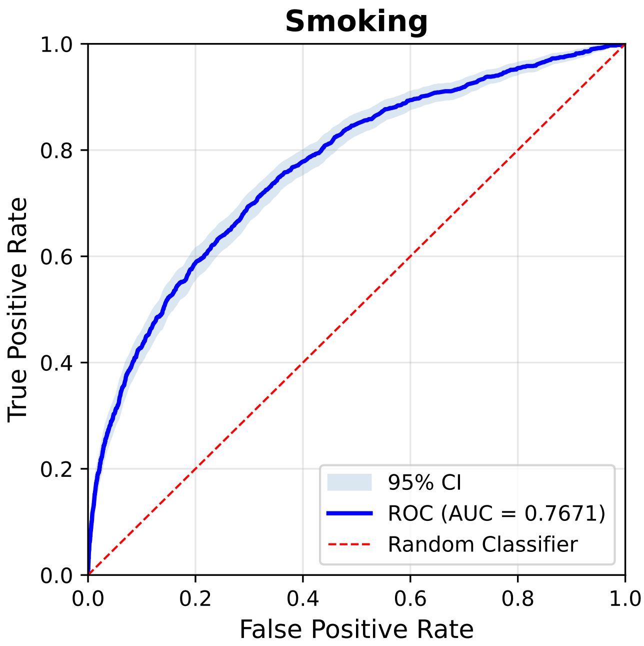


ROC

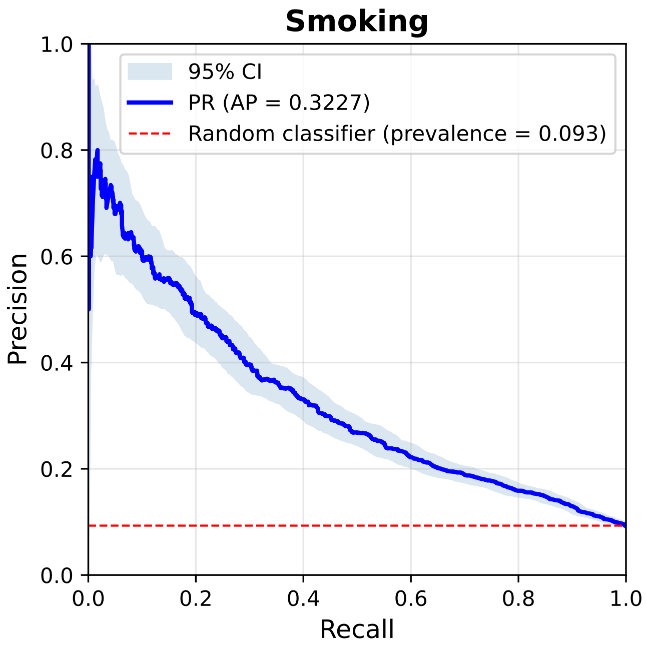


PR

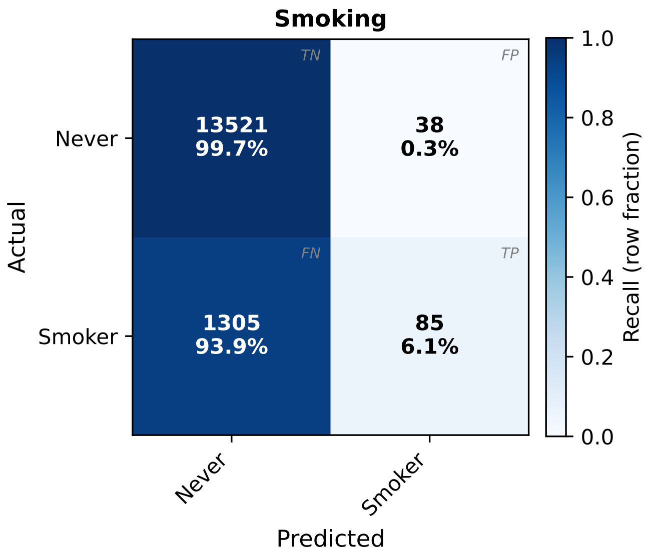


Confusion matrix

Figure S15: Classification diagnostics for smoking-status classification on UK Biobank (RFM only). Top row: ROC and precision–recall curves, with shaded 95% confidence intervals where available. Bottom: confusion matrix at the 0.5 decision threshold on the positive-class probability (rows: true class; columns: predicted class).

### 8. Supplementary Figures: Additional Glaucoma and DR Diagnostics

Figures S16 and S21 show precision–recall (PR) curves complementing the ROC curves and confusion matrices reported in the main paper for GFID glaucoma detection and IDRiD DR grading (Sections 4.2–4.3); PR curves are more sensitive than ROC to performance on the minority (positive) class under class imbalance. For PAPILA glaucoma detection (Figures S17–S18) and APTOS 2019 and Messidor-2 DR grading (Figures S19–S20 and S22–S23), the full diagnostic figure (ROC curves, confusion matrices, and PR curves) is reported here rather than in the main paper for space; GFID and IDRiD are retained in the main paper as representative examples of each comparison (glaucoma detection and DR grading, the latter discussed in Section 5.2 of the main paper).

**RFM**

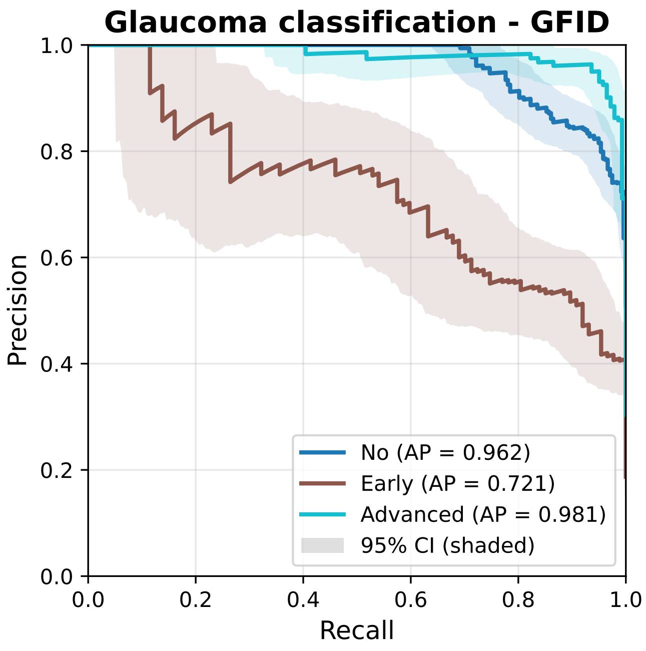


AUPRC

**DINOv3b-9L**

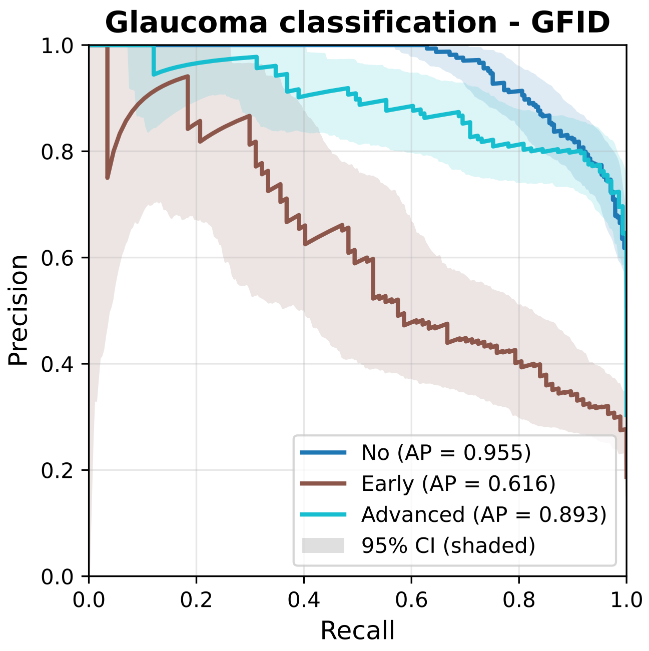


AUPRC

Figure S16: Per-class precision–recall curves for glaucoma detection on GFID.

RFM

DINOv3b-9L

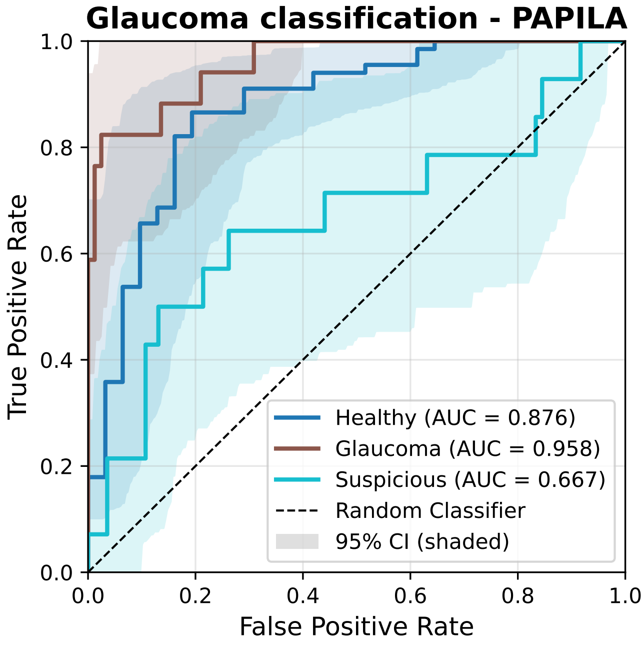


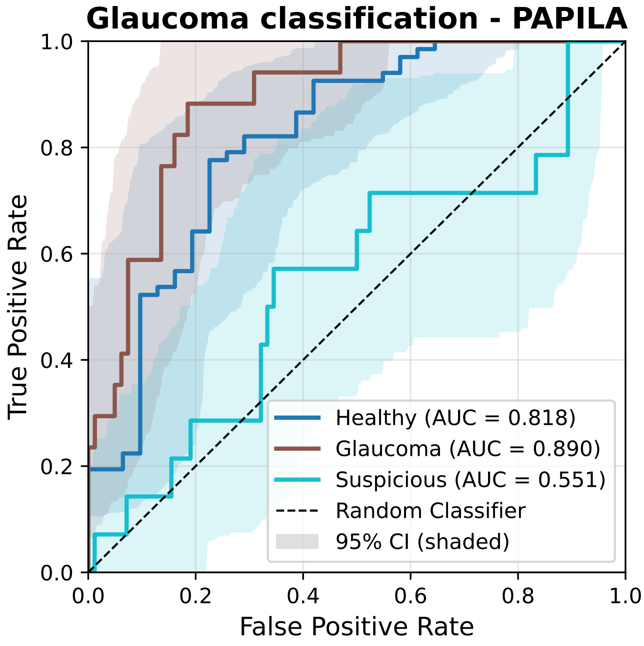


ROC

ROC

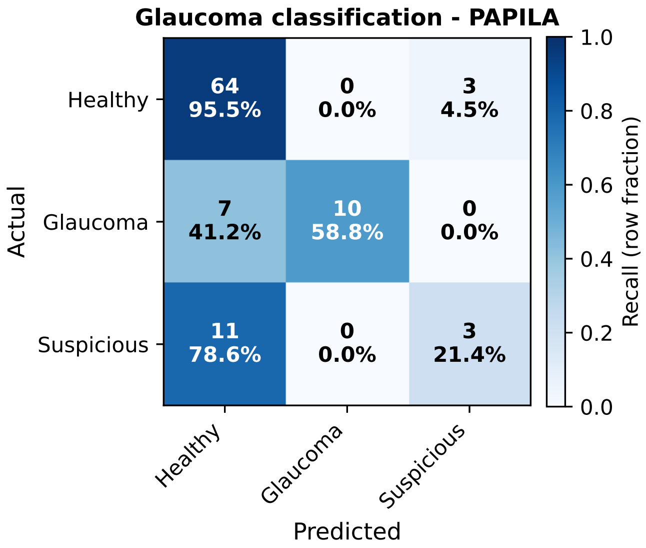


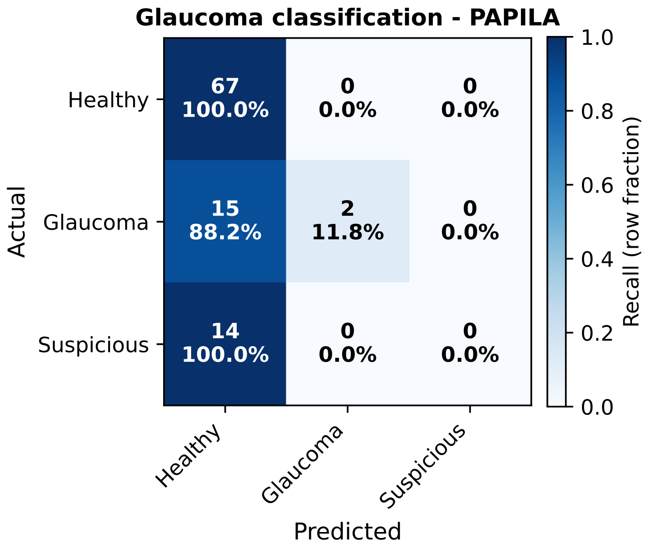


Confusion matrix

Confusion matrix

Figure S17: Per-class ROC curves (top row) and confusion matrices of the argmax prediction (bottom row; rows: true class, columns: predicted class) for glaucoma detection on PAPILA.

RFM

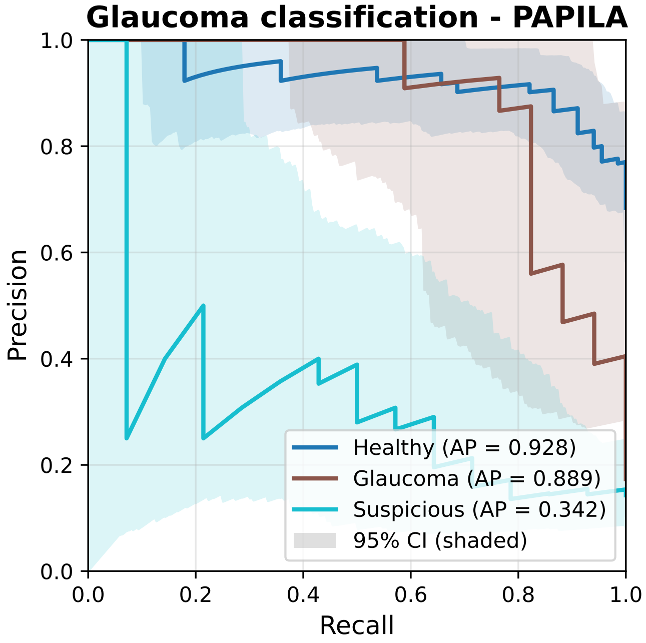


AUPRC

DINOv3b-9L

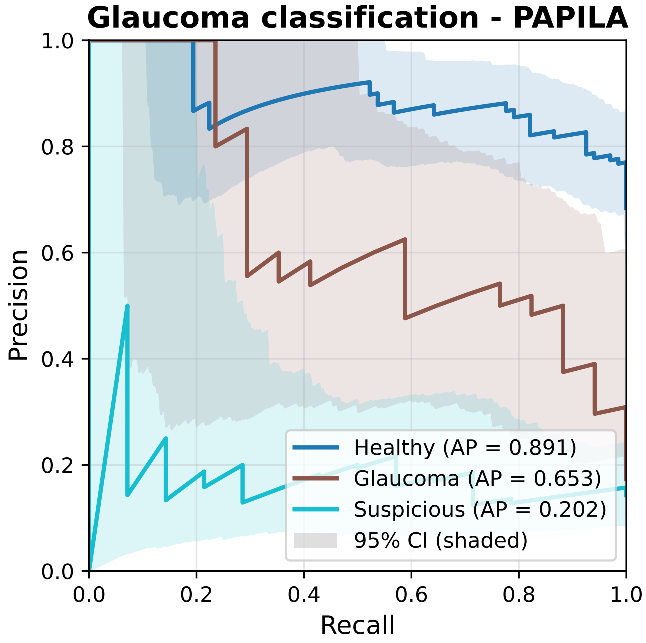


AUPRC

Figure S18: Per-class precision–recall curves for glaucoma detection on PAPILA.

RFM

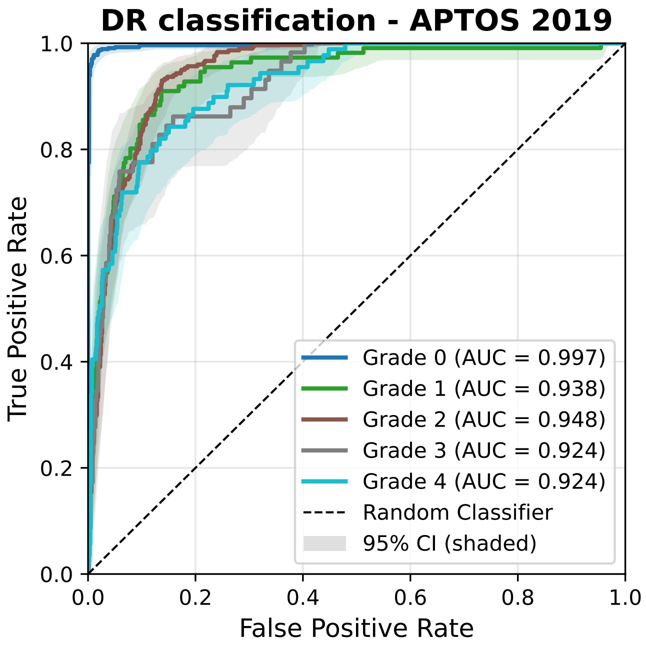


ROC

DINOv3b-9L

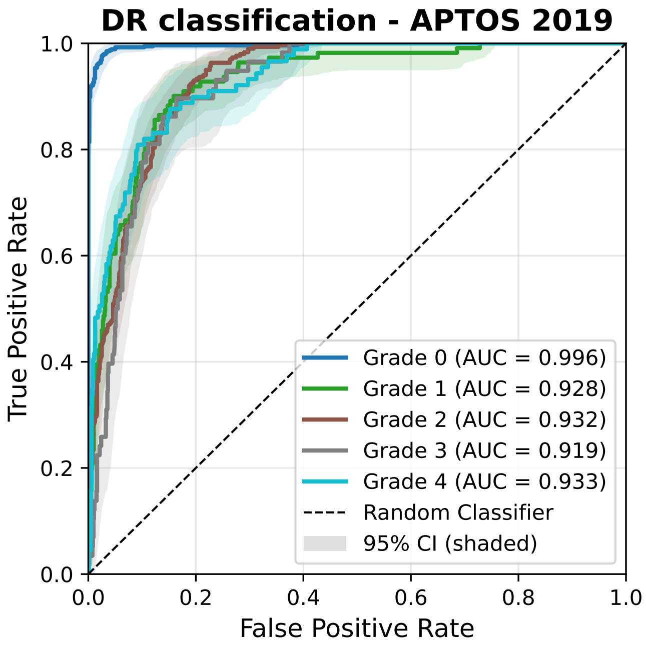


ROC

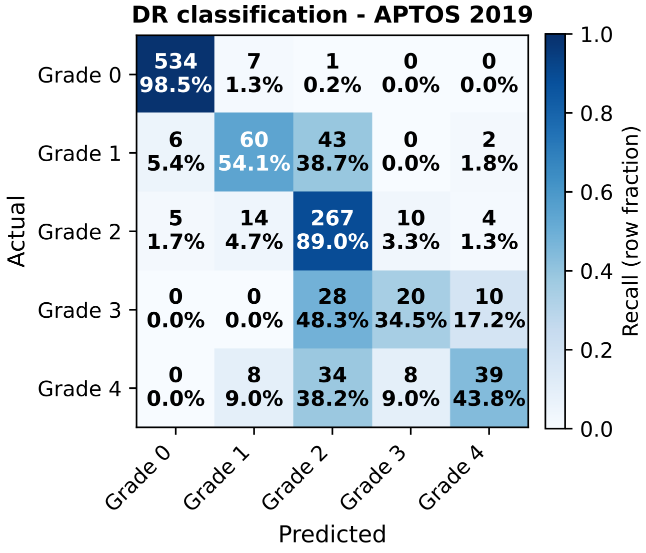


Confusion matrix

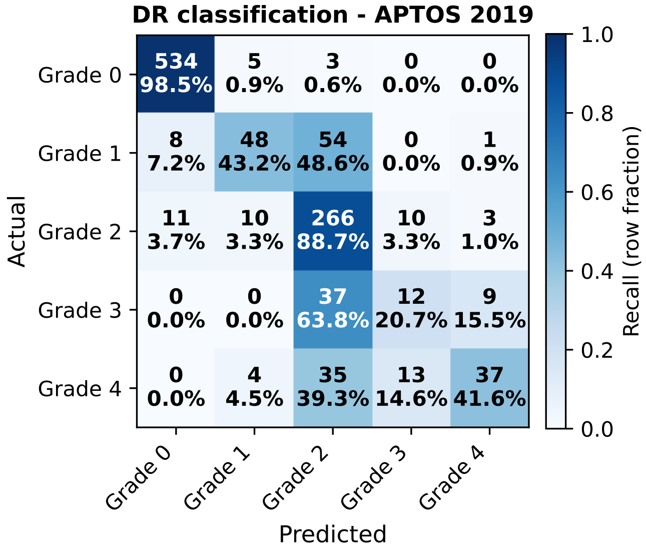


Confusion matrix

Figure S19: Per-class ROC curves (top row) and confusion matrices of the argmax prediction (bottom row; rows: true class, columns: predicted class) for diabetic retinopathy grading on APTOS 2019.

RFM DINOv3b-9L

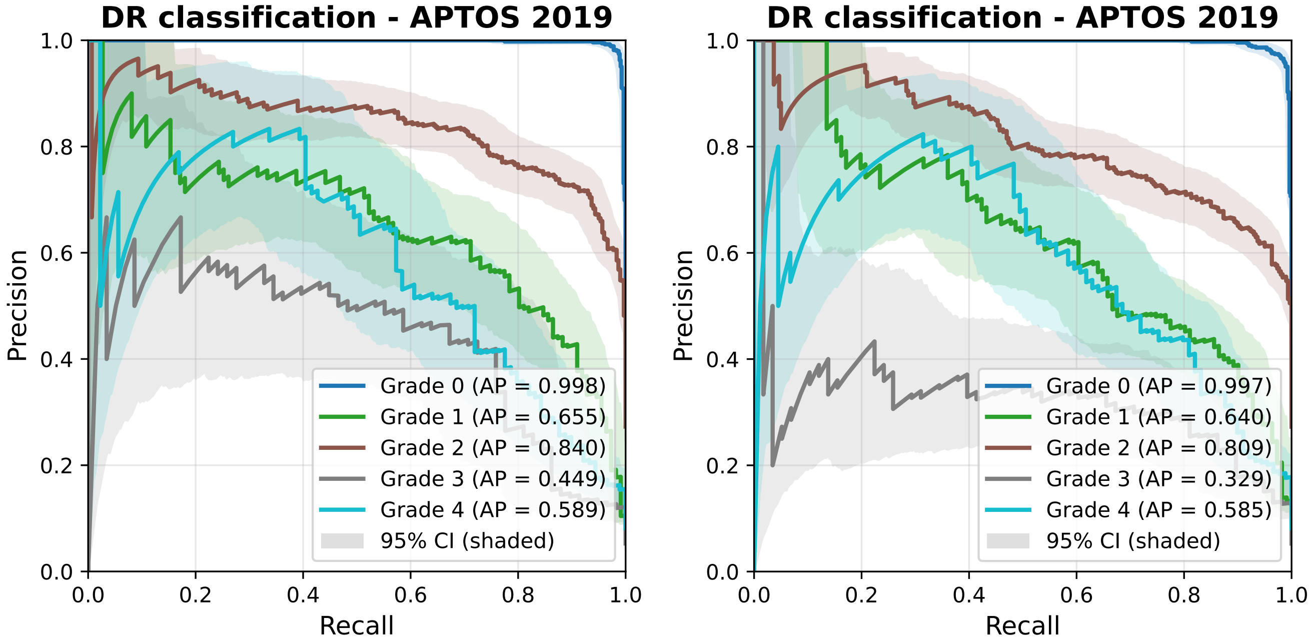


AUPRC AUPRC

Figure S20: Per-class precision–recall curves for diabetic retinopathy grading on APTOS 2019.

RFM DINOv3b-9L

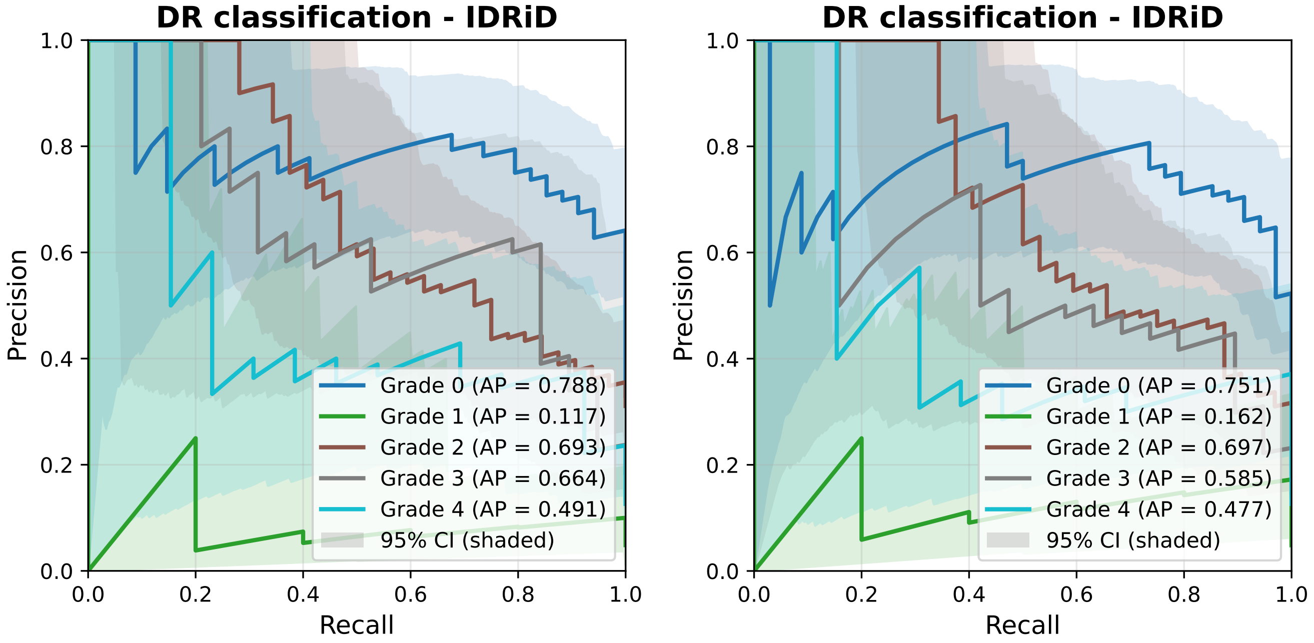


AUPRC AUPRC

Figure S21: Per-class precision–recall curves for diabetic retinopathy grading on IDRiD.

RFM

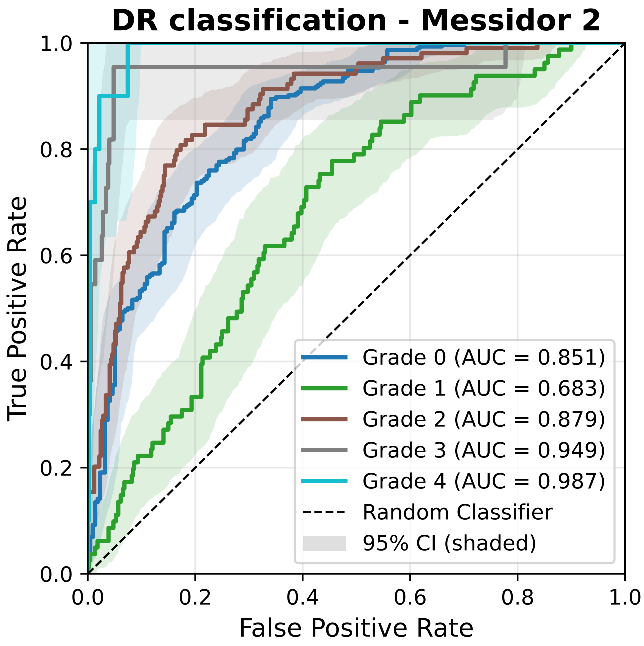


ROC

DINOv3b-9L

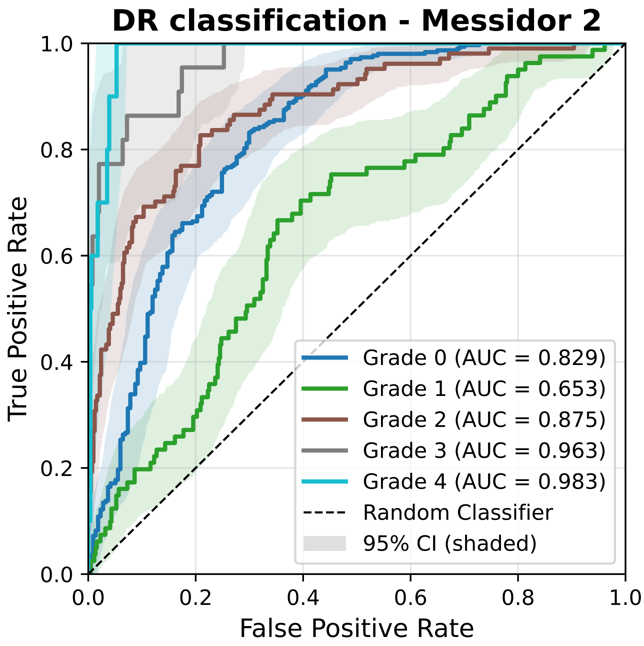


ROC

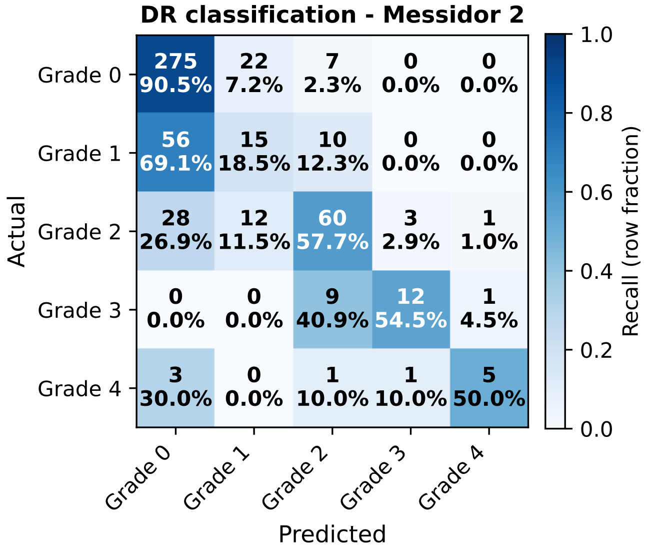


Confusion matrix

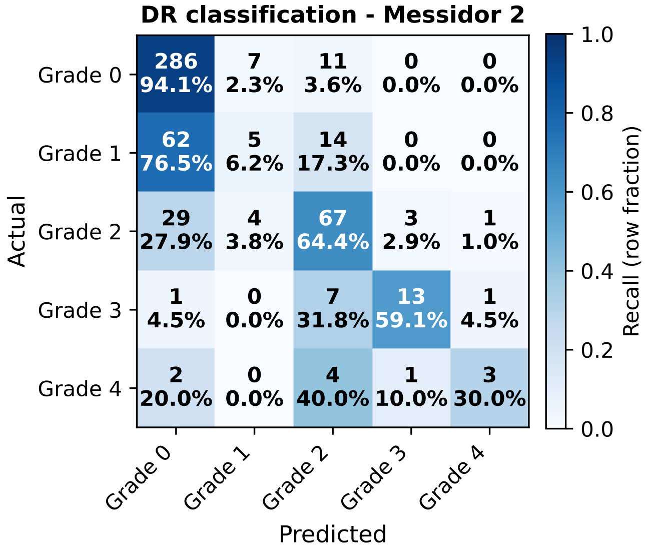


Confusion matrix

Figure S22: Per-class ROC curves (top row) and confusion matrices of the argmax prediction (bottom row; rows: true class, columns: predicted class) for diabetic retinopathy grading on Messidor-2.

**RFM**

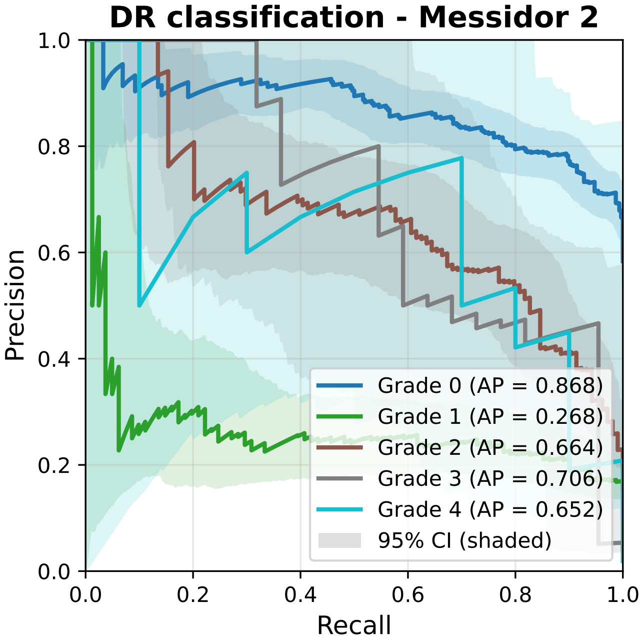


AUPRC

**DINOv3b-9L**

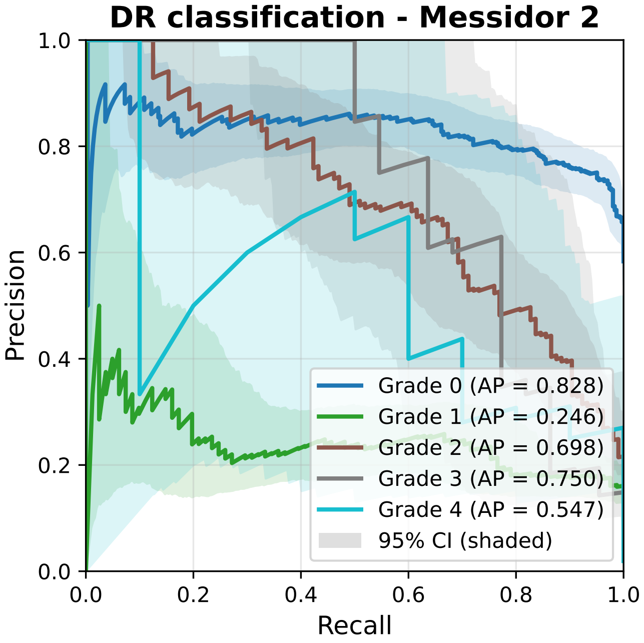


AUPRC

Figure S23: Per-class precision–recall curves for diabetic retinopathy grading on Messidor-2.
